# The complete blood count and cardiovascular disease: analyses across six cohorts of 23,370 adults

**DOI:** 10.1101/2024.10.17.24315694

**Authors:** Sascha N. Goonewardena, Venkatesh L. Murthy

## Abstract

**Background:** The complete blood count (CBC) is one of the most performed laboratory studies. However, the CBC and its components are not commonly used to understand and quantify cardiovascular disease (CVD) risk.

**Objective:** We sought to define the relationships between the CBC, traditional CVD risk factors, and common CVD biomarkers and their joint association with all-cause mortality and CVD.

**Methods:** We examined the relationships between the CBC, traditional CVD risk factors, and mortality in NHANES (n=7843). We validated and extended these findings to more refined CVD endpoints in five additional cohorts (n=15,527).

**Results:** We first examined the variance accounted for by common laboratory studies (lipid panel, HbA1c, hs-CRP, and basic metabolic panel) by traditional risk factors in NHANES. Except for hemoglobin (Hb) components, we found that traditional risk factors accounted for less than 20% of the variance in the CBC, suggesting that the CBC provides unique information beyond traditional risk factors and CVD biomarkers. Additionally, the CBC was strongly associated with all-cause mortality (p<0.0001), even more than traditional CVD biomarkers (lipid panel, HbA1c, and CRP). We validated and extended these findings across five additional cohorts with a mean follow-up of 16 years and more refined CVD endpoints. In the fully adjusted analyses, several CBC components, including the white blood cell (WBC) count, neutrophil (PMN) count, Hb level, and an integrated immune cell score, were associated with individual CVD endpoints (incident stroke, MI, or revascularization) and a composite CV endpoint (MACE3) with standardized hazard ratios of 1.13 (p=0.002), 1.15 (p=0.0006), 0.82 (p<0.0001), and 2.16 (p<0.0001) respectively.

**Conclusion:** This study represents the first systematic examination of the relationship between the CBC, all-cause mortality, and CVD in a diverse cohort of 23,370 adults. These findings underscore the added value of the CBC over traditional risk factors and common CVD biomarkers for CVD risk assessment. Future studies should explore the integration of CBC parameters into predictive models to enhance our understanding, early identification, and prevention strategies for CVD.

## Introduction

Traditional risk factors for CVD include age, sex, and smoking.^1,2^ In addition to traditional demographic risk factors, quantitative markers including hemoglobin A1c (HbA1c), creatinine (Cr), and low-density lipoprotein cholesterol (LDL-C) can be used to estimate CVD risk.^3,4^ Importantly, quantitative markers such as LDL-C and C-reactive protein (CRP) have not only advanced our risk assessment but also have provided critical insights into the biological mechanisms of CVD and have informed the development of cardiovascular therapies.^5,6^ The complete blood count (CBC) is the laboratory study used to quantify circulating cells (CC). Although the CBC is one of the most frequently performed laboratory studies in the United States, there is a paucity of information on the utility of the CBC for quantitative CVD risk assessment.^7^

Most laboratory studies measure circulating molecular mediators (CMM) of CVD (e.g., Cr, CRP, HbA1c, and LDL-C). CC can also drive CVD, and the CBC is the only common laboratory study that profiles CC.^8^ However, little is known about the relationship between traditional CVD risk factors and the CBC and how they together contribute to CVD risk.

Each of the three major components of the CBC (i.e., RBCs, platelets, and WBCs) has been associated with CVD (primarily in the secondary prevention setting), including atherosclerotic cardiovascular disease (ASCVD).^9^ Specifically, oxygen-carrying capacity (RBC component), thrombosis (platelet component), and inflammation (WBC component) are all intimately connected to atherogenesis and specific CVD phenotypes.^10^ With respect to oxygen-carrying capacity, several studies have found associations between RBC counts and CVD. For example, Sabatine and colleagues found that anemia (low RBC counts) was an independent predictor of MACE in acute coronary syndromes.^11^ Platelet components have not only been associated with CVD risk but also are canonical cell targets to reduce CVD risk.^12^ Inflammation is associated with the initiation, progression, and clinical manifestations of ASCVD and CVD.^13^ More than simply an association, several randomized control trials (RCT) of anti-inflammatory therapies have yielded promising results for CVD.^14–16^

Although individual CBC components have been associated with CVD, the influence of traditional risk factors on the CBC and the effect of CBC components on incident CVD endpoints in diverse, stable patients remain largely unknown. To further elucidate these relationships, we examined the associations between the CBC, traditional risk factors, and mortality in NHANES (n=7843). We grouped CBC parameters from NHANES using principal component (PC) analysis. We then examined the relationships between individual CBC components and these PCs on incident CVD endpoints using adjusted Cox proportional hazards models in 15,527 individuals from five longitudinal cohorts (Framingham Offspring, Framingham 3rd Gen, MESA, CRIC, and CARDIA cohorts). We demonstrated that CBC components are powerful predictors of all-cause mortality and incident CVD in diverse adult populations, providing novel insights into the contributions of the CBC to the pathobiology and risk of CVD.

## Materials and Methods

### NHANES

#### Cohort description

The National Health and Nutrition Examination Survey (NHANES) is a biennial weighted survey with sampling weights designed to be representative of non-institutionalized residents of the United States. We obtained data from the 2003-4, 2005-6, 2007-8, and 2009-10 cycles of NHANES and linked mortality outcomes from the National Center for Health Statistics (https://www.cdc.gov/nchs/nhanes/index.htm). NHANES participants provided informed consent and the overall study was approved by the NCHS Research Ethics Review Board. Because NHANES data are publicly available this analysis was exempt from local IRB review.

#### Sample selection

Starting with 41,156 individual participants, we excluded individuals who were aged <20 years (N=18,983), who did not attend a fasting examination (N=11,935), and who had incomplete key covariates (N=959). We further excluded participants who did not have a complete blood count (CBC) (N=374) or differential (N=29), basic metabolic profile [BMP] (N=137), C-reactive protein (CRP) (N=3), hemoglobin A1c (HbA1c) (N=24), or lipid profile (N=214). We also excluded individuals with zero or missing weight in sampling weights computed for fasting participants (N=651) and individuals where the total of all components of the differential summed to <95% or 105% (N=4).

#### Laboratory measurements

We evaluated commonly obtained laboratory studies including fasting lipid profile (total cholesterol, HDL cholesterol, LDL cholesterol, and triglycerides), CBC with differential (hematocrit, hemoglobin, RBC count, WBC count, platelet count, red cell distribution width [RDW], mean corpuscular hemoglobin [MCH], mean corpuscular hemoglobin concentration [MCHC], lymphocyte percentage, neutrophil percentage, monocyte percentage, basophil percentage, eosinophil percentage, lymphocyte count, neutrophil count, monocyte count, basophil count, and eosinophil count), BMP (sodium, potassium, glucose, chloride, bicarbonate, blood urea nitrogen [BUN], creatinine, and total calcium), CRP, and HbA1c. Laboratory values were hyperbolic arcsine transformed to improve normality and scaled to zero mean and unit variance.

#### Variance explained in laboratory measurements by clinical parameters

We first evaluated the extent of independent information in each lab parameter by computing the variance of each transformed and scaled lab variable explained by standard clinical risk factors (*i.e.* age, sex, race, smoking, body mass index, systolic and diastolic blood pressure, use of antihypertensive medications, self-reported diabetes status) using generalized linear models accounting for fasting survey weights and type I analysis of variance.

#### Outcome analyses

We then related each laboratory panel (with CRP and HbA1c treated as separate panels) to all-cause mortality, cardiac mortality and cardiovascular mortality using Cox proportional hazard models, accounting for fasting survey weights. We computed unadjusted models as well as models adjusted for age, sex, and race as well as models adjusted for standard clinical risk factors (age, sex, race, smoking, body mass index, systolic and diastolic blood pressure, use of antihypertensive medications, and diabetes status).

#### Principal Component Analyses

Finally, we performed principal component analyses across all common laboratory panels as well as on each laboratory panel separately using varimax post-rotation, selecting the minimum number of components to explain at least 60% of total variance to group related measures within each panel and to reduce dimensionality. PCA scores were then computed for each participant and used as the key predictor in survival analysis repeated, analogous to above.

#### Repeatability of Laboratory Measures

To evaluate the stability of repeated laboratory measures over time, we leverage a subsample of the 2001-2 cycle of NHANES (N=551) who were asked to return within 8 days for repeat laboratory assessments. We computed within and between subjects coefficient of variation (CV) as well as two way intra-class correlation (ICC) to assess consistency.

### Longitudinal Cohorts

#### Cohort descriptions

We then sought to validate the prognostic relevance of the CBC across multiple well-characterized longitudinal cohort studies: Coronary Artery Risk Development in Young Adults (CARDIA), Chronic Renal Insufficiency Cohort (CRIC), Framingham Heart Study (FHS) Offspring (OS) and 3rd Generation cohorts (3G), and the Multi-Ethnic Study of Atherosclerosis (MESA). Data for CARDIA, FHS OS, FHS 3G, and MESA were obtained from NHLBI BioLINCC (https://biolincc.nhlbi.nih.gov/home/) while data for CRIC were obtained from the NIDDK Central Repository (https://repository.niddk.nih.gov/home/). Participants in each study provided informed consent and each study was approved by the local IRBs at each recruitment site. This secondary analysis was approved by the University of Michigan Institutional Review Board. Further details on cohorts are provided in the Supplemental methods.

#### Outcomes analyses

Within each cohort, we computed PCA scores for the CBC with differential variables using weights from the varimax-rotated PCA performed in NHANES. We then related the predictors (individual underlying CBC and differential variables and the PCA scores) to each outcome [incident myocardial infarction [MI], incident stroke, incident coronary revascularization, MACE3 (composite of mortality, incident MI, and incident stroke), and MACE4 (composite of mortality, incident MI, incident stroke, and incident coronary revascularization), and all-cause mortality] using Cox proportional hazards regression. We performed unadjusted, adjusted for age, sex, and race, and adjusted for clinical covariates (age, sex, race, smoking, body mass index, systolic and diastolic blood pressure, use of antihypertensive medications, diabetes status, total cholesterol, HDL, LDL, and triglycerides) regression analyses. Further details of exclusions in each cohort are provided in the Supplemental methods section.

#### Meta-analyses

We combined standardized hazard ratios (standardized to NHANES mean and variance) using random effects meta-analyses. We performed leave one out meta-analyses to evaluate impact of individual cohorts on estimates.

#### Statistical analyses

All statistical analyses were performed in R 4.4.1 (R Foundation for Statistical Research, Vienna Austria). Key modules included: metafor (for meta-analyses), survival (for survival analyses), `survey` (for survey weighted analyses), `srvyr` (for survey weighted analyses), and `irr` (for intra-class correlation). Modules were obtained from the Comprehensive R Archive Network (CRAN).

## Results

### CBC, traditional risk factors, and all-cause mortality

Because traditional risk factors can influence quantitative biomarker assessments, we first sought to determine the variance in standard laboratory studies that traditional risk factors can explain. We leveraged data from the 2003-2010 cycles of NHANES. 7,843 subjects had complete data and were included in this analysis. The average age was 49, and 52% were female (**Supplemental Table 1**). We used principal component analyses to partition the total variance across all laboratory values, including individual components of the CBC. Eight principal components (PC) explained 63% of the total variance across all common laboratory studies. Interestingly, many of the most highly weighted features were related to the CBC, including the lymphocyte, neutrophil (PMN), and monocyte counts (**Supplemental Figure 1**). Because of the importance and strong weighting of the CBC components with all common laboratory studies, we sought to define the influence of traditional CVD risk factors on the values of all individual laboratory parameters (**Figure 1A**). With respect to total cholesterol and LDL-C, less than 10% of the total variance is explained by traditional CVD risk factors. In contrast, traditional CVD risk factors can account for over 30% of the variance in the values of creatinine Cr and Hb. Conversely, most components of the CBC are more like LDL-C (with low variance explained by traditional risk factors) than CRP (which has a moderate amount of variance that traditional risk factors can explain). Importantly, the CBC itself contained a large amount of independent information relative to the other laboratory studies. Building on these observations, we next examined the weighted association between all-cause mortality and each laboratory study using incremental chi-squared (**Figure 1B**). Intriguingly, we found that the CBC and CBC components have the strongest contribution to all-cause mortality when compared to the lipid panel, HbA1c, CRP, and basic metabolic panel (lipid panel Δχ²=27.0, p = 0.0008; HbA1c Δχ²=6.0, p=0.01; CRP Δχ²=64.8, p<0.0001; BMP Δχ²=88.8, p<0.0001; CBC Δχ²=215.8, p<0.0001; CBC with differential Δχ²=258.9, p<0.0001). Because the individual components of the CBC had distinct and complementary information, we performed PCA on the CBC components to reduce the dimensions. Approximately 63% of the variance in CBC components can be captured by 4 PCs (**Figure 2A**). Interestingly, the four major CBC PCs could be broadly categorized as 1) PMN/lymphocytes, 2) RBC quantity, 3) other inflammatory cells, and 4) RBC properties. Additionally, the CBC and its individual components had favorable stability and intra– and intersubject variability compared with other laboratory studies, considerations important when considering the clinical utility of any quantitative biomarker (**Supplemental Figure 2**).

**Figure 1.**
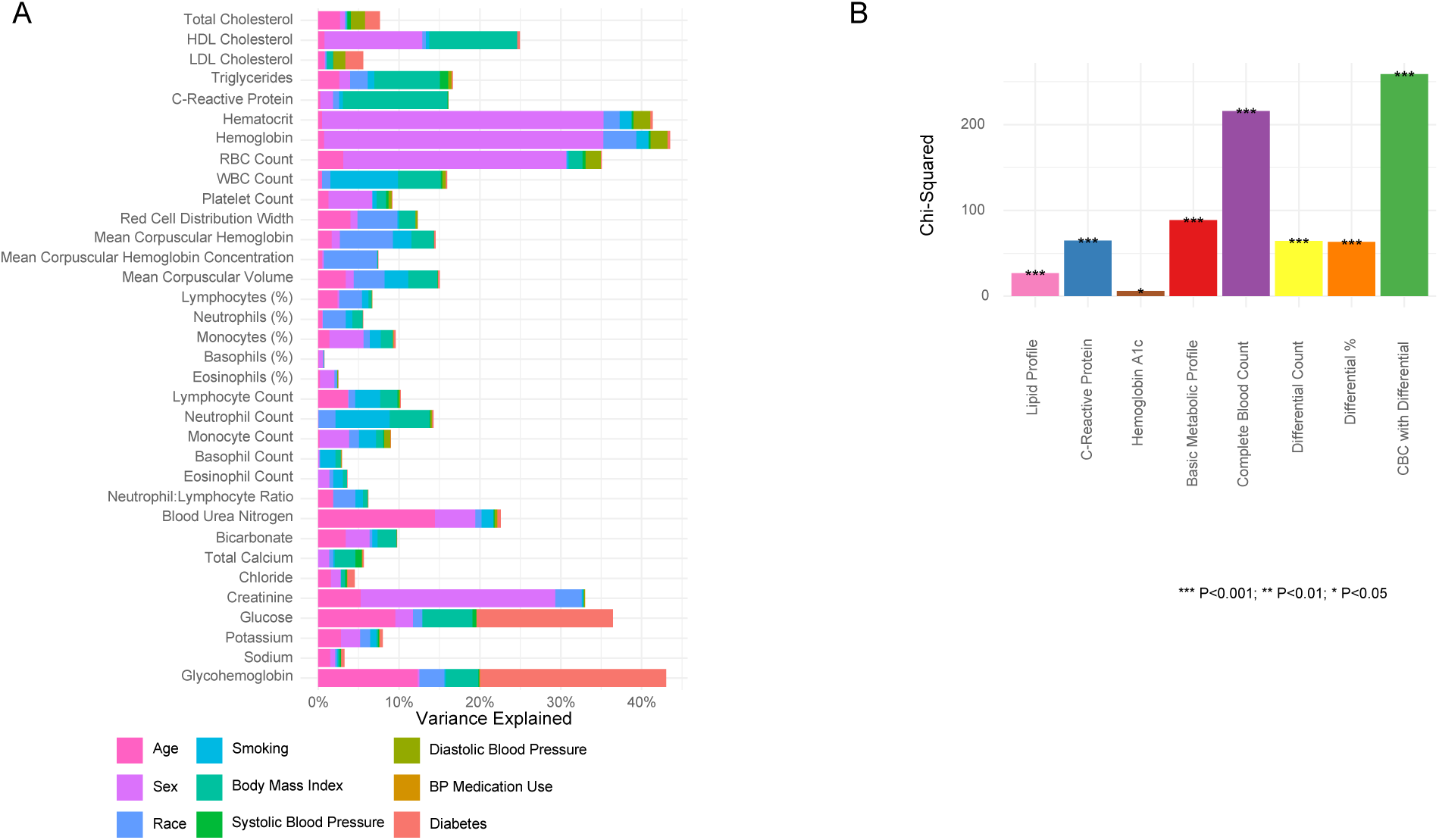
The variance explained of common laboratory studies by traditional risk factors and information content for all-cause mortality. A) We leveraged data from the 2003-2010 cycles of NHANES to assess the variance explained of common laboratory measures by traditional risk factors using 6,162 subjects with complete data and fasting status. Type I analysis of variance accounting for survey weights was performed to estimate percent variance explained after hyperbolic arcsine transformation. These results illustrate that values from the CBC had comparable independent information content beyond traditional risk factors and comparable to that of lipid and basic metabolic panels. B) Using incremental chi-squared, we examined the survey-weighted association between common laboratory panels and all-cause mortality. Adjustments were made for age, sex, race, smoking, BMI, blood pressure, use of antihypertensive medications, and self-reported diabetes. Higher ꭓ^2^ indicates greater information content.

**Figure 2.**
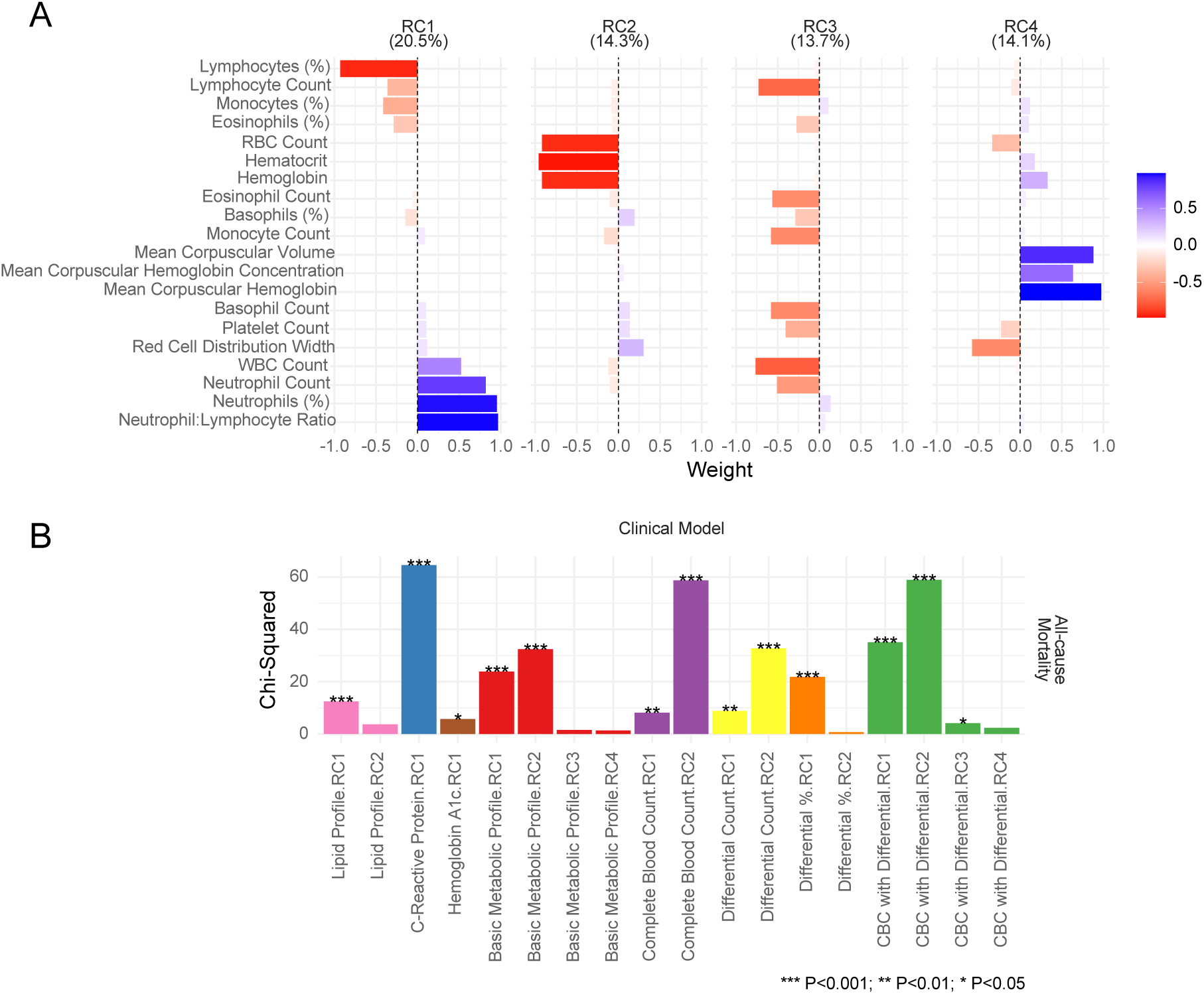
Dimension reduction of the CBC and information content for all-cause mortality. A) Principal component analysis (PCA) of the CBC with differential demonstrated that four components explain 62.9% of the total variance in the CBC with differential study. PCA was performed with varimax post-rotation and accounted for survey sampling weights for the fasting subpopulation in NHANES. The first component (explaining 20.5% of the total variance) was positively loaded on total WBC count, neutrophils, and NLR and negatively loaded on lymphocytes. The second component (explaining for 14.3% of the total variance) was loaded on red blood cell quantity measures, negatively loaded on RBC count, hematocrit, and hemoglobin. The third component (explaining 13.7% of total variance) was negatively loaded with multiple inflammatory cell parameters including total WBC count, lymphocytes, and eosinophils. Finally, the fourth component (explaining 14.1% of total variance) was loaded on red blood cell characteristics with positive loadings for MCH, MCHC, and MCV and negative loading for RDW. B) Analogously to Figure 2, we computed incremental ꭓ^2^ for Cox proportional hazards models for all-cause mortality, adding rotated principal components of common laboratory panels to clinical variables (age, sex, race, smoking, body mass index, systolic and diastolic blood pressure, use of antihypertensive medications, self-reported diabetes status). Higher ꭓ^2^ indicates greater information content. Models accounted for survey sampling weights for the fasting subpopulation in NHANES.

### CBC features are independently associated with refined CVD phenotypes in diverse adults

Building on our cross-sectional NHANES analyses, we wished to validate the all-cause mortality findings and extend our analyses to more diverse cohorts with longitudinal follow-up leveraging more refined CVD endpoints, including incident including myocardial infarction (MI), stroke, and coronary revascularization. We examined 15,527 subjects in the FHS offspring, FHS third generation, MESA, CARDIA, and CRIC cohorts. The average age in this longitudinal analysis was diverse, encompassing young (CARDIA; 25 years) and older adults (FHS OS; 69 years). The baseline characteristics for individuals from each cohort are shown in **Table 1**. Over a total of 249,098 follow-up person-years, there were 697 incident MI, 458 incident strokes, 234 incident coronary revascularizations, and 2,459 deaths. We used two broad approaches to quantify the associations between the CBC and these CVD endpoints. First, we quantified CVD risk across the spectrum of individual CBC components, and second, we quantified CVD risk based on the four CBC principal component scores that we derived from the cross-sectional NHANES analysis (**Supplemental data file** and **Figure 3**). In the fully adjusted model, several observations are worth noting. All CBC components except basophil, eosinophil, and platelet counts were associated with all-cause mortality. WBC, PMN, lymphocyte, and monocyte counts were associated with the CVD composite endpoint (MACE3). Additionally, even after adjusting for hs-CRP (a marker of systemic inflammation), the CBC components remained significantly related to all-cause mortality and several CVD endpoints (**Supplemental Figure 3**). Building on the PCA of CBC components in NHANES, we applied the PC dimensions to the cohorts. We examined their association with more refined CVD endpoints and all-cause mortality in FHS offspring, FHS 3G, MESA, CARDIA, and CRIC cohorts (n=15,527). Numerous strong associations were evident with the refined CVD endpoints and all-cause mortality with the CBC PC dimensions (**Supplemental Figure 4**). Of note, the PMN/lymphocyte (hazard ratio 2.45 [1.42-4.24]; p=0.001) and RBC quantity (hazard ratio 1.86 [1.34-2.59]; p=0.0002) PC scores were strongly associated with all-cause mortality in each cohort and when all cohorts were combined (**Figure 4**).

**Figure 3.**
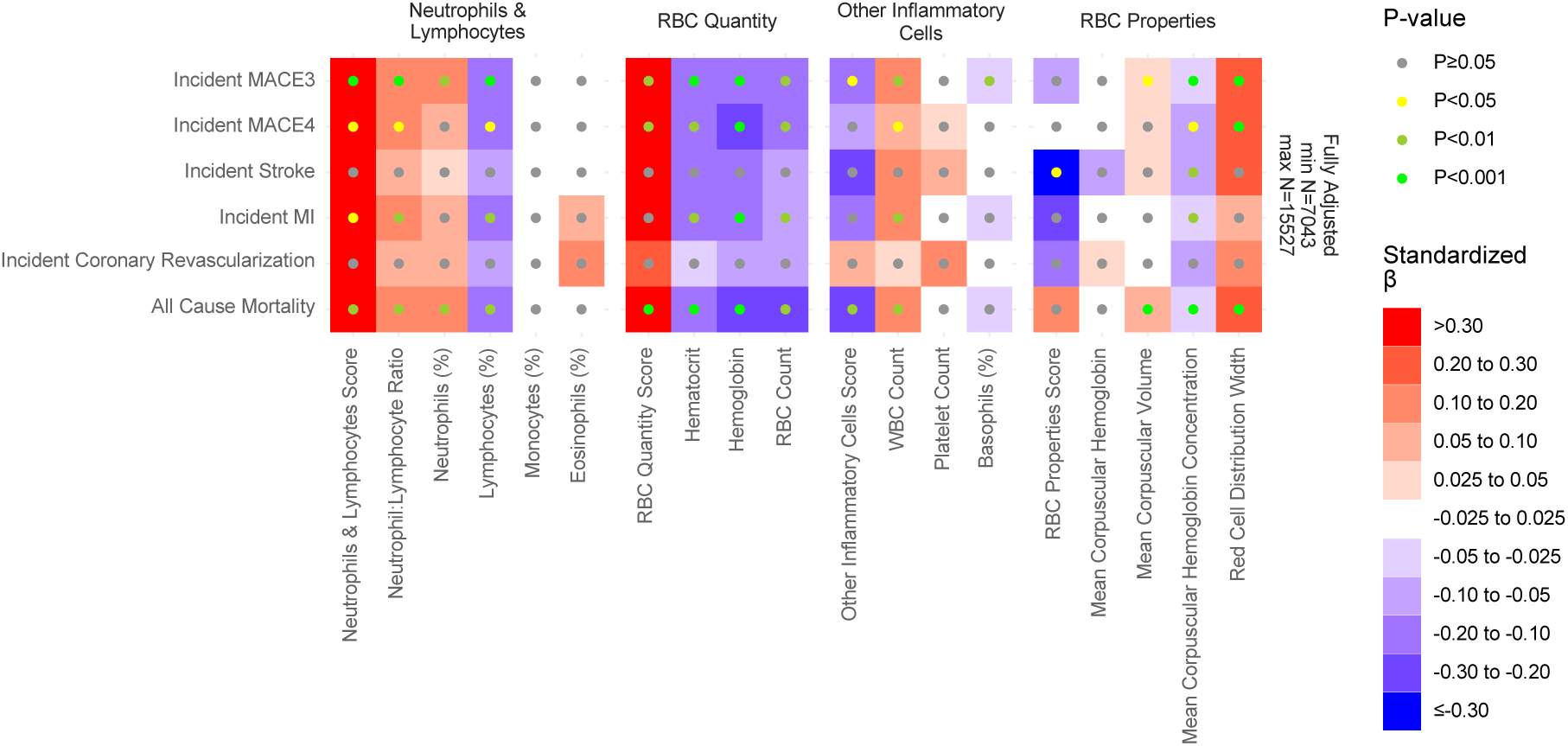
CBC components are associated with refined CV endpoints in five diverse, longitudinal cohorts. Heatmap of standardized ꞵ-coefficients from Cox proportional hazards models relating CBC components to CV endpoints. Values of standardized ꞵ-coefficients directionality (color), magnitude (color gradient), and statistical significance (color of circle) are depicted. All models are fully adjusted for age, sex, race, smoking, body mass index, systolic and diastolic blood pressure, use of antihypertensive medications, diabetes status, total cholesterol, HDL, LDL, and triglycerides.

**Figure 4.**
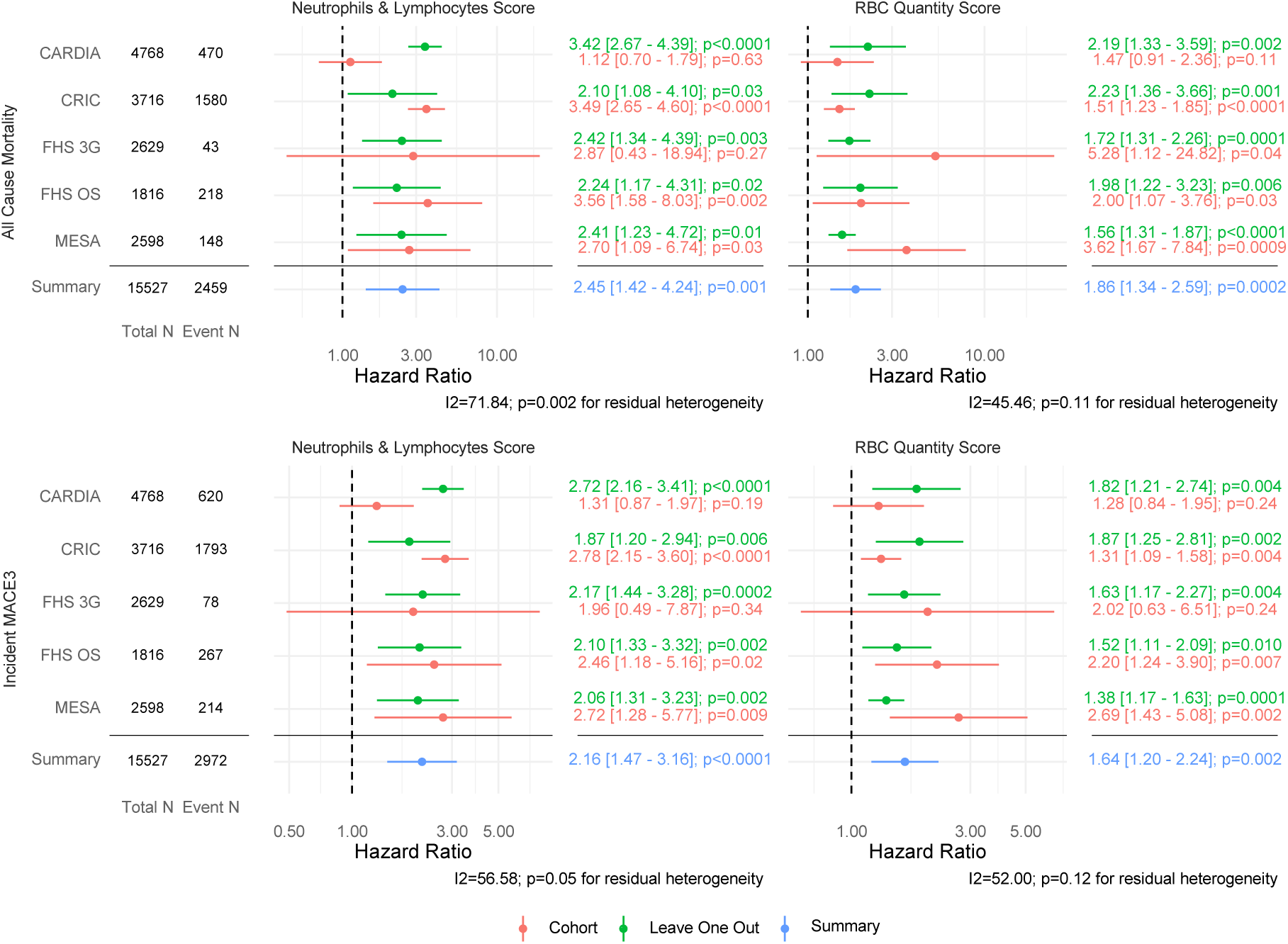
Key CBC-derived scores for all-cause mortality and MACE3 in five longitudinal cohorts. Forest plots of standardized β-coefficients of key CBC-derived scores for all-cause mortality and MACE3. Tables on the left indicate the number of total subjects and events in each cohort. Hazard ratios (circles) and their 95% confidence intervals (bars) are indicated in different colors for individual cohort estimates (coral), summary meta-analysis (blue), and leave one cohort analysis sensitivity analyses (green). Cox models in each cohort were adjusted for age, sex, race, smoking, body mass index, systolic and diastolic blood pressure, use of antihypertensive medications, diabetes status, total cholesterol, HDL, LDL, and triglycerides.

**Table 1.** Participant characteristics of longitudinal cohorts.

| Characteristic | FHS OS<br>N = 1,816<sup>1</sup> | FHS 3G<br>N = 2,629<sup>1</sup> | MESA<br>N=2,598<sup>1</sup> | CARDIA<br>N = 4,768<sup>1</sup> | CRIC<br>N = 3,716<sup>1</sup> |
| --- | --- | --- | --- | --- | --- |
| Age | 69 (64, 75) | 48 (41, 54) | 68 (61, 76) | 25.0 (22.0, 28.0) | 59 (52, 66) |
| Female Sex | 1,062 (58%) | 1,379 (52%) | 1,406 (54%) | 2,607 (55%) | 1,669 (45%) |
| Race |  |  |  |  |  |
| White | 1,731 (95%) | 2,358 (90%) | 1,118 (43%) | 2,329 (49%) | 1,736 (47%) |
| Non-White | 15 (0.8%) | 271 (10%) |  |  |  |
| Black |  |  | 724 (28%) | 2,439 (51%) | 1,489 (40%) |
| Chinese-American |  |  | 24 (0.9%) |  |  |
| Hispanic |  |  | 732 (28%) |  |  |
| American Indian/Alaska native |  |  |  |  | 15 (0.4%) |
| Asian |  |  |  |  | 105 (2.8%) |
| More than one race |  |  |  |  | 78 (2.1%) |
| Native Hawaiian/Pacific Islands |  |  |  |  | 6 (0.2%) |
| Unknown | 70 (3.9%) |  |  |  | 287 (7.7%) |
| Smoking |  |  |  |  |  |
| current | 111 (6.1%) | 243 (9.2%) | 211 (8.1%) | 1,461 (31%) | 488 (13%) |
| former | 984 (54%) | 819 (31%) | 1095 (42%) | 638 (13%) | 1,542 (41%) |
| never | 721 (40%) | 1,567 (60%) | 1,275 (49%) | 2,669 (56%) | 1,686 (45%) |
| Body Mass Index | 27.7 (24.7, 31.1) | 27.1 (24.1, 31.1) | 28.5 (25.2, 32.4) | 23.5 (21.1, 26.4) | 31 (27, 36) |
| Total Cholesterol | 187 (163, 210) | 184 (163, 207) | 181 (157, 206) | 173 (153, 197) | 178 (152, 208) |
| HDL Cholesterol | 61 (49, 75) | 57 (47, 71) | 53 (44, 64) | 52 (44, 61) | 45 (37, 54) |
| LDL Cholesterol | 99 (82, 121) | 102 (83, 123) | 104 (83, 126) | 106 (87, 127) | 98 (78, 123) |
| Triglycerides | 99 (75, 137) | 94 (68, 134) | 93 (70, 130) | 62 (45, 84) | 128 (89, 185) |
| Systolic Blood Pressure | 126 (115, 137) | 116 (107, 126) | 120 (110, 136) | 110 (103, 117) | 125 (113, 141) |
| Diastolic Blood Pressure | 72 (66, 79) | 75 (68, 81) | 68 (61, 75) | 68 (62, 75) | 71 (62, 80) |
| BP Medication Use | 949 (52%) | 488 (19%) | 1,433 (55%) | 107 (2.2%) | 3,608 (97%) |
| Diabetes |  |  |  |  |  |
| Diabetic | 246 (14%) | 128 (4.9%) | 511 (20%) | 40 (0.8%) | 1,800 (48%) |
| Non-Diabetic | 1,570 (86%) | 2,501 (95%) | 2,087 (80%) |  | 1,916 (52%) |
| High Sensitivity C-Reactive Protein | 1.54 (0.77, 3.21) | 1.27 (0.61, 2.95) | 2.1 (0.9, 4.4) |  | 3 (1, 6) |
| Hematocrit | 40.0 (37.7, 42.5) | 40.8 (38.4, 43.7) | 40.0 (37.5, 42.7) | 42.0 (39.0, 45.5) | 37.4 (34.2, 40.9) |
| Hemoglobin | 13.70 (12.90, 14.50) | 14.00 (13.10, 15.00) | 13.40 (12.40, 14.40) | 14.20 (13.20, 15.30) | 12.55 (11.40, 13.80) |
| RBC Count | 4.33 (4.07, 4.63) | 4.51 (4.23, 4.84) | 4.48 (4.15, 4.79) | 4.78 (4.41, 5.17) | 4.26 (3.86, 4.66) |
| WBC Count | 5.75 (4.80, 6.90) | 5.80 (4.90, 6.80) | 5.80 (4.80, 6.90) | 5.80 (4.80, 7.10) | 6.30 (5.20, 7.70) |
| Mean Corpuscular Hemoglobin | 31.60 (30.50, 32.50) | 31.10 (30.10, 32.00) | 30.20 (28.90, 31.40) | 29.93 (28.66, 31.07) | 30.00 (28.40, 31.00) |
| Mean Corpuscular Hemoglobin Concentration | 34.20 (33.60, 34.70) | 34.20 (33.70, 34.60) | 33.40 (32.80, 34.10) | 33.76 (33.18, 34.30) | 33.70 (33.00, 34.20) |
| Mean Corpuscular Volume | 92.5 (89.8, 95.2) | 90.8 (88.2, 93.5) | 90.0 (87.0, 93.0) |  | 88.8 (85.0, 92.0) |
| Red Cell Distribution Width | 13.00 (12.60, 13.50) | 12.70 (12.40, 13.20) | 13.90 (13.40, 14.60) |  |  |
| Platelet Count | 229 (197, | 241 (207, | 224 (187, | 267 (227, | 237 (195, |

|  | 268) | 278) | 266) | 313) | 284) |
| --- | --- | --- | --- | --- | --- |
| Platelet Volume | 8.80 (8.20, 9.40) | 8.40 (7.90, 9.00) |  |  |  |
| Lymphocytes (%) | 26 (21, 31) | 28 (24, 33) | 30 (24, 36) | 37 (30, 43) | 27 (21, 33) |
| Neutrophils (%) | 61 (55, 66) | 59 (54, 64) | 59 (53, 65) | 53 (46, 60) | 62 (55, 68) |
| Monocytes (%) | 8.80 (7.50, 10.20) | 8.40 (7.20, 9.80) | 8.00 (6.00, 9.00) | 5.00 (4.00, 7.00) | 7.30 (6.00, 8.94) |
| Basophils (%) | 0.60 (0.40, 0.80) | 0.80 (0.60, 1.00) | 1.00 (0.00, 1.00) | 0.00 (0.00, 1.00) | 0.40 (0.00, 0.90) |
| Eosinophils (%) | 2.90 (2.00, 4.20) | 2.60 (1.80, 3.90) | 2.00 (1.00, 4.00) | 2.00 (1.00, 3.00) | 2.78 (1.72, 4.12) |
| Lymphocyte Count | 1.50 (1.20, 1.80) | 1.60 (1.30, 1.90) | 1.70 (1.35, 2.10) | 2.10 (1.73, 2.54) | 1.65 (1.30, 2.09) |
| Neutrophil Count | 3.50 (2.80, 4.30) | 3.40 (2.70, 4.20) | 3.40 (2.70, 4.30) | 3.02 (2.30, 4.02) | 3.83 (2.95, 4.96) |
| Monocyte Count | 0.50 (0.40, 0.60) | 0.50 (0.40, 0.60) | 0.40 (0.31, 0.57) | 0.30 (0.20, 0.41) | 0.46 (0.36, 0.60) |
| Basophil Count | 0.00 (0.00, 0.00) | 0.00 (0.00, 0.10) | 0.00 (0.00, 0.05) | 0.00 (0.00, 0.06) | 0.03 (0.00, 0.05) |
| Eosinophil Count | 0.20 (0.10, 0.30) | 0.20 (0.10, 0.20) | 0.10 (0.10, 0.20) | 0.12 (0.06, 0.20) | 0.18 (0.10, 0.27) |
| Neutrophil:Lymphocyte Ratio | 2.35 (1.77, 3.10) | 2.10 (1.65, 2.64) | 1.97 (1.46, 2.69) | 1.45 (1.07, 1.97) | 2.32 (1.73, 3.18) |
| Incident MACE3 | 267 (15%) | 78 (3.0%) | 214 (8.2%) | 620 (13%) | 1,793 (48%) |
| Incident MACE4 | 285 (16%) | 99 (3.8%) | 239 (9.2%) | 651 (14%) |  |
| Incident MI | 33 (1.8%) | 24 (0.9%) | 41 (1.6%) | 104 (2.2%) | 495 (13%) |
| Incident Stroke | 43 (2.4%) | 14 (0.5%) | 52 (2.0%) | 109 (2.3%) | 240 (6.5%) |
| Incident Coronary Revascularization | 42 (2.3%) | 40 (1.5%) | 46 (1.8%) | 106 (2.2%) |  |
| All Cause Mortality | 218 (12%) | 43 (1.6%) | 148 (5.7%) | 470 (9.9%) | 1,580 (43%) |

1 1 Median (Q1, Q3); n (%)

## Discussion

In this study of 23,370 diverse adults across six cohorts, we defined the relationships between the CBC, traditional CV risk factors, CVD endpoints, and all-cause mortality. Specifically, we found that: 1) traditional CVD risk factors explain less than 10% of the variance in the CBC; 2) individual CBC components were associated with all-cause mortality in all six cohorts examined; and 3) individual CBC components were strongly associated with MI, revascularization, and stroke and even after adjustment for traditional risk factors and standard laboratory values including CRP. To our knowledge, this is the first study to systematically examine the association of the CBC and its components to traditional risk factors and other common laboratory studies with CVD endpoints.

The associations between CC and CVD were first described in a landmark case-control study in patients with MI.^17^ Subsequently, several cohort studies have validated and built on findings from this seminal study.^18,19^ For example, a large UK study from the CALIBER cohort demonstrated that high PMN counts and low lymphocyte counts had strong associations with myocardial infarction, peripheral arterial disease, and heart failure.^20^ With respect to the effects of CV therapies on inflammation and WBC parameters, a recent analysis of five randomized control trials demonstrated that the NLR was associated with CV events and all-cause mortality.^21^ Intriguingly, this analysis found that specific CV therapies, including lipid-lowering and anti-inflammatory therapies, had distinct effects on the NLR, reinforcing not only that specific circulating immune cell populations are associated with CVD but also that anti-inflammatory therapeutics have distinct effects on CC populations and the immune system in general. In addition to the immune components of the CBC, RBC features have also been associated with CVD.^22^ Sabatine and colleagues found that low values of RBC features were an independent predictor of MACE in acute coronary syndromes.^11^ Studies have typically focused on anemia and elevated WBC counts; however, we found that even within normal ranges, WBC and RBC features are associated with CVD endpoints (**Supplemental Figure 5**), results that complement the recent observations by Foy and colleagues.^23^ Our study builds on these prior studies, validating CBC associations with CVD even after correction for traditional ASCVD risk factors and inflammatory biomarkers.

The quantitative risk assessment of CVD is the cornerstone of both primary and secondary prevention. It informs patient-centered diagnostic and therapeutic approaches. The current quantitative risk approaches combine patient demographic characteristics (including age and sex), biological variables (blood pressure and smoking), and laboratory studies (lipid and diabetes parameters). Our findings add to the discourse on CVD risk assessment, suggesting that readily available laboratory information from the CBC could complement existing components of quantitative CVD risk assessments.

### Strengths and Limitations

Several limitations of our study warrant consideration. First, the observational nature of our study precludes any strict determinations of causality with respect to CC pathobiology; however, we believe that our findings are hypothesis generating and can help inform future mechanistic and translational studies and underscore the importance of further characterization of CC contributions to ASCVD. Second, despite the consistency of our findings across cohorts, there is heterogeneity in the characteristics of each cohort, making comparisons imperfect. However, this heterogeneity can also be viewed as a strength, because the main conclusions are consistent across diverse cohorts, strengthens them, and makes possible more generalizations. Importantly, the triangulation from complementary approaches yielded consistent results, largely strengthening the robustness and validity of the findings.

## Conclusion

In summary, these contemporary analyses of 23,370 subjects in six cohorts systematically relate the CBC to traditional risk factors, CVD endpoints, and all-cause mortality. Our findings highlight the potential of incorporating the CBC into CVD risk assessment and suggest novel mechanistic insights connecting the components of the CBC to the pathobiology of CVD. This study adds further evidence linking CBC components to all dimensions of CVD and advances our understanding of how circulating cells can be used to identify individuals at risk of CVD.

## Author Contributions

SNG and VLM contributed equally, and both designed the research, analyzed the data, and wrote the paper.

## Data and Code Availability

Data for NHANES are freely available from the National Center for Health Statistics Data for CARDIA, FHS OS, FHS 3G and MESA were obtained from NHLBI BIoLINCC and can be obtained by qualified investigators from NHLBI with an approved research proposal, data use agreement and IRB approval.

Data for CRIC were obtained from the NIDDK Central Repository and can be obtained by qualified investigators from NHLBI with an approved research proposal, data use agreement and IRB approval.

No new software algorithms or tools were developed for this project.

## Data Availability

All data produced in the present study are available upon reasonable request to the authors.

## Abbreviations

ASCVD: atherosclerotic cardiovascular disease
BMP: basic metabolic panel
CAC: coronary artery calcification
CARDIA: Coronary Artery Risk Development in Young Adults
CBC: Complete blood count
CC: circulating cells
CM: circulating mediators
CVD: cardiovascular disease
CRIC: chronic renal insufficiency cohort
CRP: C-reactive protein
HbA1c: hemoglobin A1c
HDL: high-density lipoprotein
Hb: hemoglobin
LDL: low-density lipoprotein
MESA: Multi-Ethnic Study of Atherosclerosis
MI: myocardial infarction
NHANES: National Health and Nutrition Examination Survey
PCA: principal component analysis
PMN: neutrophil
RBC: red blood cell
WBC: white blood cell

## Acknowledgements

Although not directly involved in the conduct of this study, we thank the research volunteers and staff of the underlying studies for their contributions and commitment to research.

