## Supplemental materials for "The complete blood count and cardiovascular disease: analyses across six cohorts of 23,370 adults" (from another version of this preprint: version 1, 2024-10-18)

**Supplemental methods and data.**

**CARDIA**

The Coronary Artery Risk Development in Young Adults (CARDIA) is an NIH-sponsored longitudinal cohort study of young adults that was initiated in 1983 across four field centers (Birmingham, AL; Chicago, IL; Minneapolis, MN; Oakland, CA) which initially recruited 5,115 subjects. We excluded subjects who did not complete the baseline exam (N=2), were missing key covariates (N=103), were missing CBC (N=37) or differential data (N=2), had a total of differential percentages <95% or >105% (N=202), or had missing or invalid follow-up data (N=1).

**CRIC**

The Chronic Renal Insufficiency Cohort (CRIC) is an NIH-sponsored longitudinal cohort study designed to evaluate the progression of chronic renal insufficiency and cardiovascular disease initiated in 2003 across eight cities (Albuquerque, NM; Ann Arbor, MI; Baltimore, MD; Chicago, IL; Cleveland, OH; New Orleans, LA; Oakland, CA; Philadelphia, PA). We considered the earliest available CBC measure as the baseline of 5,625 subjects with data available through the NIDDK Central Repository. We excluded subjects with missing covariates at the baseline exam (N=1717), incomplete CBC variables (N=28) or differential (N=31), had a total of differential percentages <95% or >105% (N=108), or had missing or invalid follow-up data or experienced MACE before the CBC collection (N=25).

**FHS OS**

Framingham Heart Study (FHS) Offspring (OS) Study is a longitudinal cohort study of the offspring of the participants of the original Framingham Heart Study and their spouses, which was begun in 1971. CBC was measured at Exam 9 (conducted between 4/2011 and 3/2014). Of 5,478 subjects with data available through BioLINCC, we excluded subjects who did not attend the 9th examination (N=3078), had missing covariates (N=175), had incomplete CBC variables (N=91), or had missing or invalid follow-up data or experienced MACE before CBC collection (N=318).

**FHS 3G**

Framingham Heart Study (FHS) Third Generation Study is a longitudinal cohort study of the offspring of the participants of the Framingham Heart Study Offspring Study, which was begun in 2002. CBC was measured at Exam 2 (conducted between 2009 and 2011). Of 4,529 subjects with data available through BioLINCC, we excluded subjects who did not attend the 2nd examination (N=798), had missing covariates (N=75), had incomplete CBC variables (N=952), or had missing or invalid follow-up data or experienced MACE before CBC collection (N=117).

**MESA**

The Multi-Ethnic Study of Atherosclerosis (MESA) is a longitudinal cohort study conducted across 6 centers (Baltimore, MD; Chicago, IL; Los Angeles, CA; Minneapolis, MN; New York, NY; Winston-Salem, NC) which was begun in 2000. CBC was performed at the 5th Exam. Of 6,814 individuals with data available through BioLINCC, we excluded individuals who did not complete the 5th exam (N=2,214), were missing covariates (N=174), had incomplete CBC measures (N=1,618) or differential (N=1), had a total of differential percentages <95% or >105% (N=11), or had missing or invalid follow-up data or experienced MACE before the CBC collection (N=198).

Supplemental Table 1: Characteristics of NHANES Cohort (Without Weighting)

| **Characteristic** | **N = 7,843***^1^* |
| --- | --- |
| cycle |  |
| 2003-2004 | 1,681 (21%) |
| 2005-2006 | 1,679 (21%) |
| 2007-2008 | 2,091 (27%) |
| 2009-2010 | 2,392 (30%) |
| Age | 49 (34, 64) |
| Sex |  |
| female | 4,063 (52%) |
| male | 3,780 (48%) |
| Race |  |
| Mexican American | 1,523 (19%) |
| non-Hispanic Black | 1,426 (18%) |
| non-Hispanic White | 3,927 (50%) |
| other Hispanic | 618 (7.9%) |
| other race | 349 (4.4%) |
| Smoking |  |
| former | 2,041 (26%) |
| never | 4,177 (53%) |
| present | 1,625 (21%) |
| Body Mass Index | 28 (24, 32) |
| Systolic Blood Pressure | 119 (110, 132) |
| Diastolic Blood Pressure | 69 (61, 76) |
| BP Medication Use | 7,435 (95%) |
| Diabetes |  |
| borderline DM | 6,883 (89%) |
| don't know | 5 (<0.1%) |
| no DM | 819 (11%) |
| Unknown | 136 |
| Total Cholesterol | 194 (167, 222) |
| HDL Cholesterol | 52 (43, 64) |
| LDL Cholesterol | 113 (91, 138) |
| Triglycerides | 111 (79, 160) |
| C-Reactive Protein | 0.20 (0.08, 0.48) |
| Glycohemoglobin | 5.50 (5.20, 5.80) |
| Blood Urea Nitrogen | 12.0 (9.0, 15.0) |
| Bicarbonate | 25.00 (24.00, 27.00) |
| Total Calcium | 9.40 (9.20, 9.60) |
| Chloride | 104.00 (102.00, 106.00) |
| Creatinine | 0.84 (0.70, 1.00) |
| Glucose | 93 (87, 103) |
| Potassium | 4.00 (3.80, 4.20) |
| Sodium | 139.00 (138.00, 141.00) |
| Hematocrit | 42.2 (38.9, 45.3) |
| Hemoglobin | 14.40 (13.30, 15.40) |
| RBC Count | 4.70 (4.35, 5.05) |
| WBC Count | 6.50 (5.40, 7.90) |
| Mean Corpuscular Hemoglobin | 30.70 (29.50, 31.80) |
| Mean Corpuscular Hemoglobin Concentration | 34.00 (33.40, 34.60) |
| Mean Corpuscular Volume | 90.0 (86.7, 93.0) |
| Red Cell Distribution Width | 12.60 (12.20, 13.30) |
| Platelet Count | 249 (209, 294) |
| Lymphocyte Count | 1.90 (1.60, 2.30) |
| Neutrophil Count | 3.70 (2.90, 4.80) |
| Monocyte Count | 0.50 (0.40, 0.60) |
| Basophil Count | 0.00 (0.00, 0.10) |
| Eosinophil Count | 0.20 (0.10, 0.30) |
| Neutrophil:Lymphocyte Ratio | 1.95 (1.47, 2.64) |
| Lymphocytes (%) | 30 (24, 36) |
| Neutrophils (%) | 58 (52, 65) |
| Monocytes (%) | 7.80 (6.40, 9.30) |
| Basophils (%) | 0.60 (0.40, 0.90) |
| Eosinophils (%) | 2.50 (1.70, 3.80) |
| All Cause Mortality | 1,343 (17%) |
| Cardiac Mortality | 349 (4.5%) |
| Cardiovascular Mortality | 427 (5.4%) |

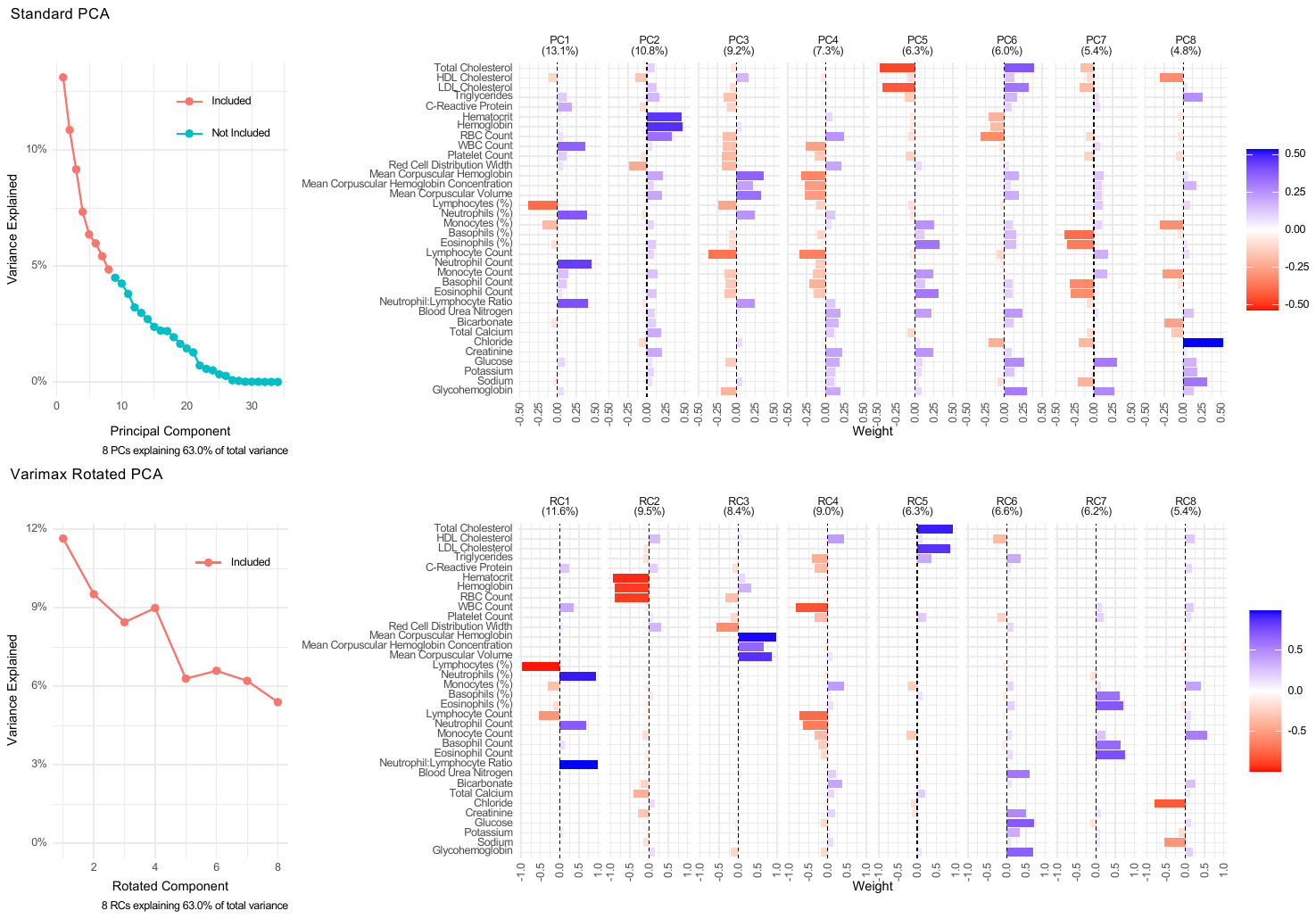

**Supplemental Figure 1. Dimension reduction of all common laboratory studies in NHANES.** Principal component analysis (PCA) of all common laboratory studies demonstrates that the eight main components explain over 60% of the total variance across all laboratory values.  PCA was performed without (top) and with (bottom) varimax post-rotation and accounted for survey sampling weights for the fasting subpopulation in NHANES. Importantly, the CBC components were highly weighted across all laboratory values.   The left panels are scree plots. The right panels are loading plots for eight components. Scree plots indicate variance explained for consecutive components. The loadings plots indicate positive (blue) and negative (red) loadings for individual lab variables in each component. The variance explained by each indicated is shown in parentheses.

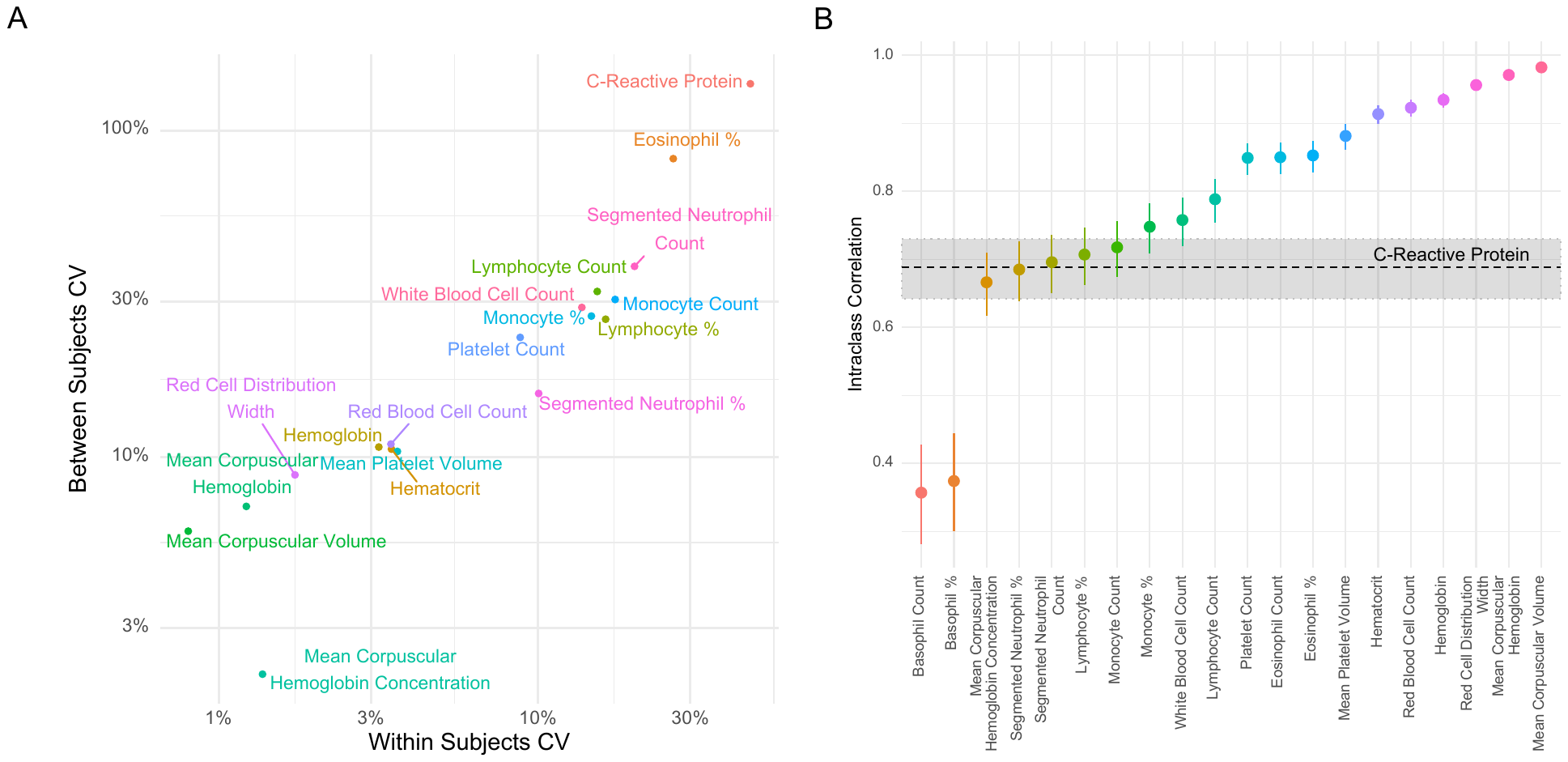

**Supplemental Figure 2. Between and within subjects coefficient of variation for standard laboratory studies and intraclass correlation coefficient for test-retest of CBC in NHANES.** A) Between subjects CV (y-axis) versus within subjects CV (x-axis) for CBC with differential variables demonstrates that repeatability of CBC with differential values are generally comparable or smaller than for C-reactive protein. Survey sampling weights did not apply to the laboratory assay subsample within NHANES. B) Intraclass correlation (ICC) (circles) and 95% confidence intervals (bars) for CBC with differential measures ordered by increasing ICC, indicating higher agreement between test and retest measures. ICC for C-reactive protein is indicated with a dashed horizontal line with the gray band representing its 95% confidence interval. Besides basophils, CBC with differential measures have comparable or higher (better) ICC than C-reactive protein.

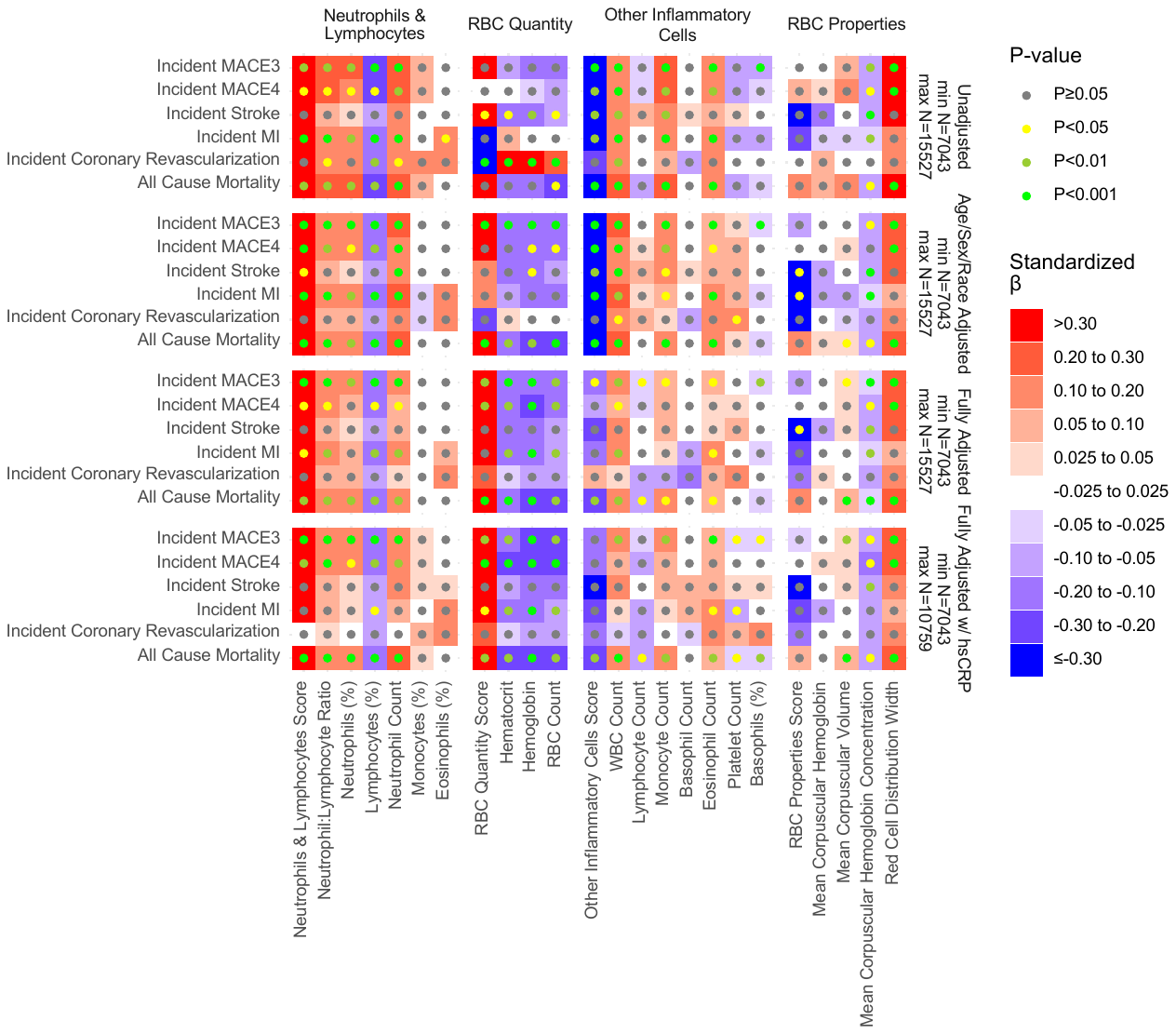

**Supplemental Figure 3. Comprehensive heatmap of standardized ꞵ-coefficients of CBC measures for CV endpoints meta-analyzed across five longitudinal cohorts.** This figure is analogous to Figure 3 with the addition of all CBC with differential measures as well as incorporating unadjusted, age/sex/race adjusted, fully adjusted (age, sex, race, smoking, body mass index, systolic and diastolic blood pressure, use of antihypertensive medications, diabetes status, total cholesterol, HDL, LDL, and triglycerides), and fully adjusted with CRP models. The magnitude of standardized ꞵ-coefficients from Cox proportional hazards models are indicated by the color intensity of heatmap cells with color indicating directionality (red = positive, meaning higher measures are associated with higher risk; blue = negative, meaning lower measures are associated with lower risk). The color of the dots in the center indicates statistical significance.

### Neutrophils & Lymphocytes Score (Unadjusted)

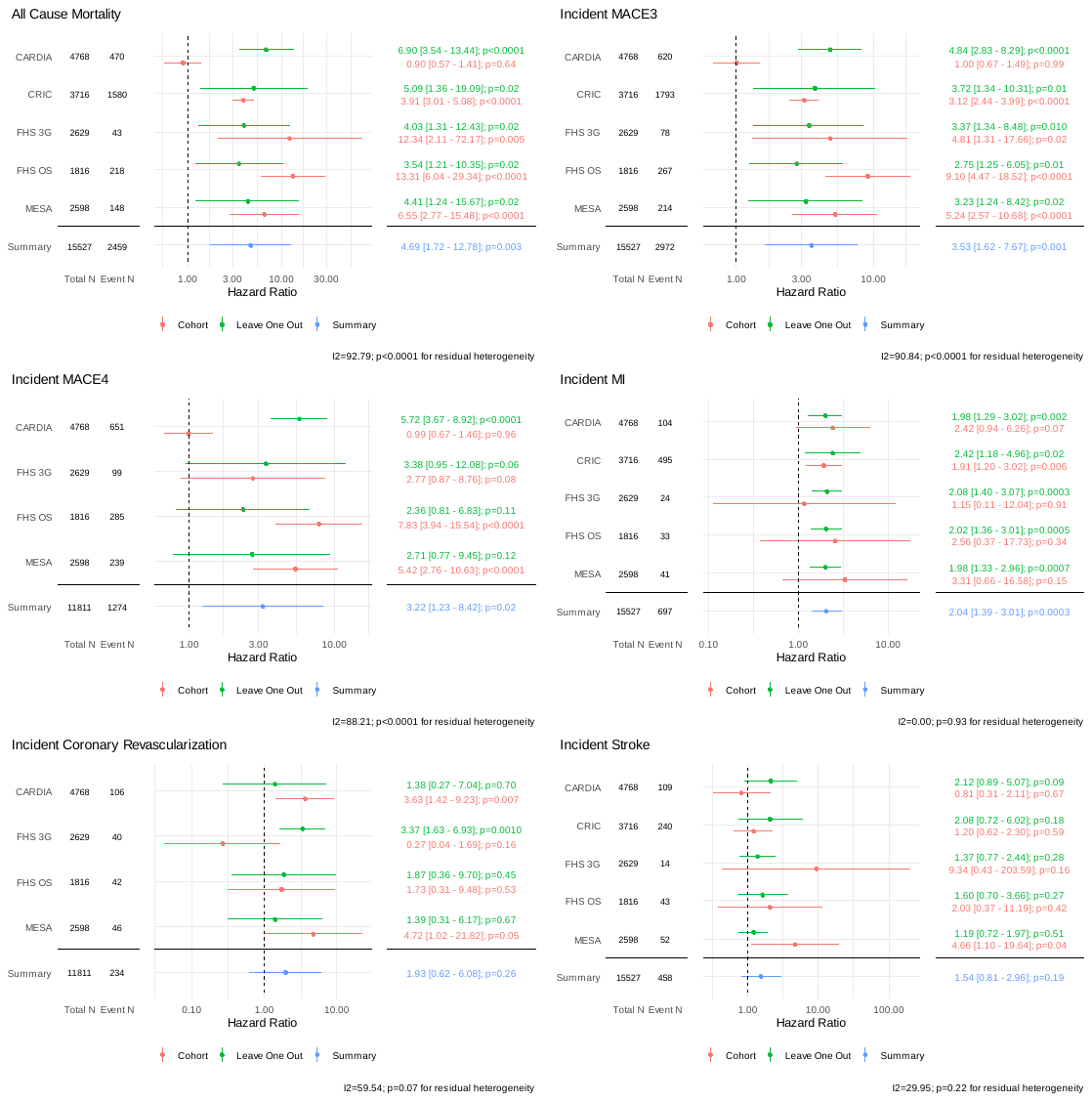

### Neutrophils & Lymphocytes Score (Age/Sex/Race Adjusted)

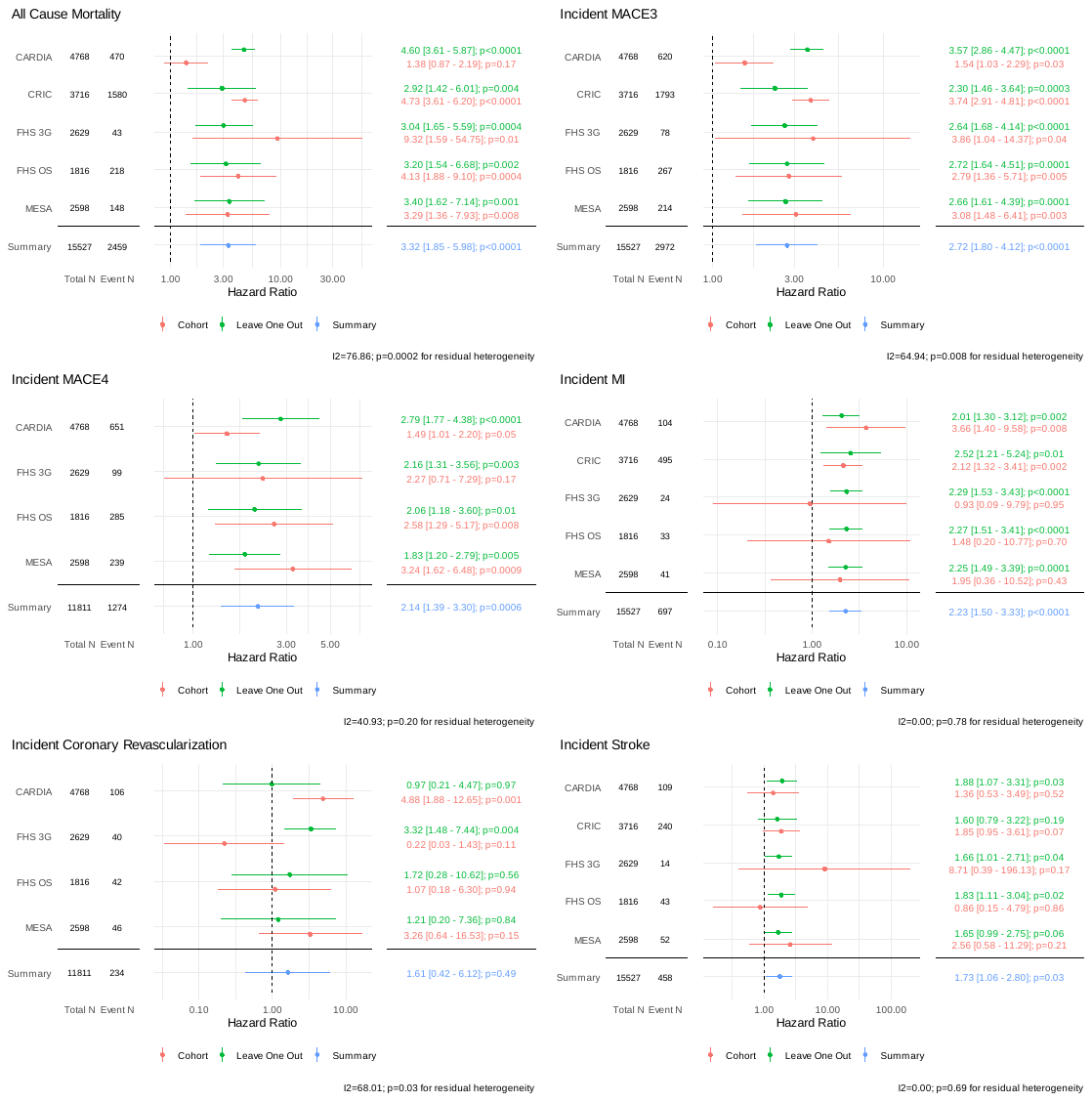

### Neutrophils & Lymphocytes Score (Fully Adjusted)

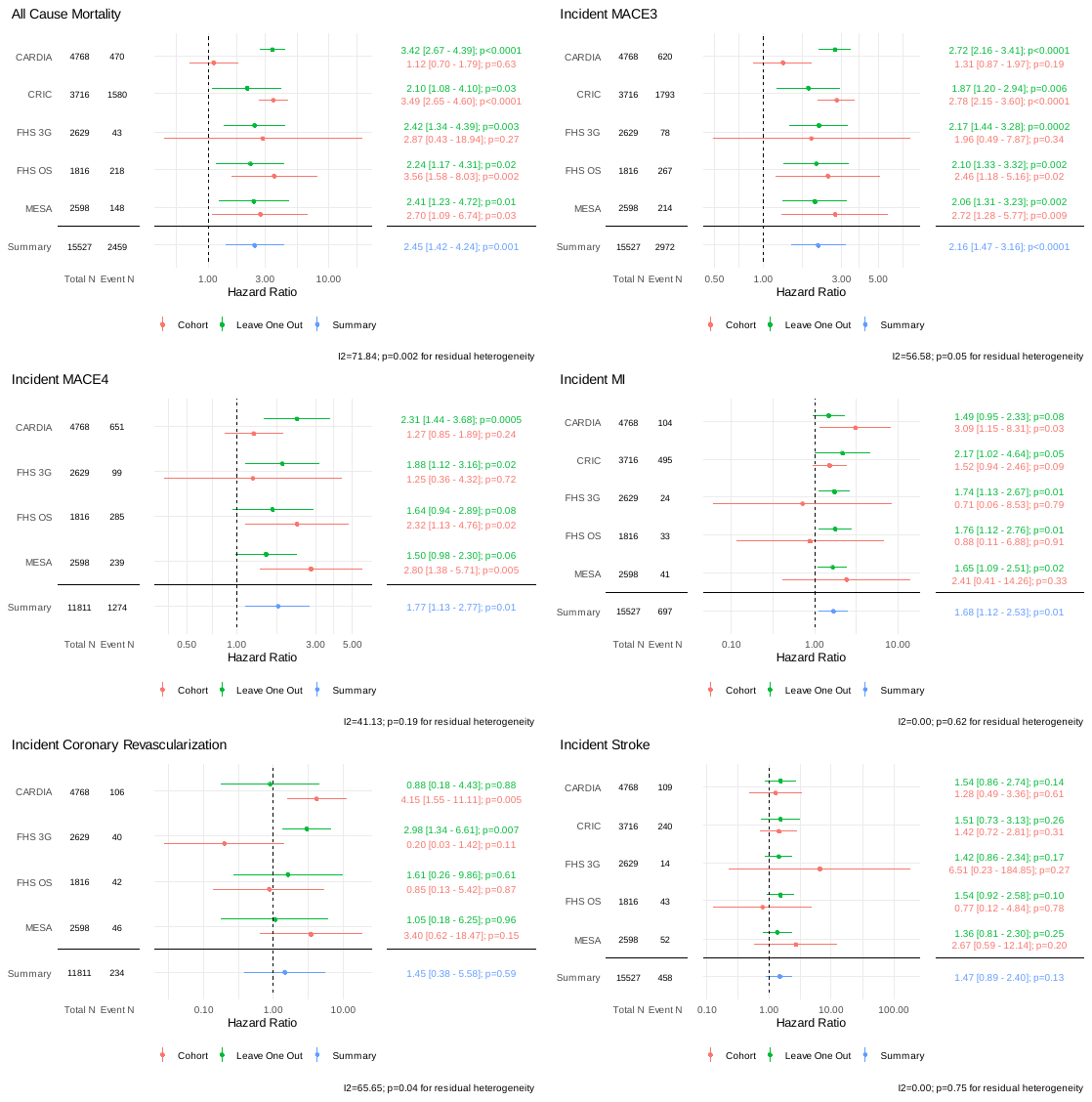

### Neutrophils & Lymphocytes Score (Fully Adjusted w/ hsCRP)

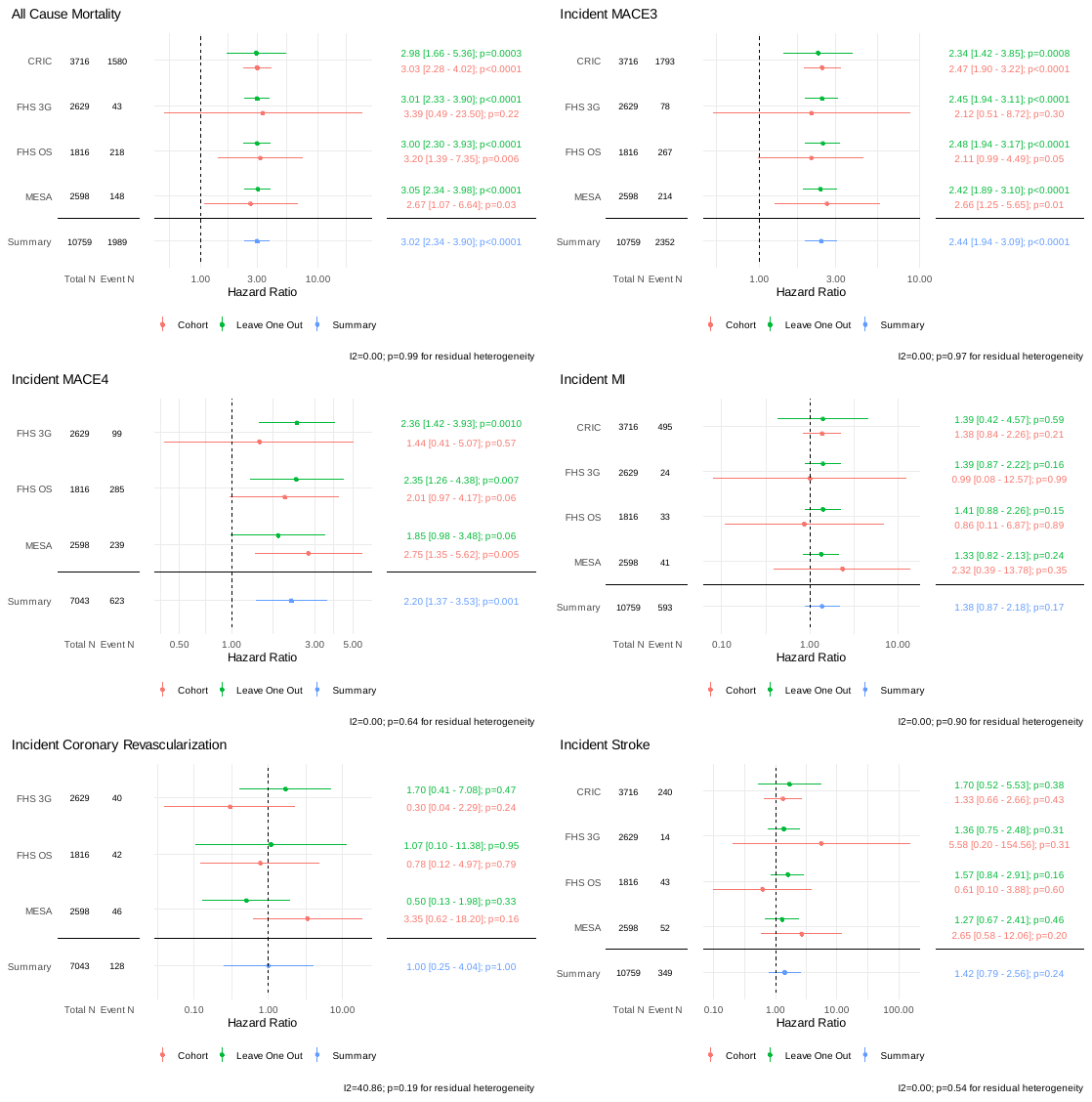

### RBC Quantity Score (Unadjusted)

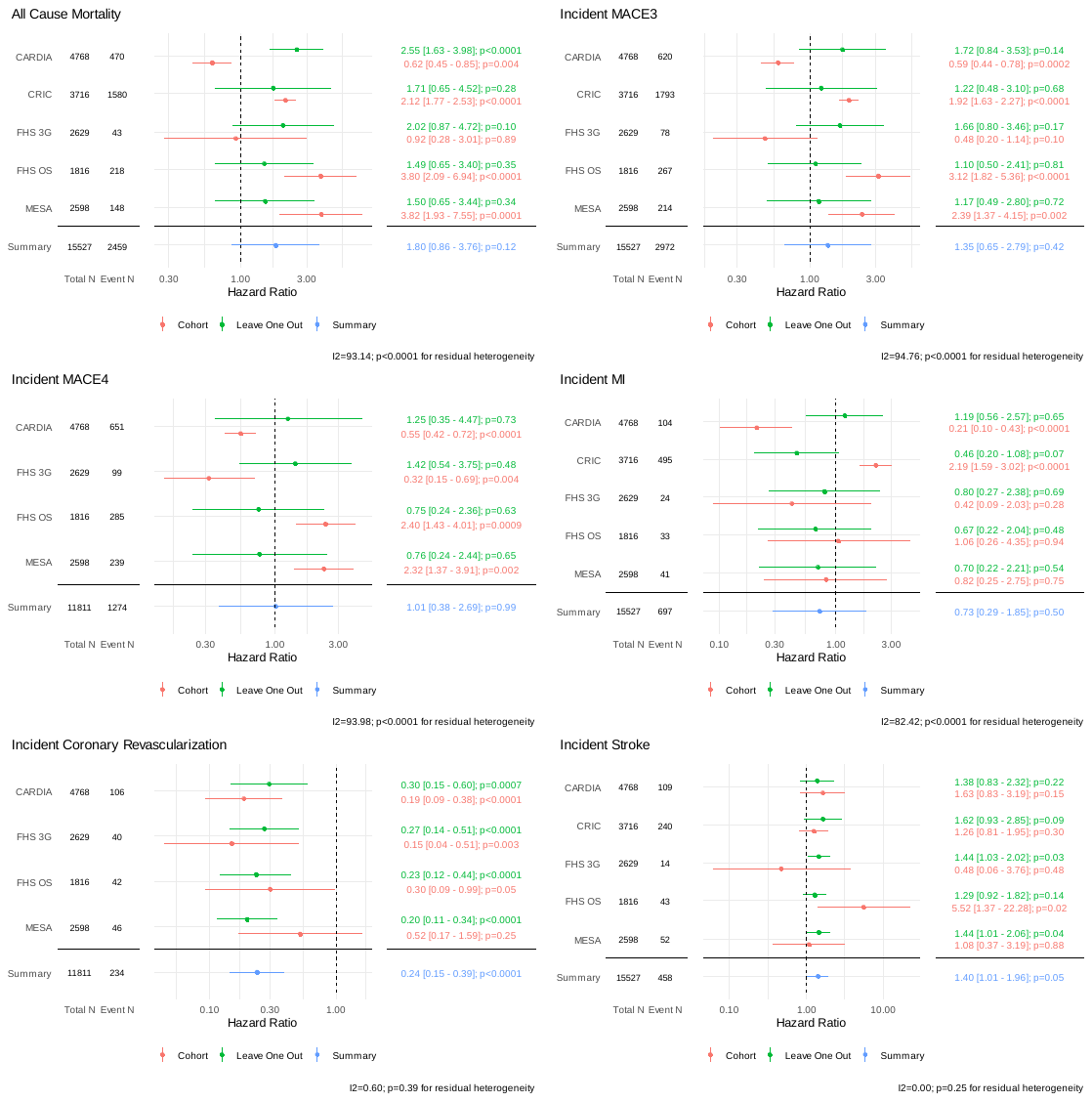

### RBC Quantity Score (Age/Sex/Race Adjusted)

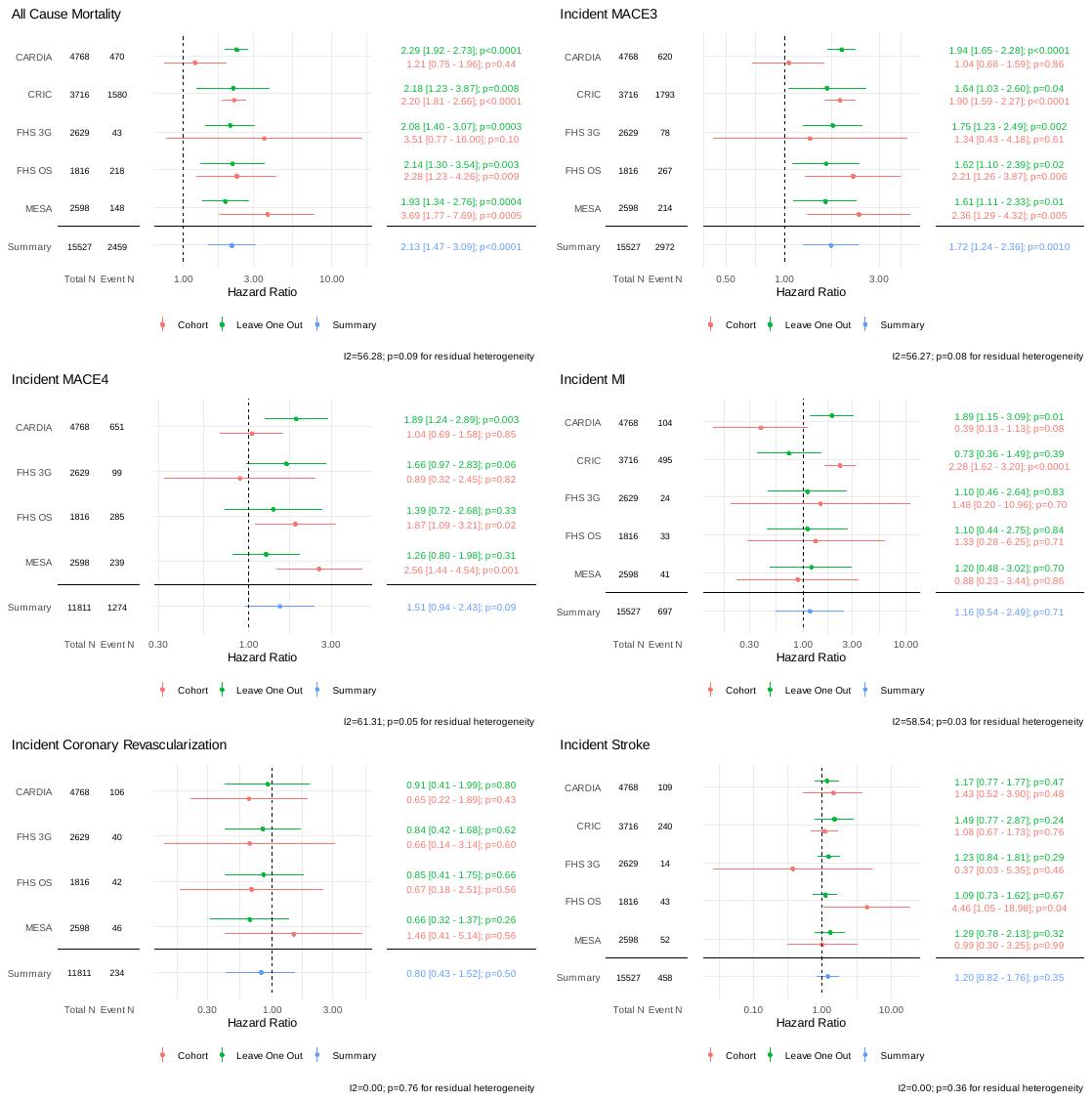

### RBC Quantity Score (Fully Adjusted)

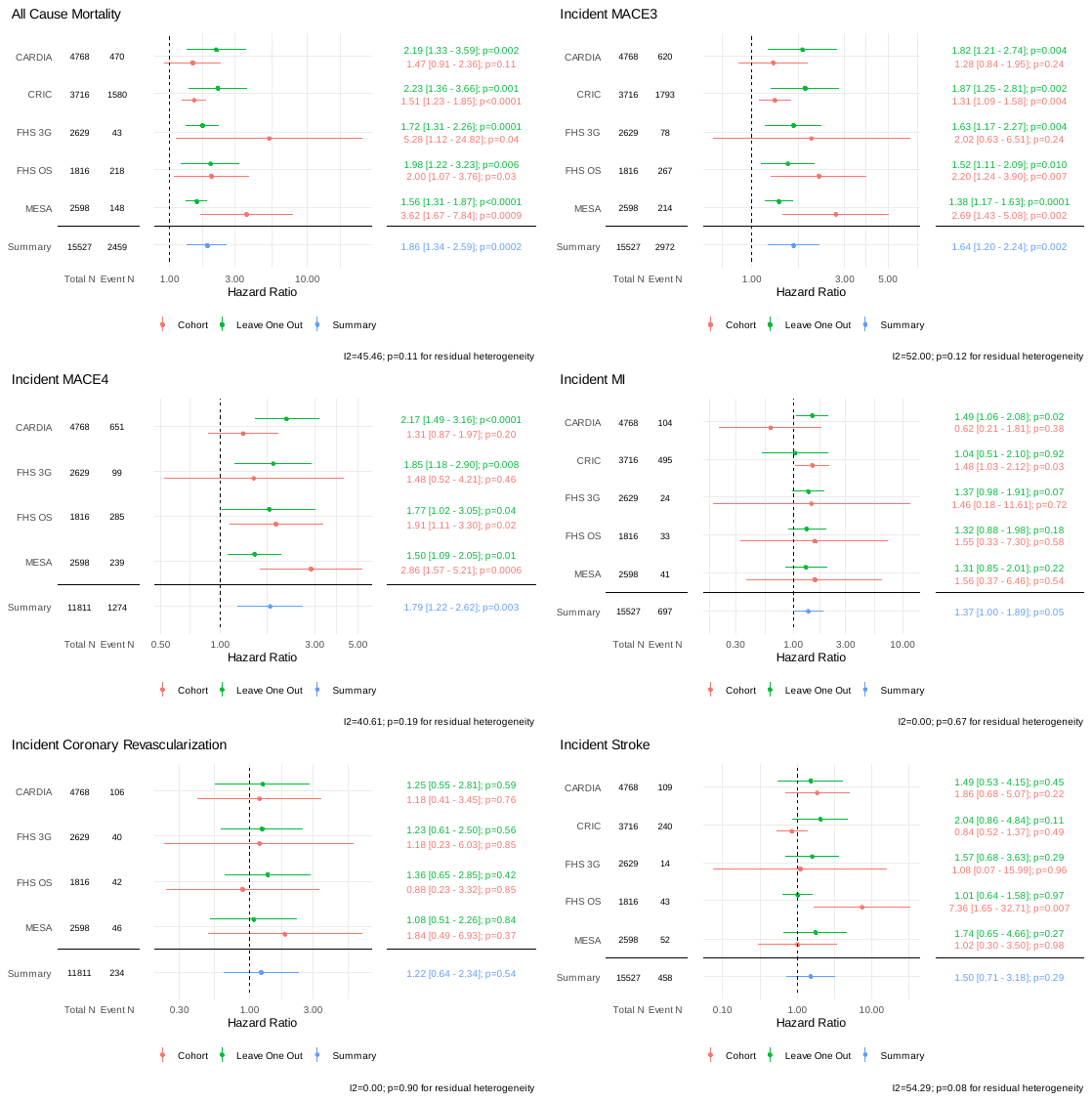

### RBC Quantity Score (Fully Adjusted w/ hsCRP)

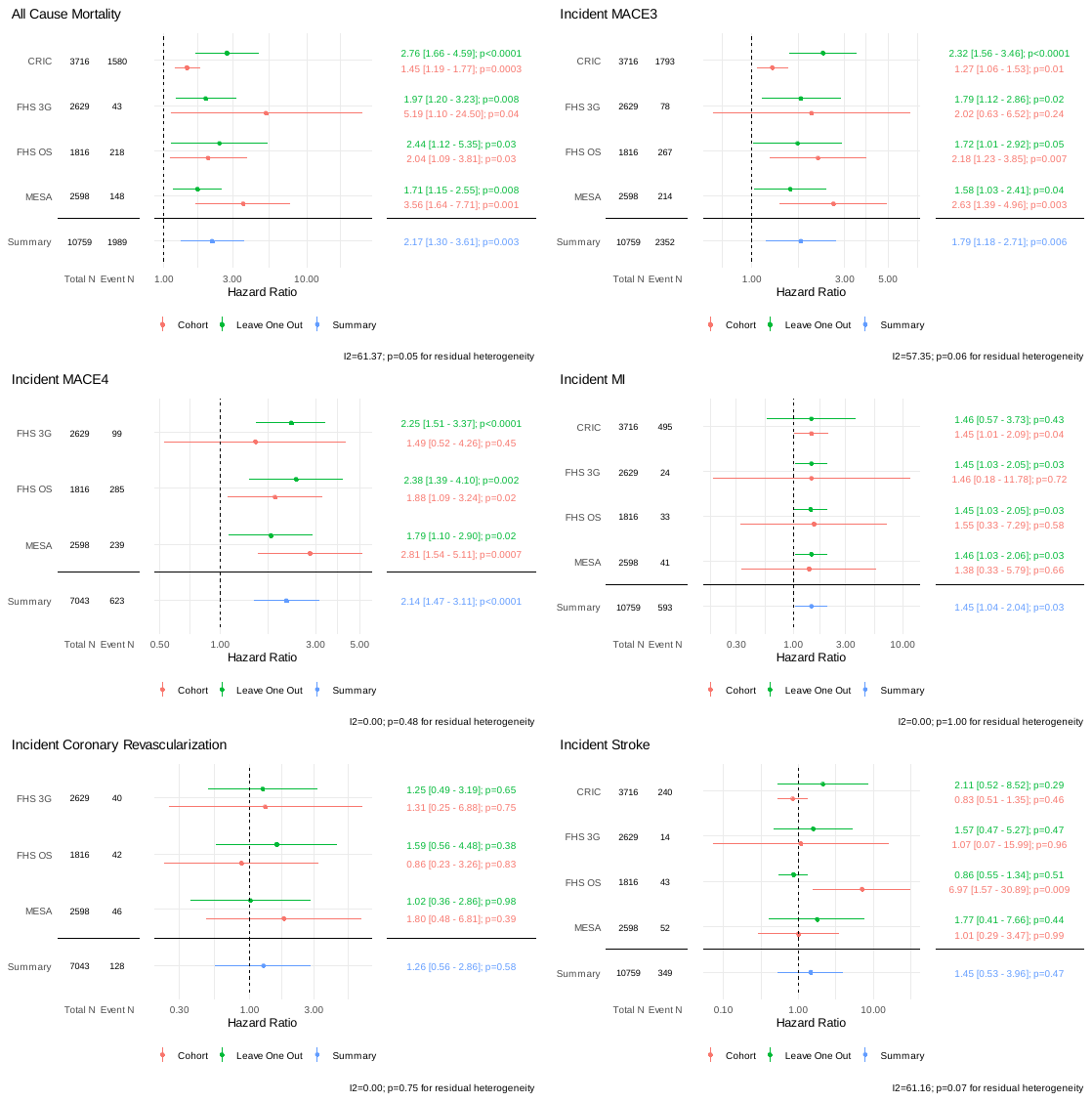

### Other Inflammatory Cells Score (Unadjusted)

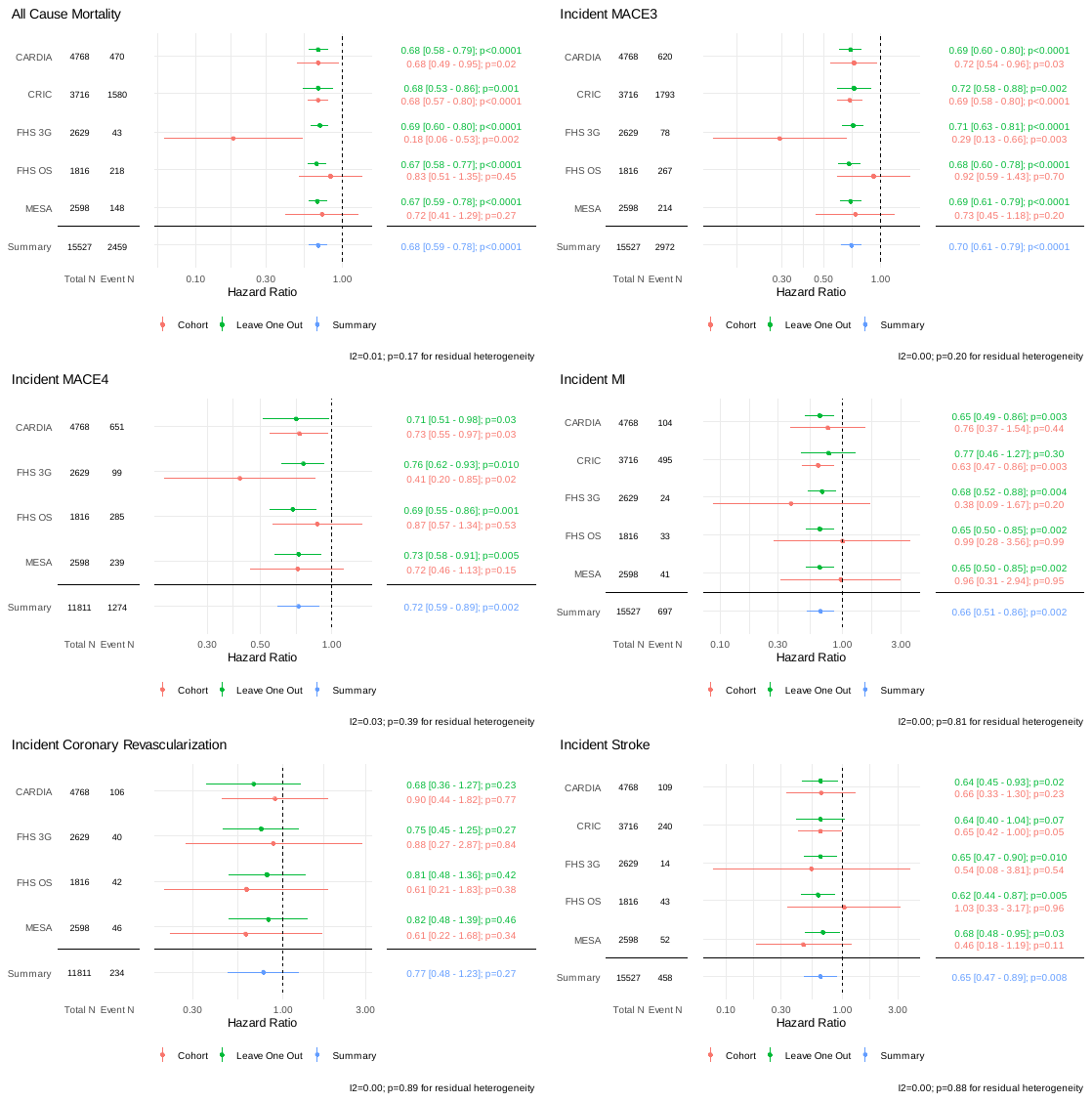

### Other Inflammatory Cells Score (Age/Sex/Race Adjusted)

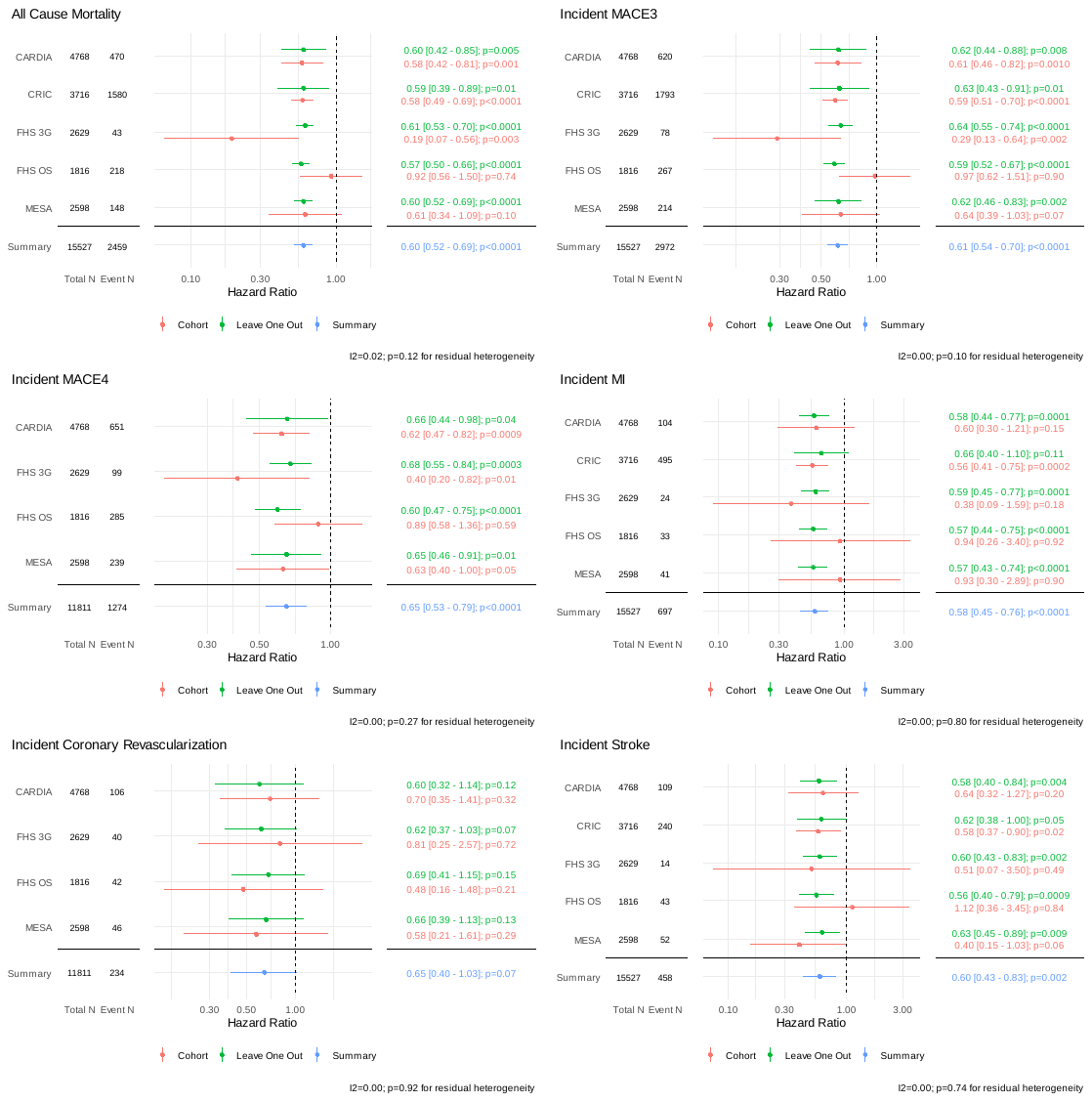

### Other Inflammatory Cells Score (Fully Adjusted)

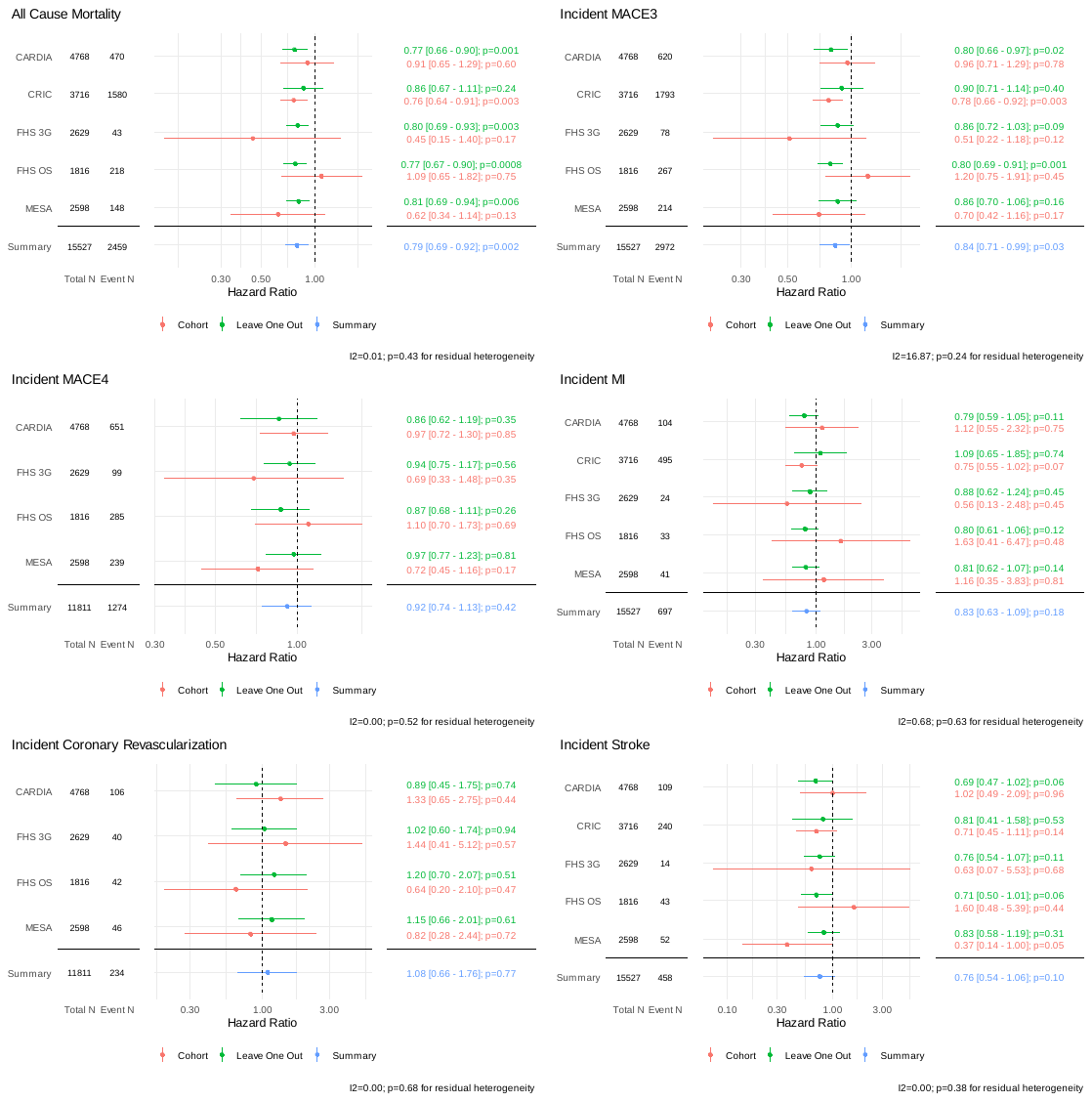

### Other Inflammatory Cells Score (Fully Adjusted w/ hsCRP)

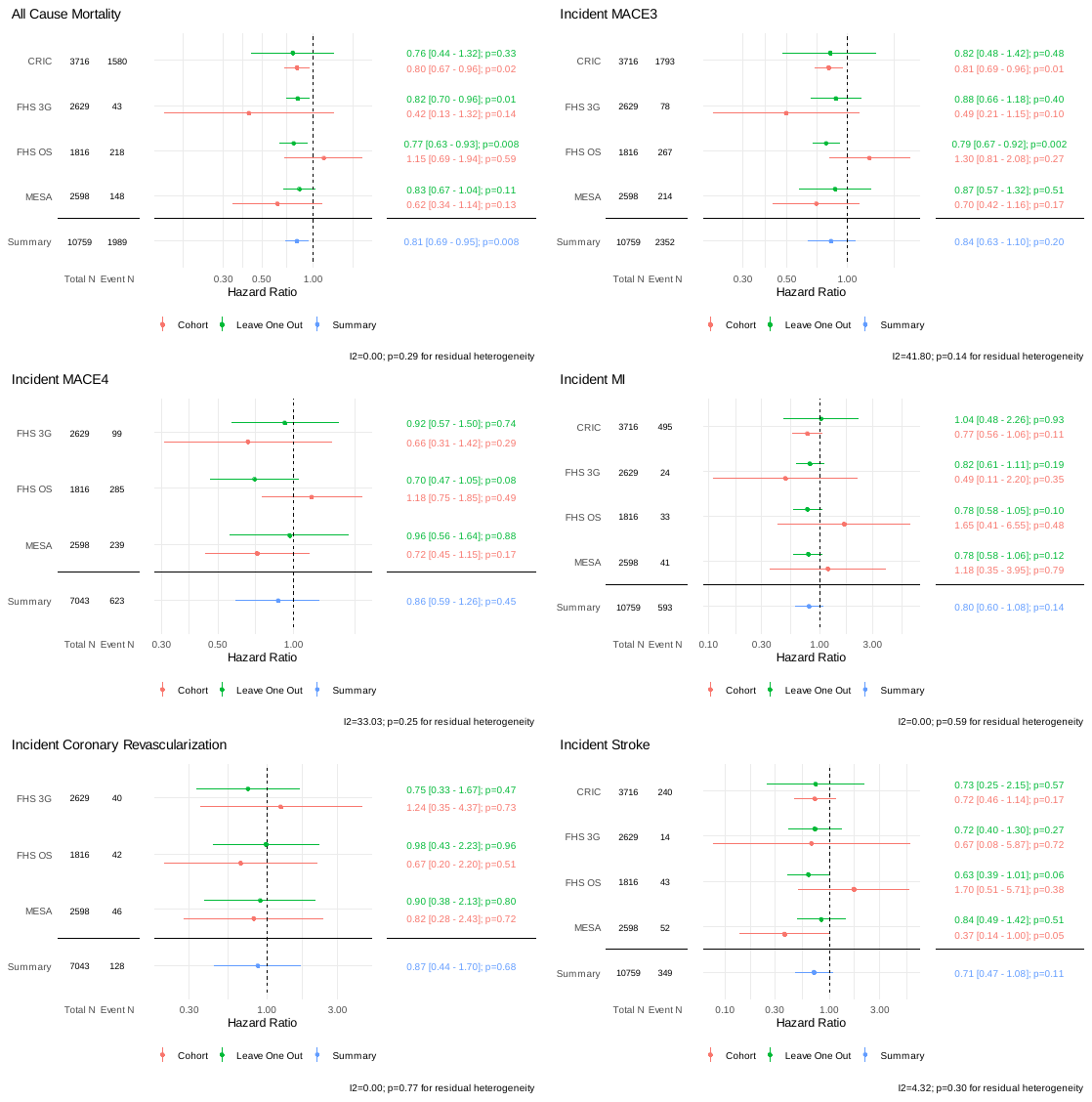

### RBC Properties Score (Unadjusted)

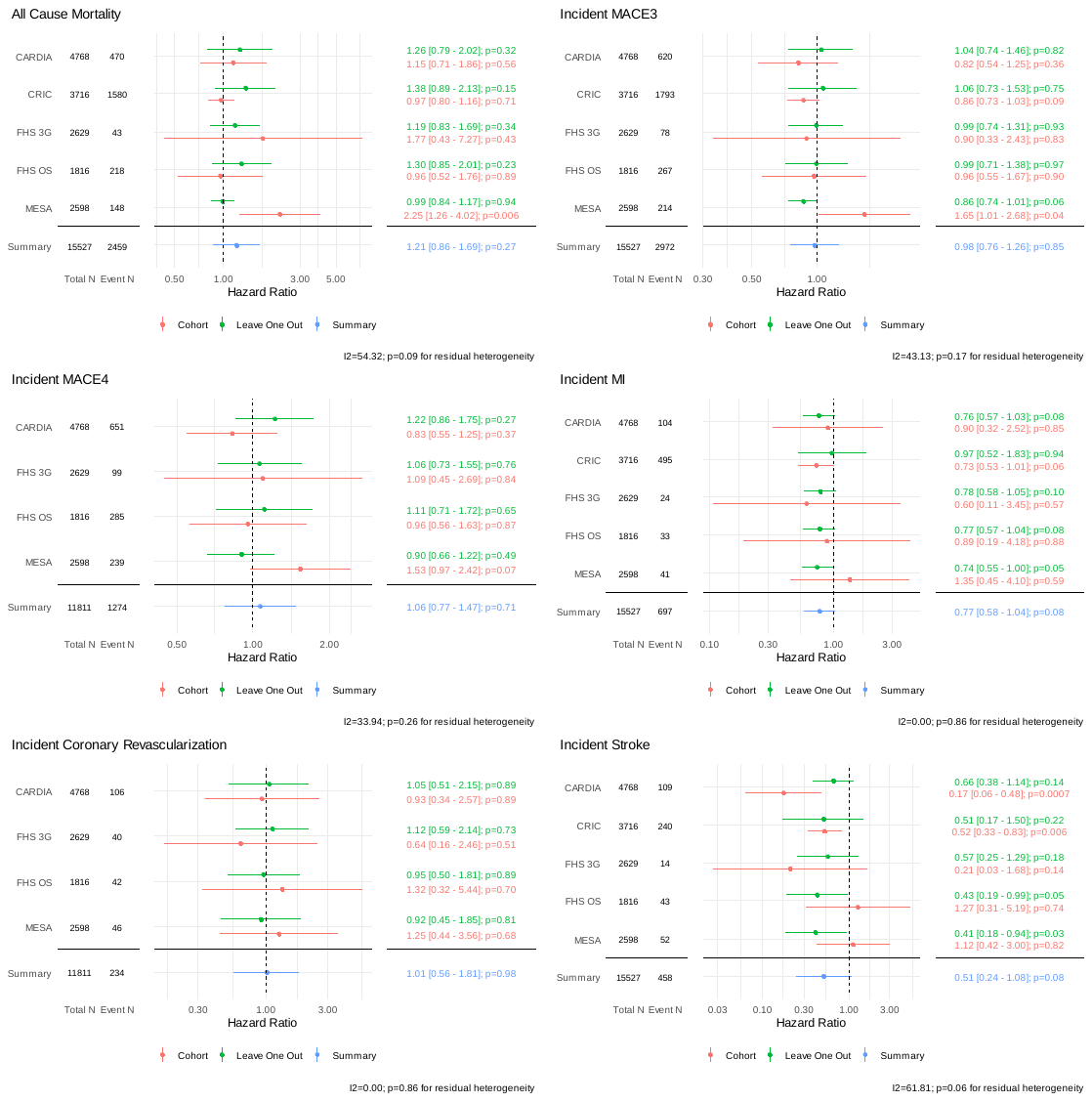

### RBC Properties Score (Age/Sex/Race Adjusted)

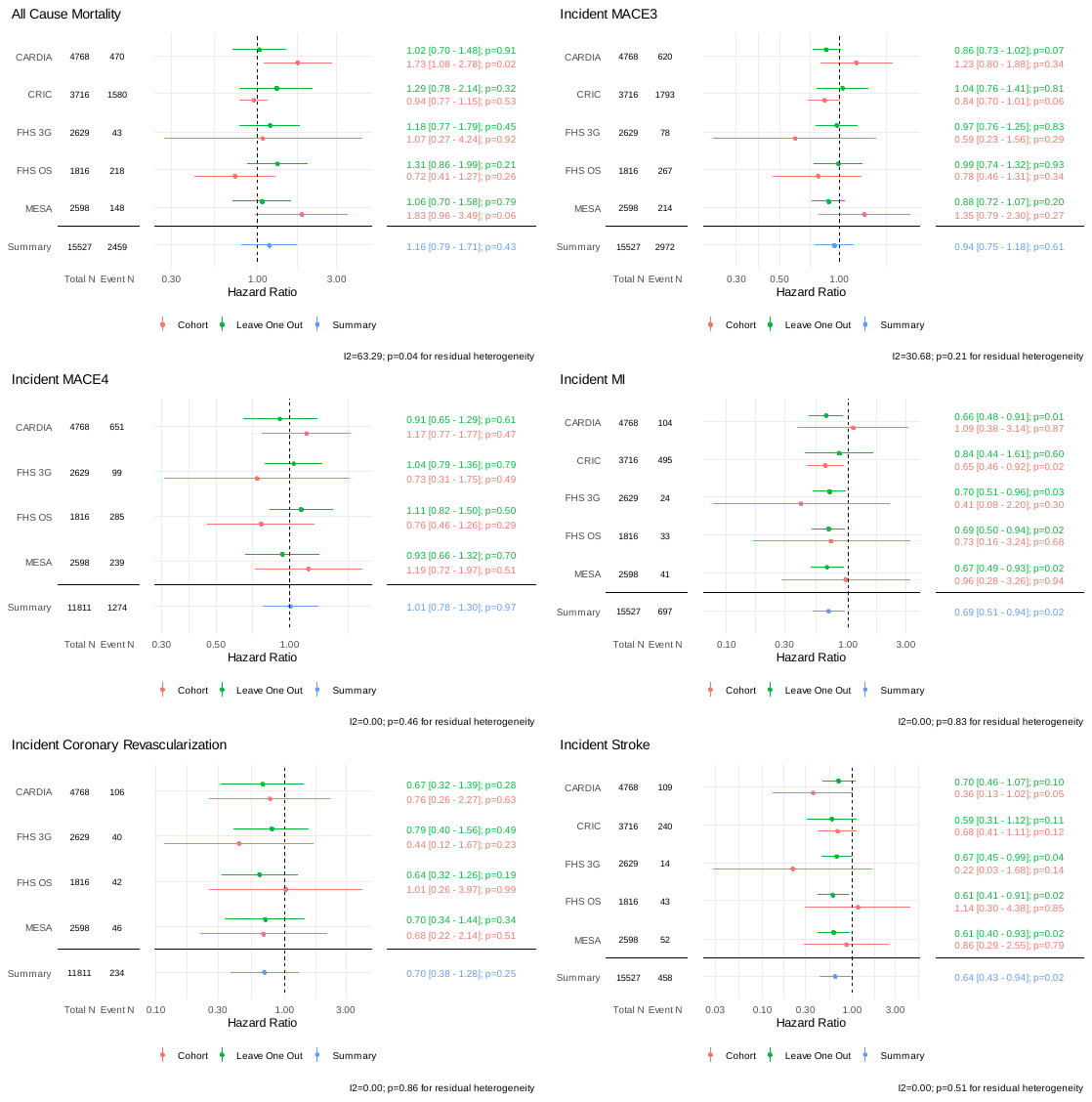

### RBC Properties Score (Fully Adjusted)

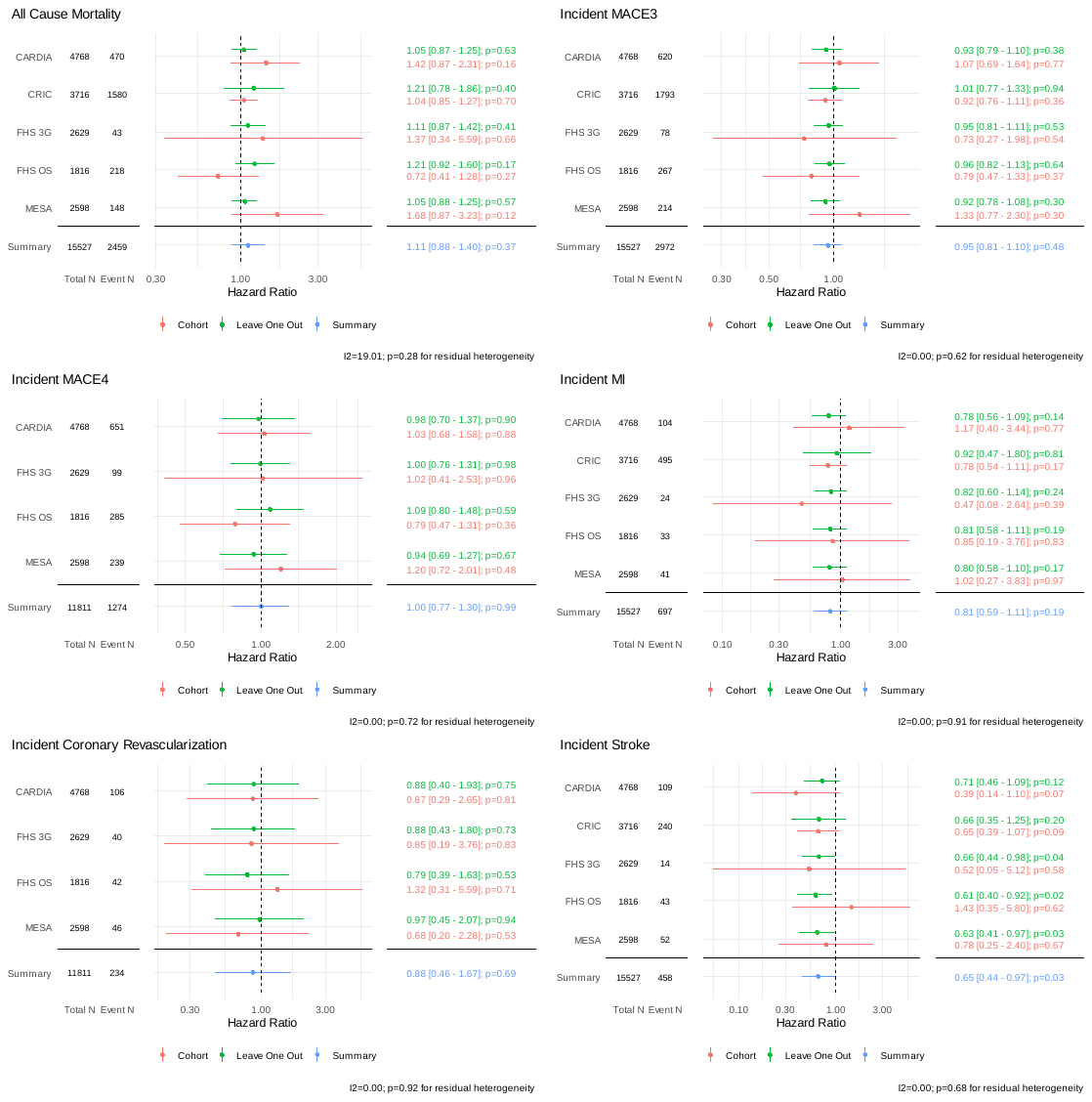

### RBC Properties Score (Fully Adjusted w/ hsCRP)

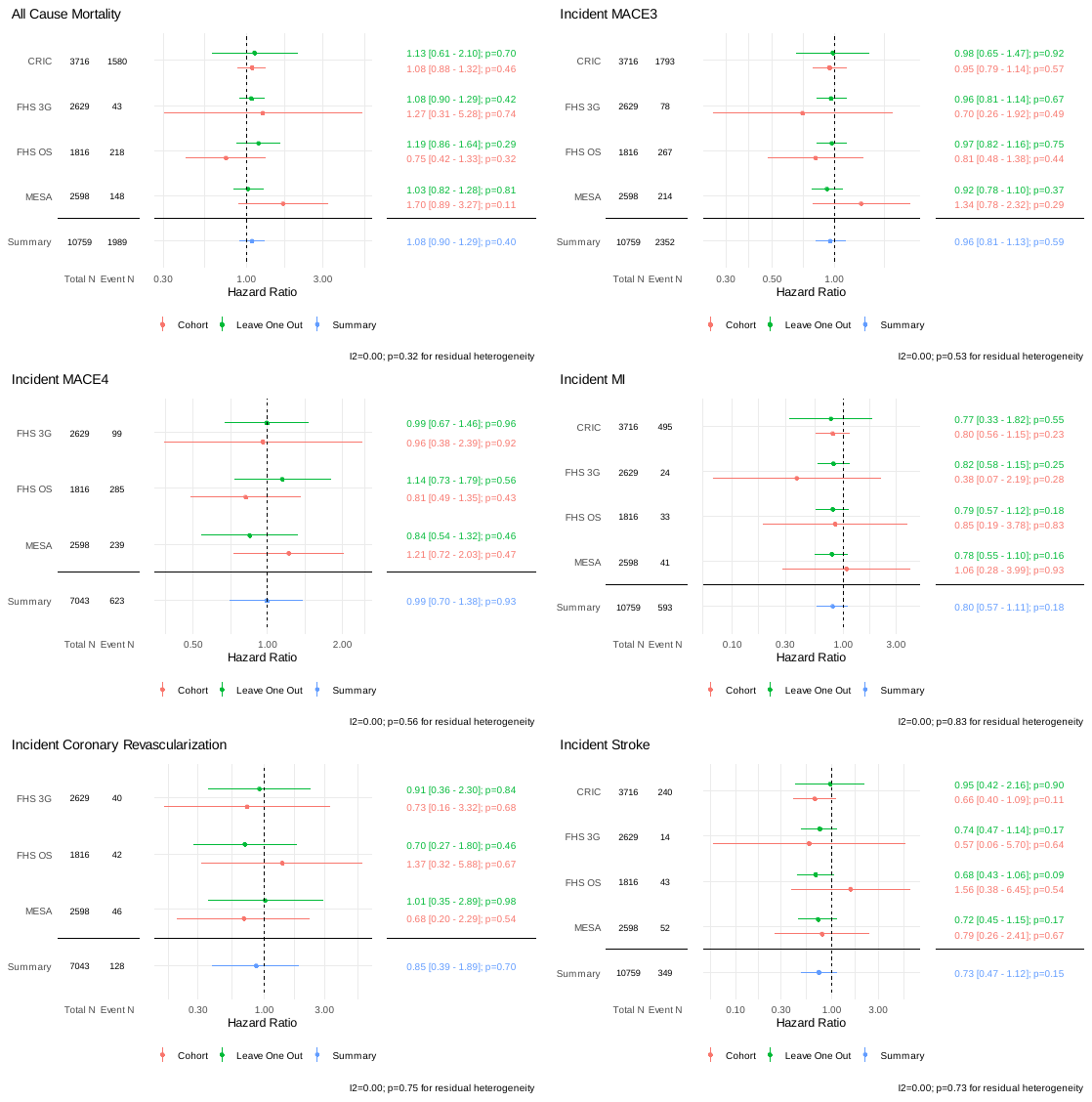

### Basophil Count (Unadjusted)

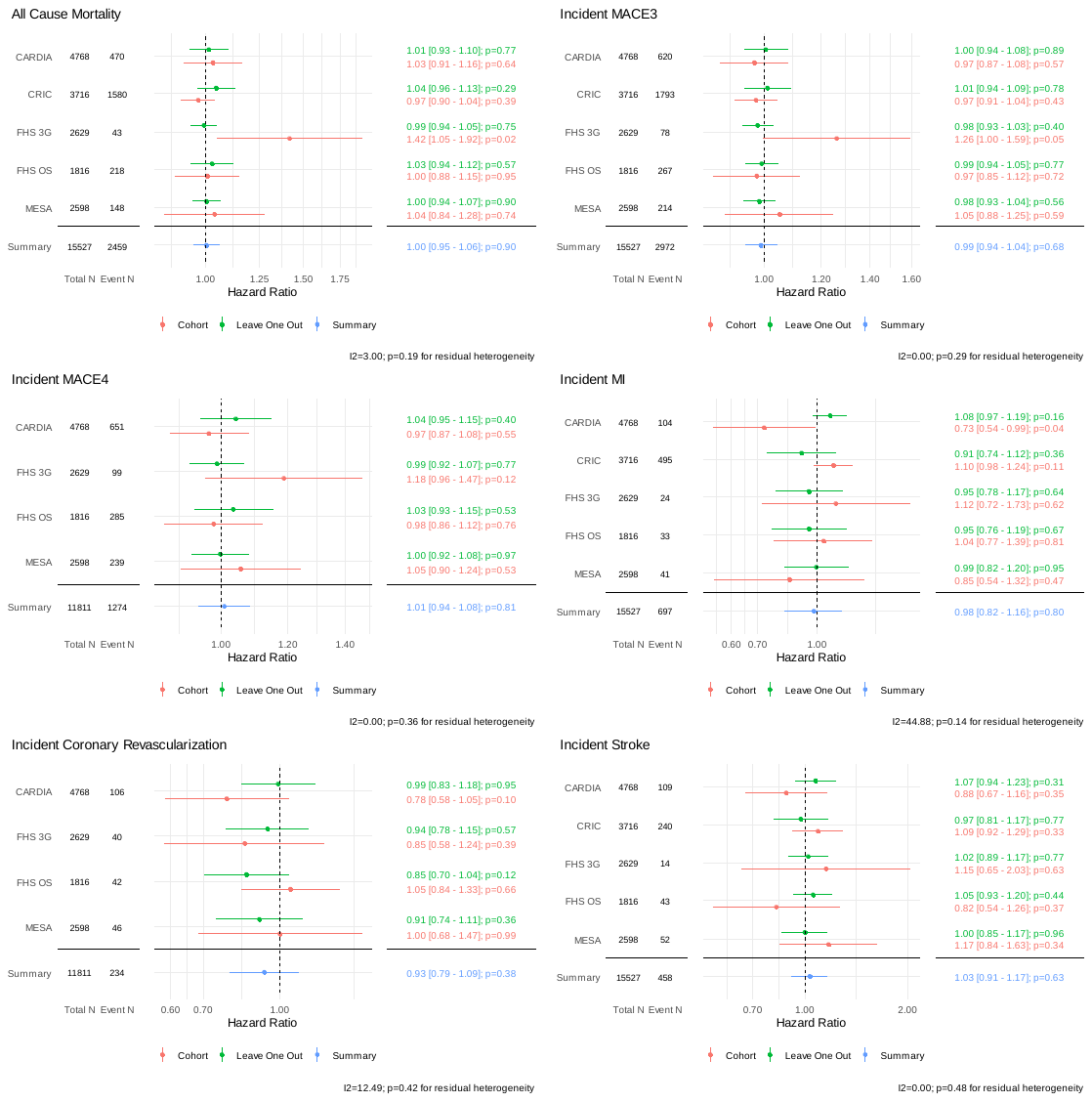

### Basophil Count (Age/Sex/Race Adjusted)

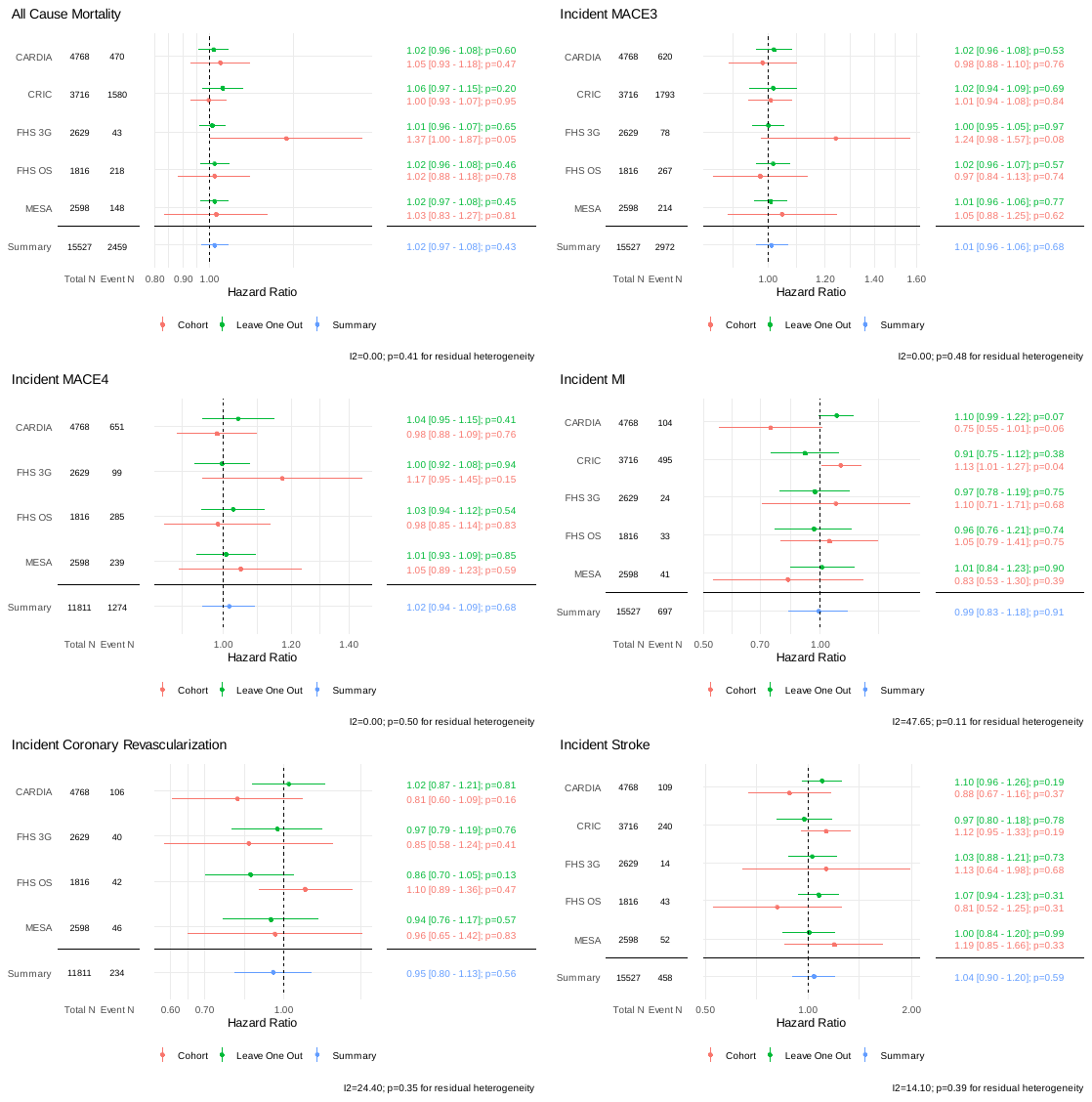

### Basophil Count (Fully Adjusted)

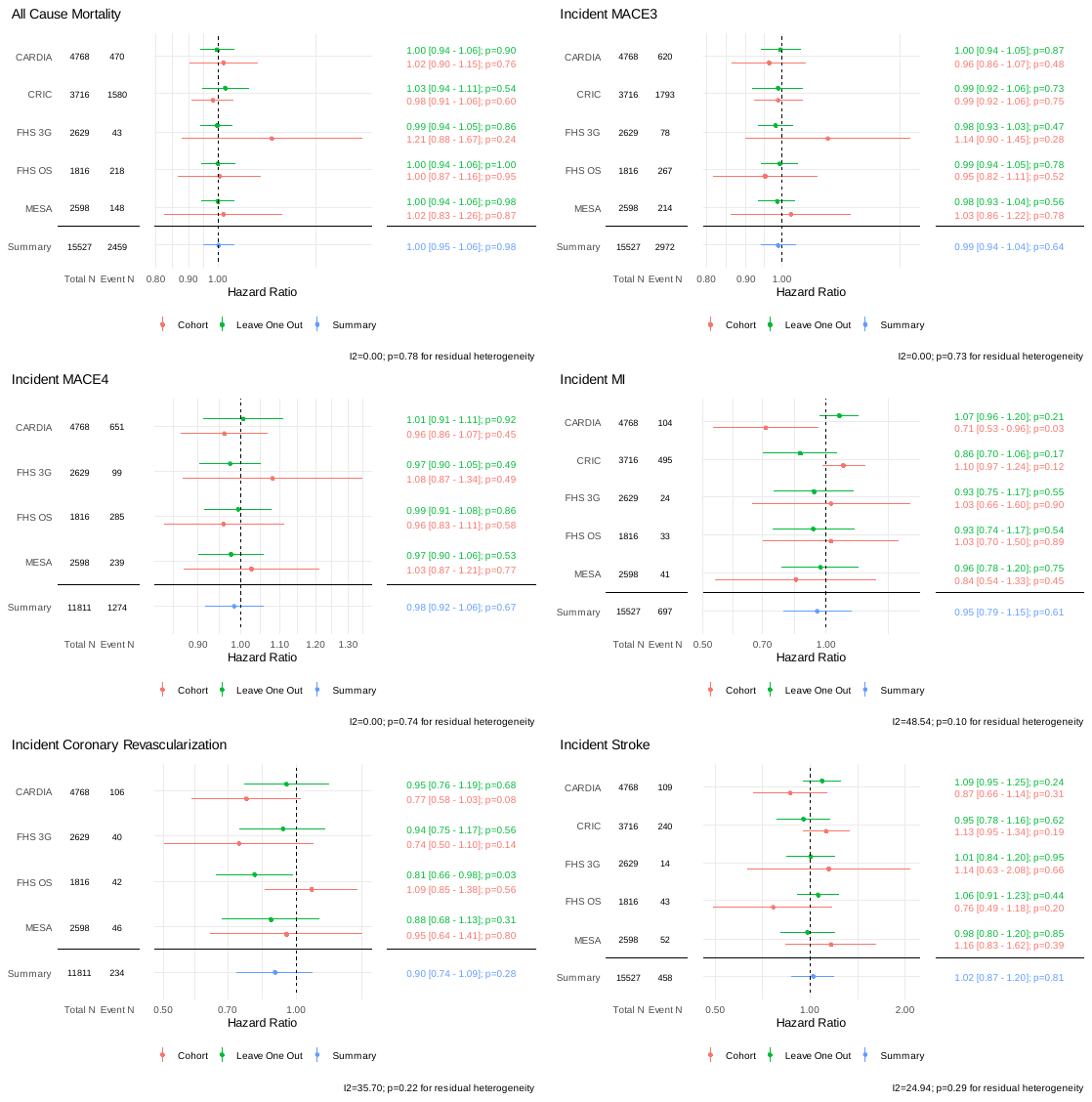

### Basophil Count (Fully Adjusted w/ hsCRP)

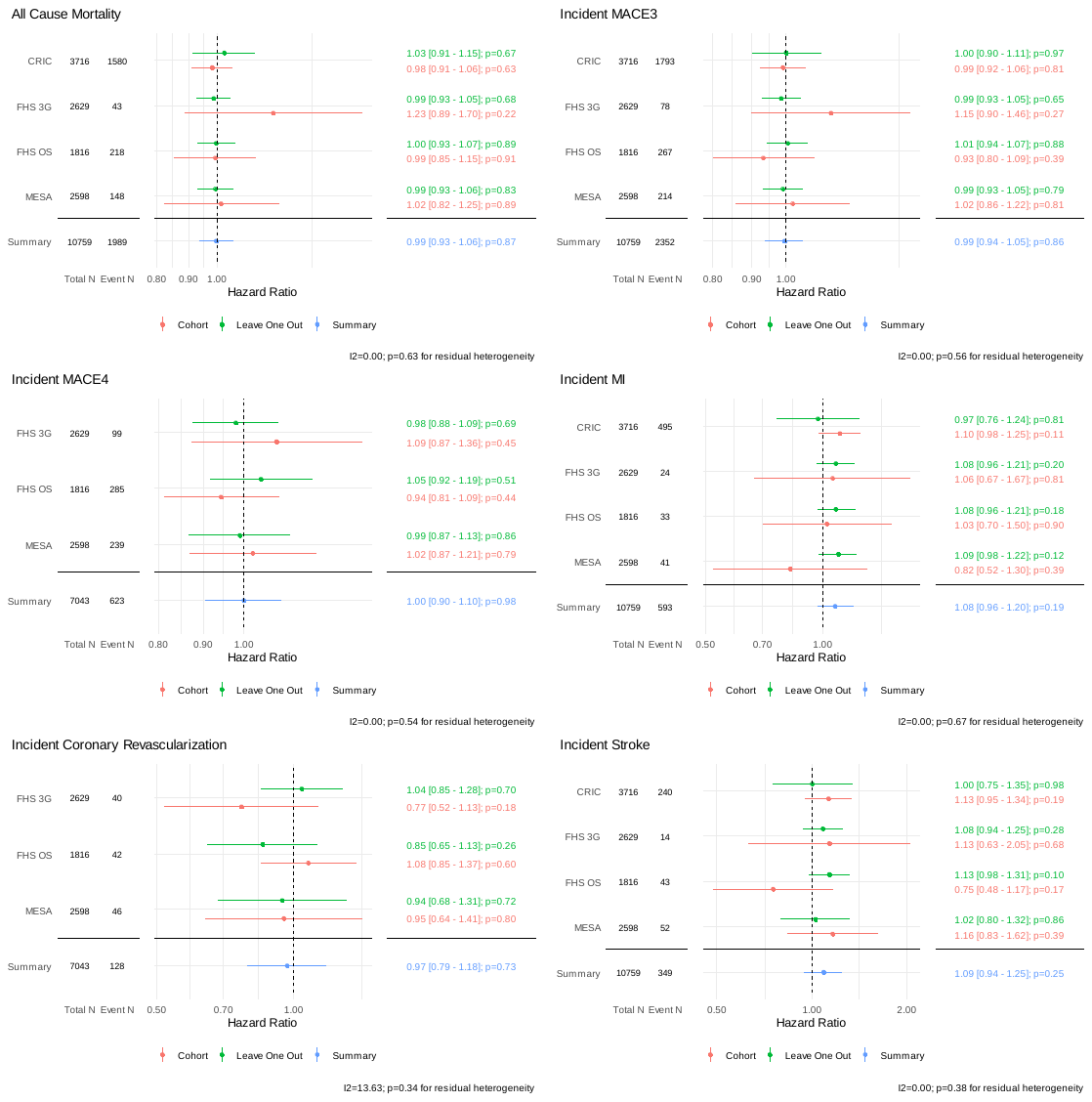

### Basophils (%) (Unadjusted)

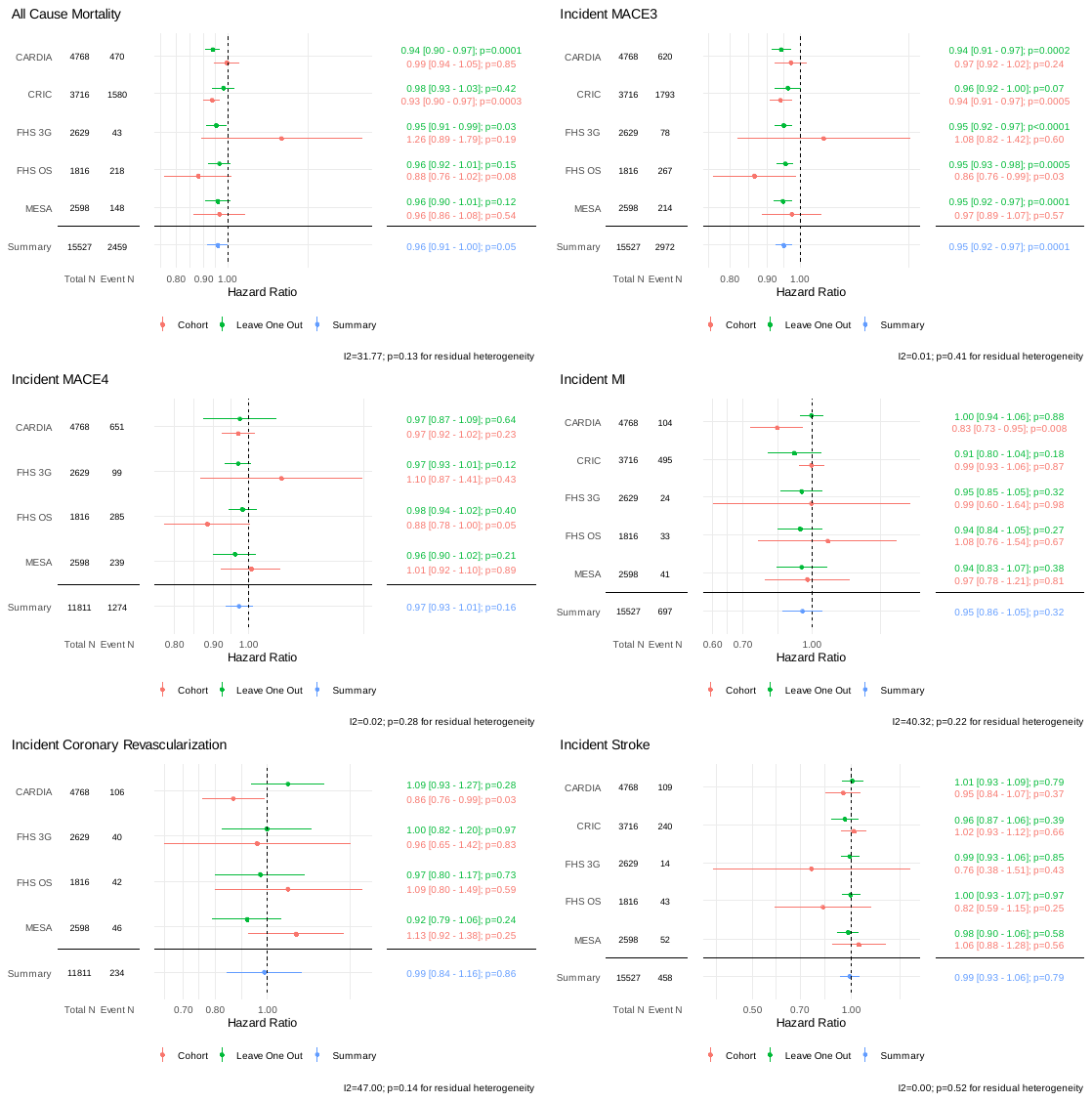

### Basophils (%) (Age/Sex/Race Adjusted)

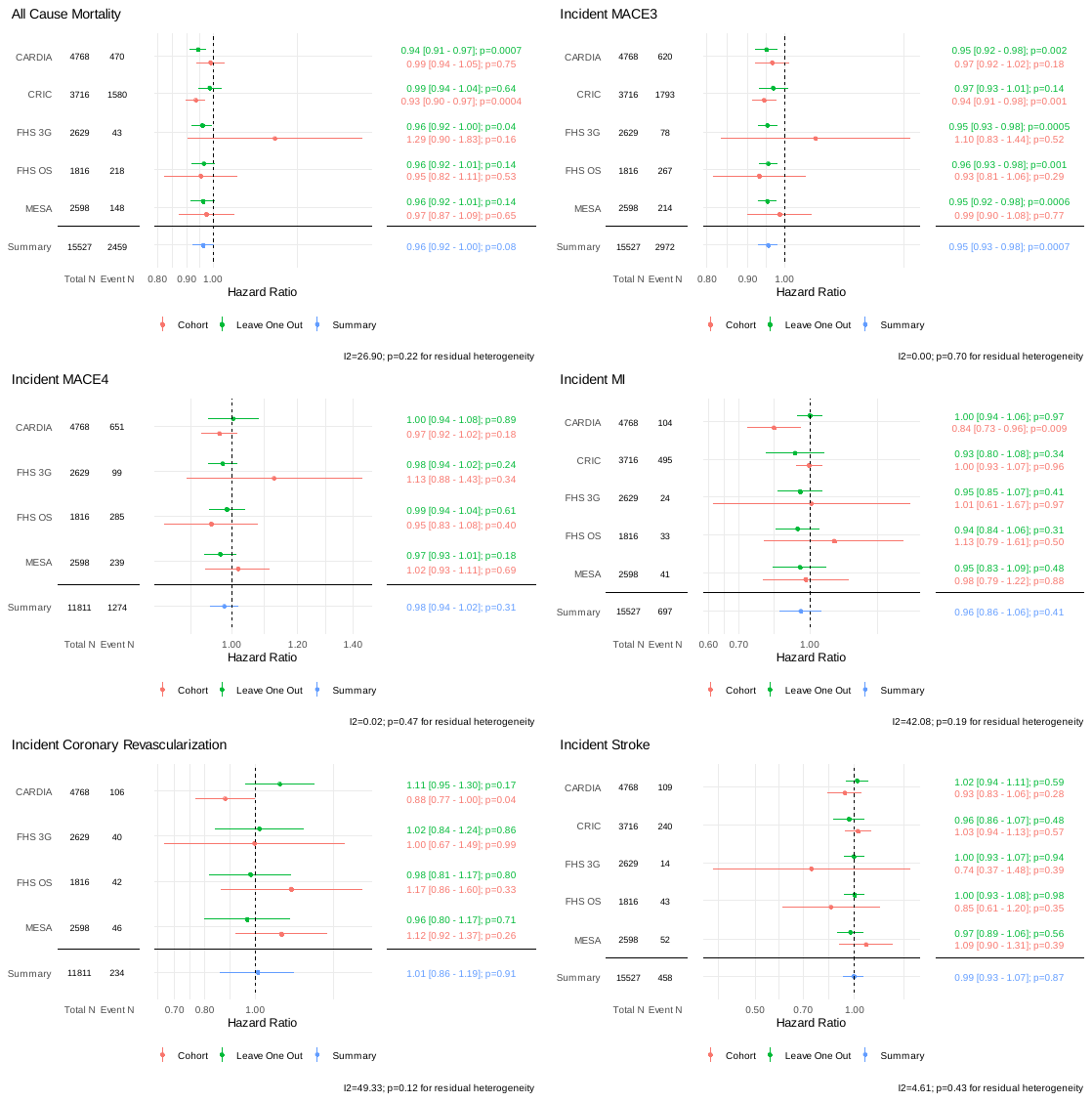

### Basophils (%) (Fully Adjusted)

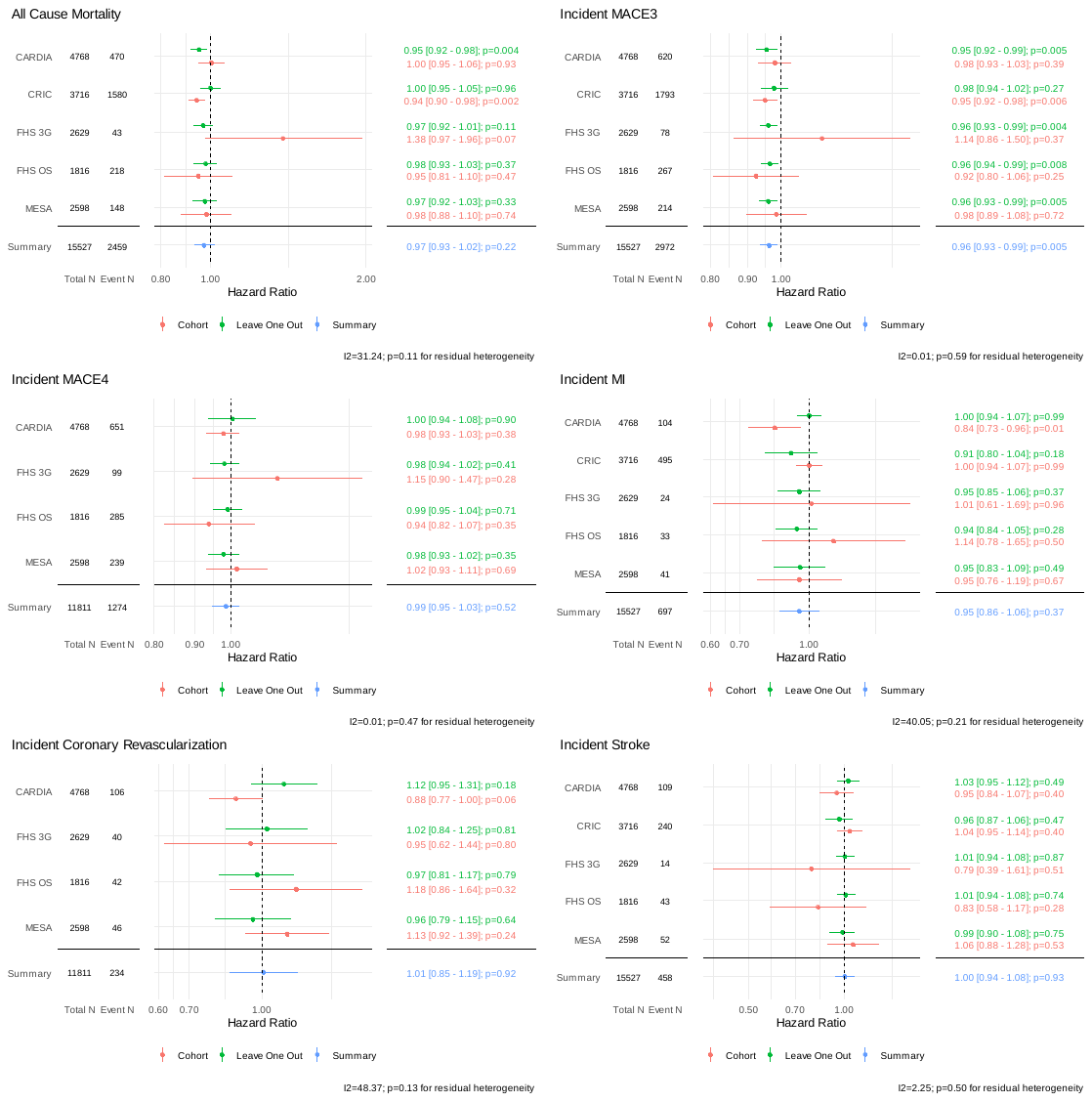

### Basophils (%) (Fully Adjusted w/ hsCRP)

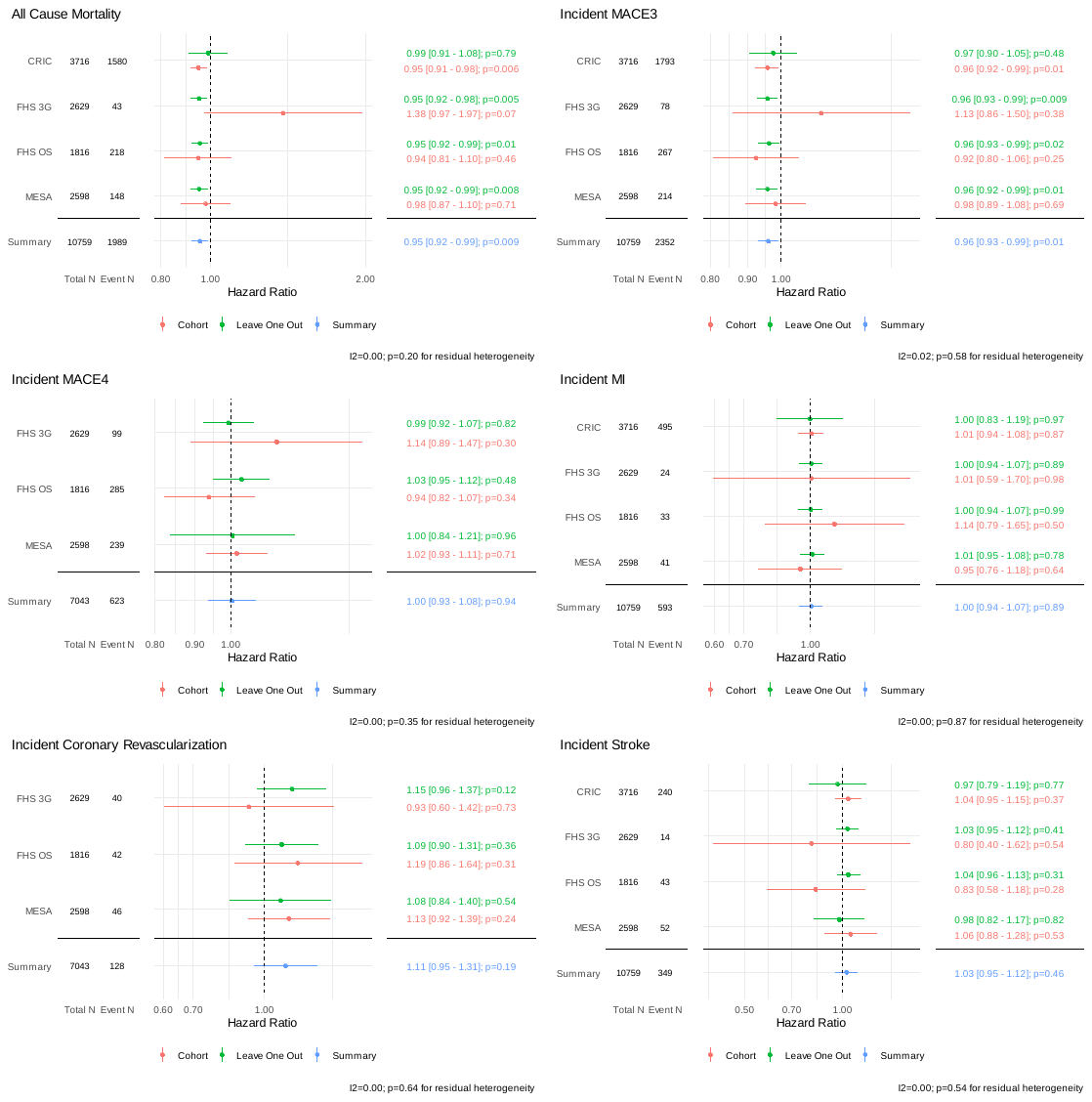

### Eosinophil Count (Unadjusted)

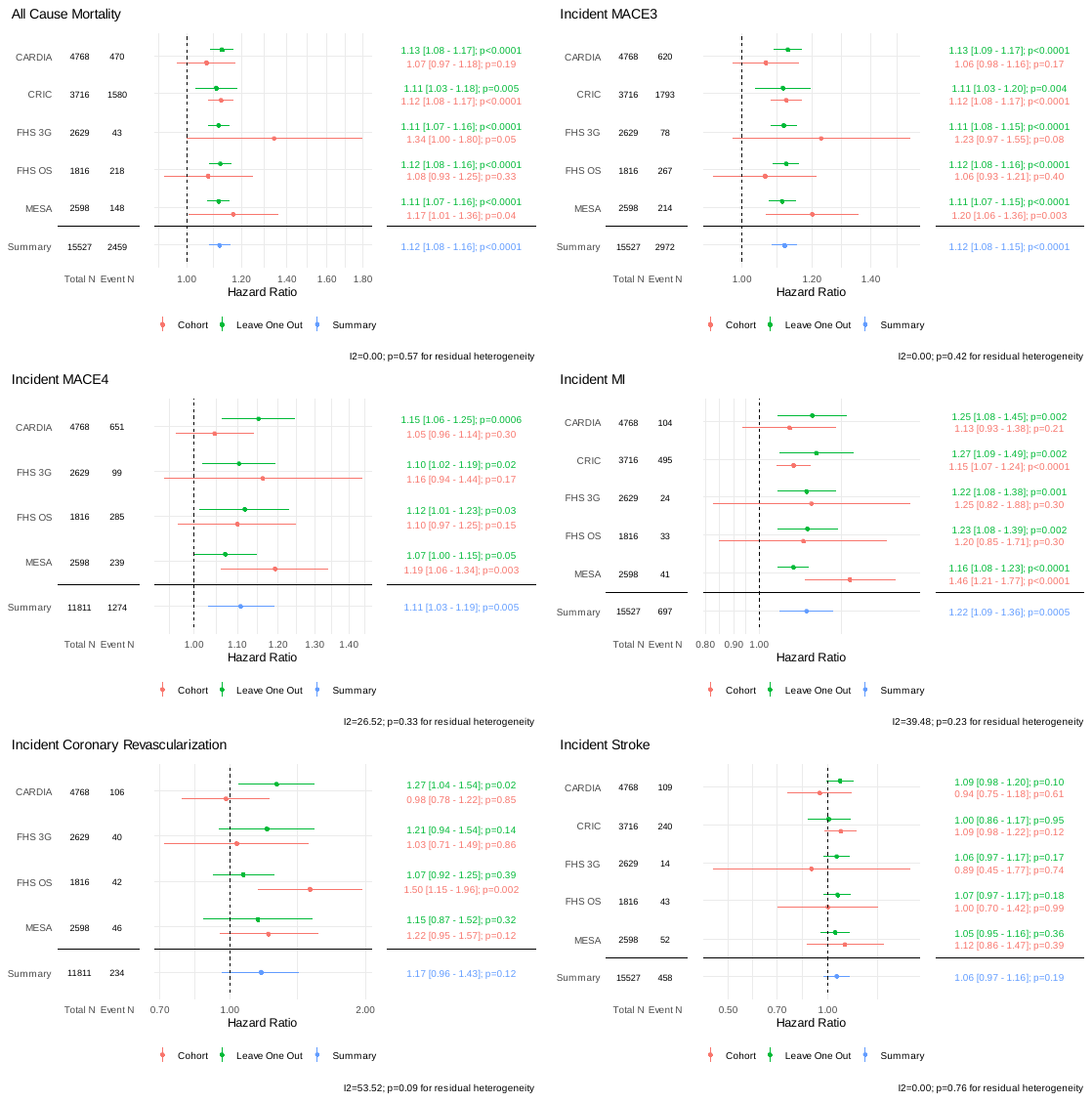

### Eosinophil Count (Age/Sex/Race Adjusted)

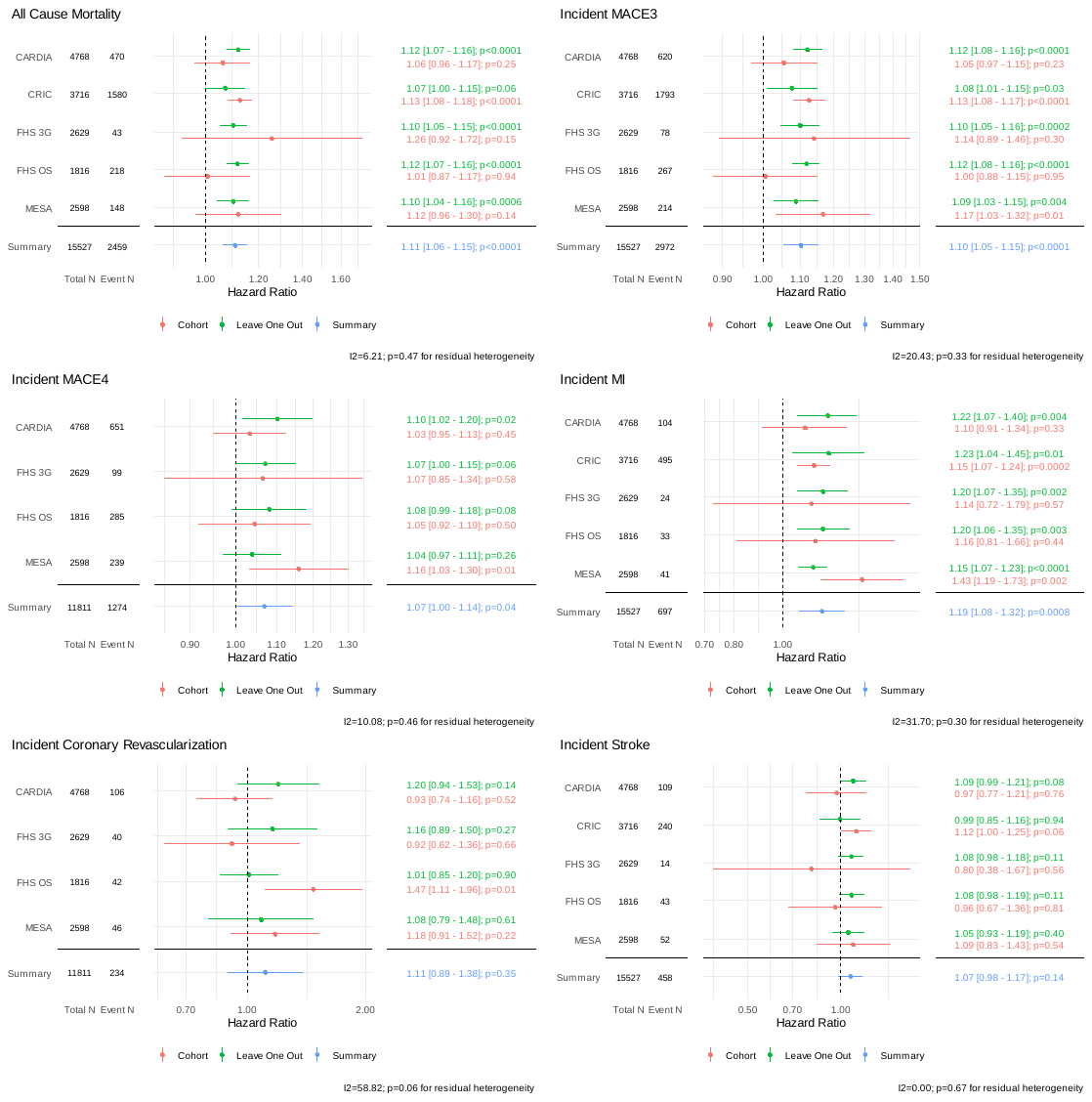

### Eosinophil Count (Fully Adjusted)

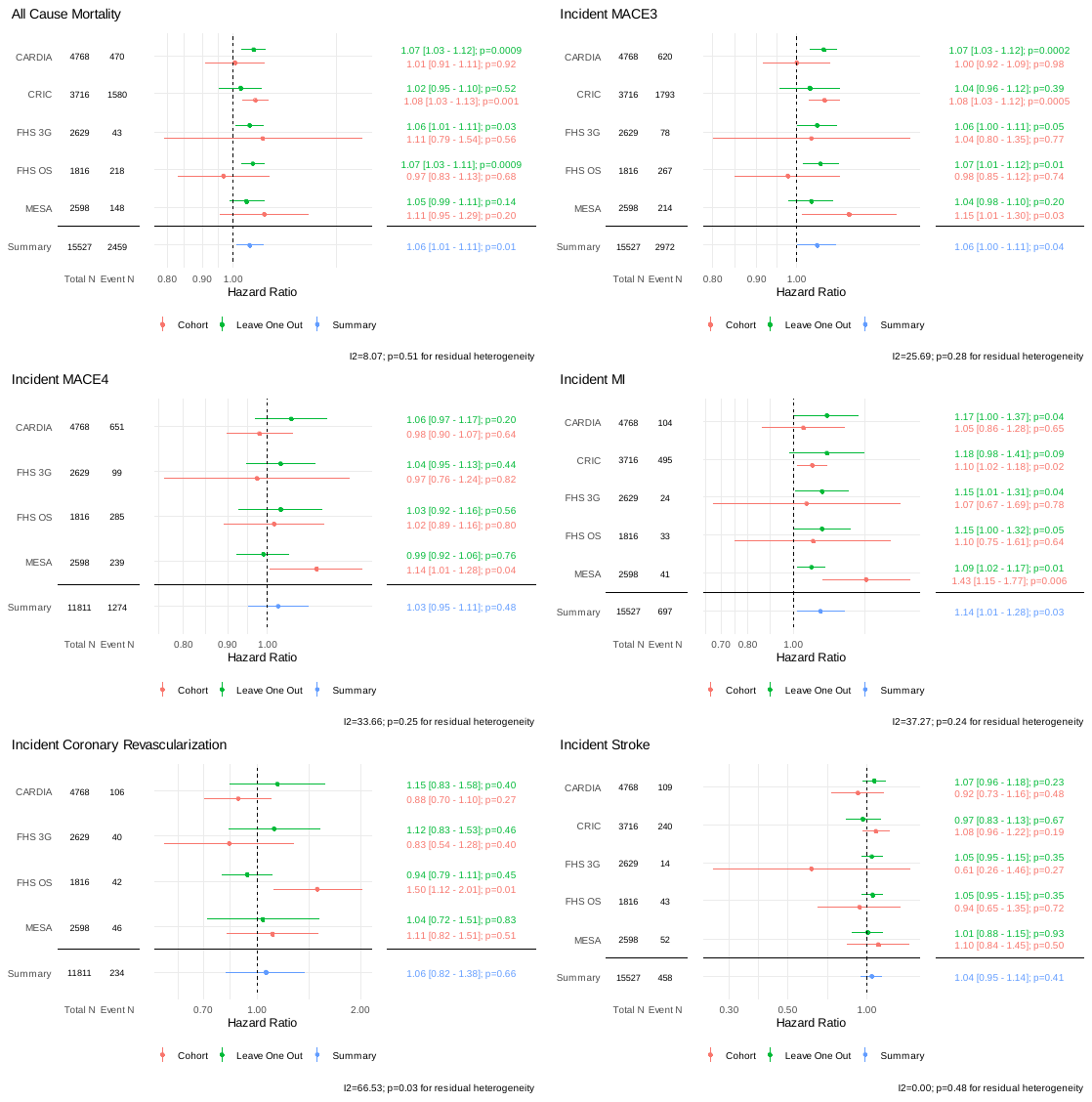

### Eosinophil Count (Fully Adjusted w/ hsCRP)

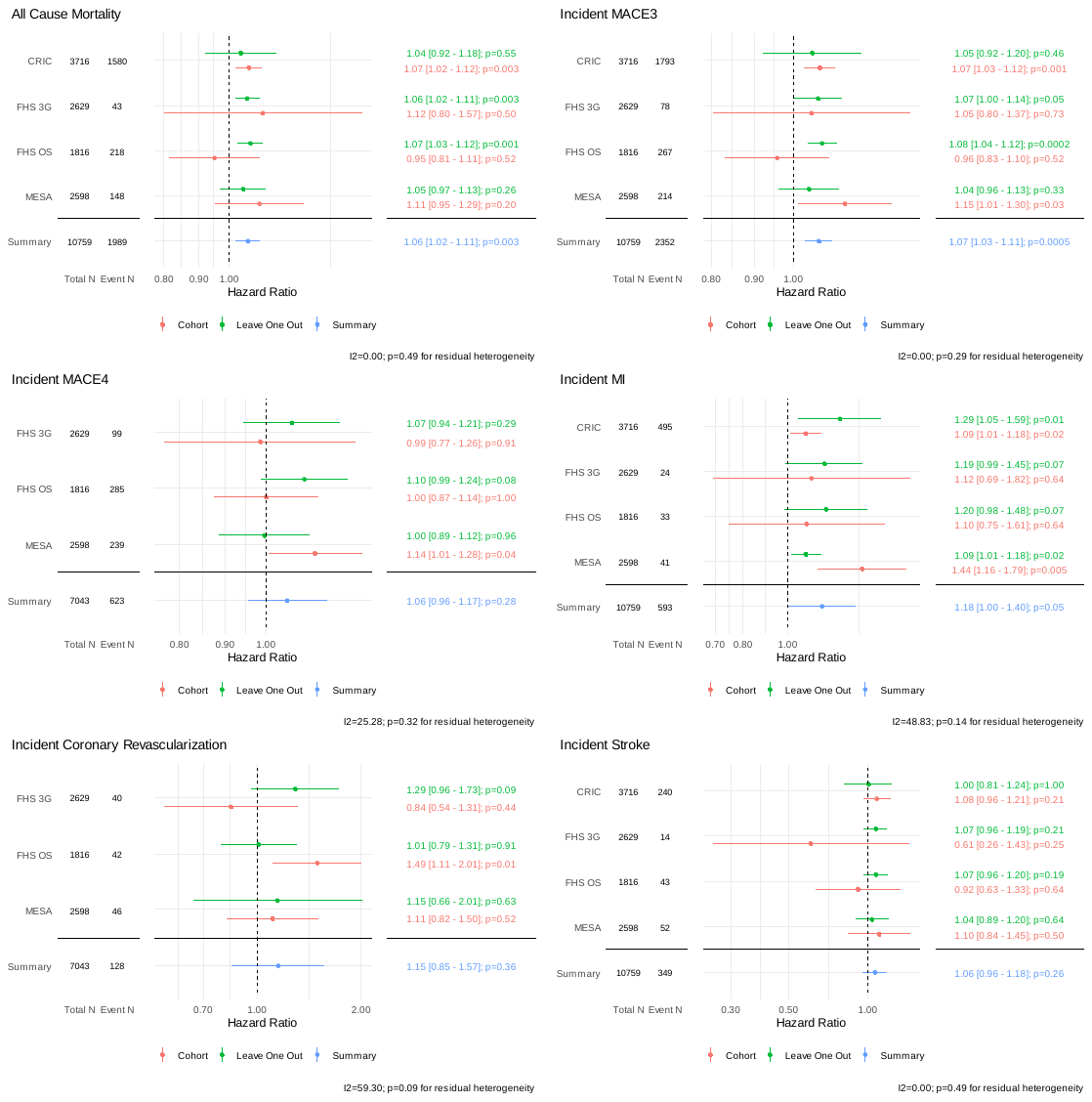

### Eosinophils (%) (Unadjusted)

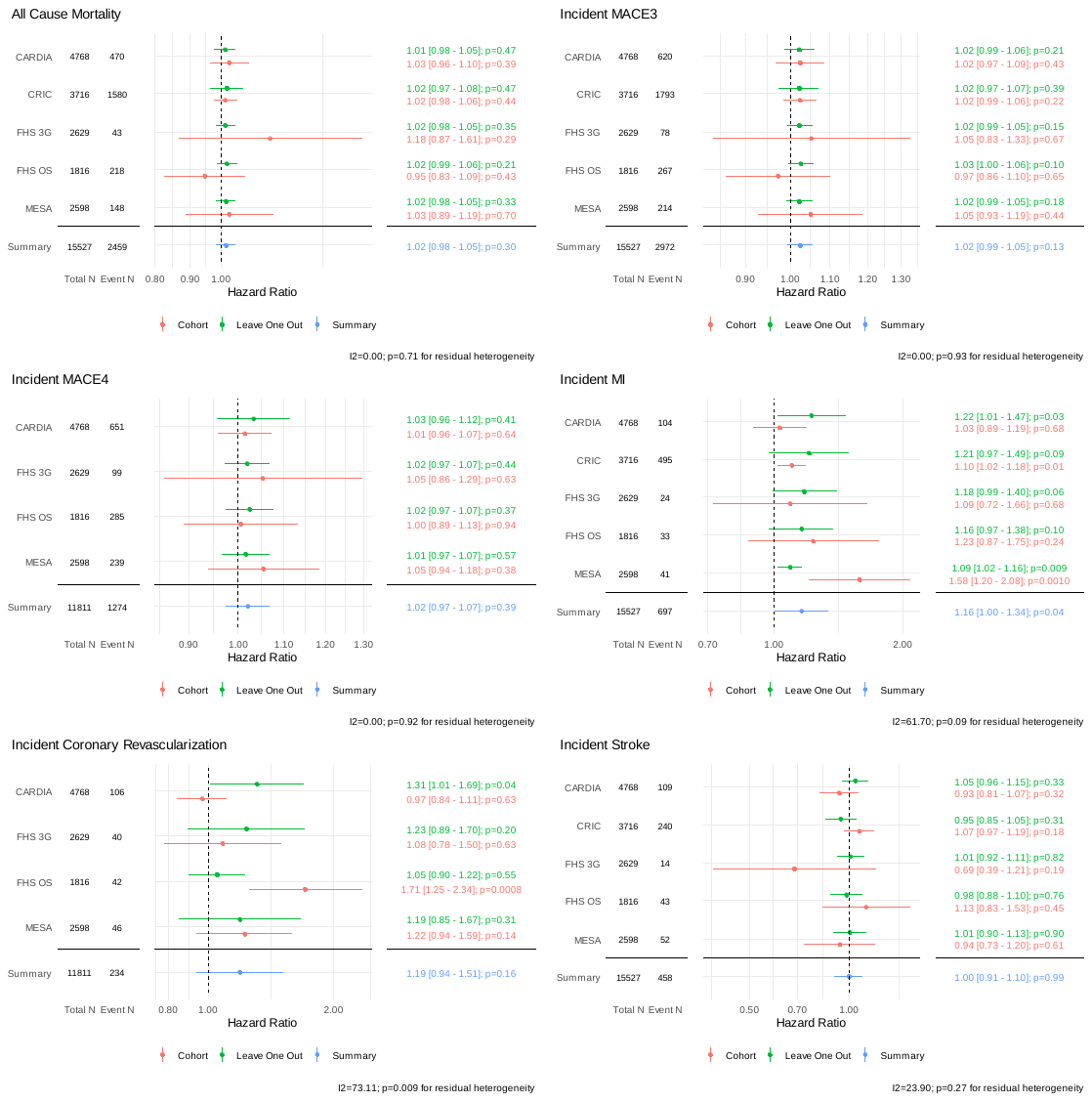

### Eosinophils (%) (Age/Sex/Race Adjusted)

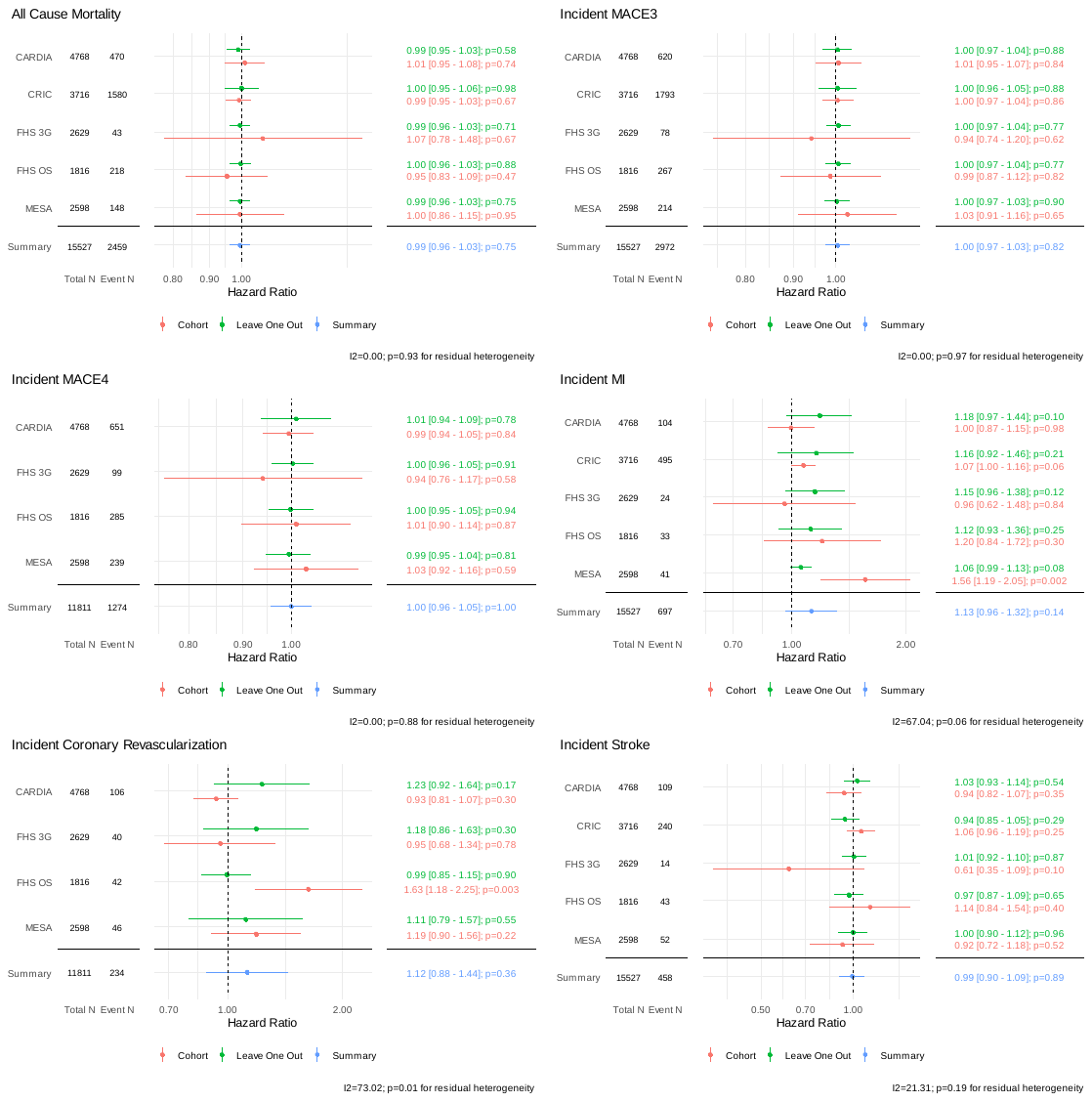

### Eosinophils (%) (Fully Adjusted)

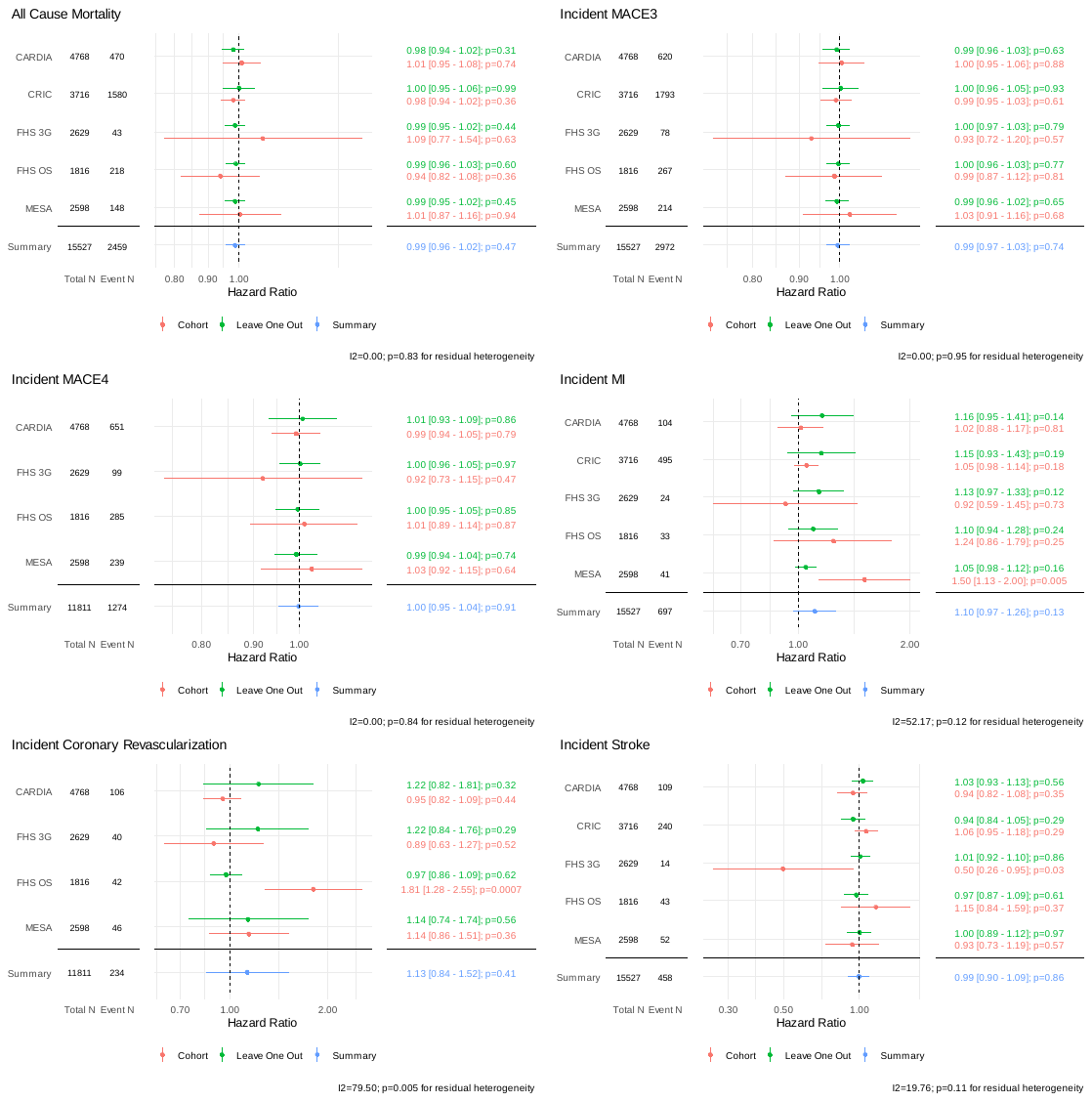

### Eosinophils (%) (Fully Adjusted w/ hsCRP)

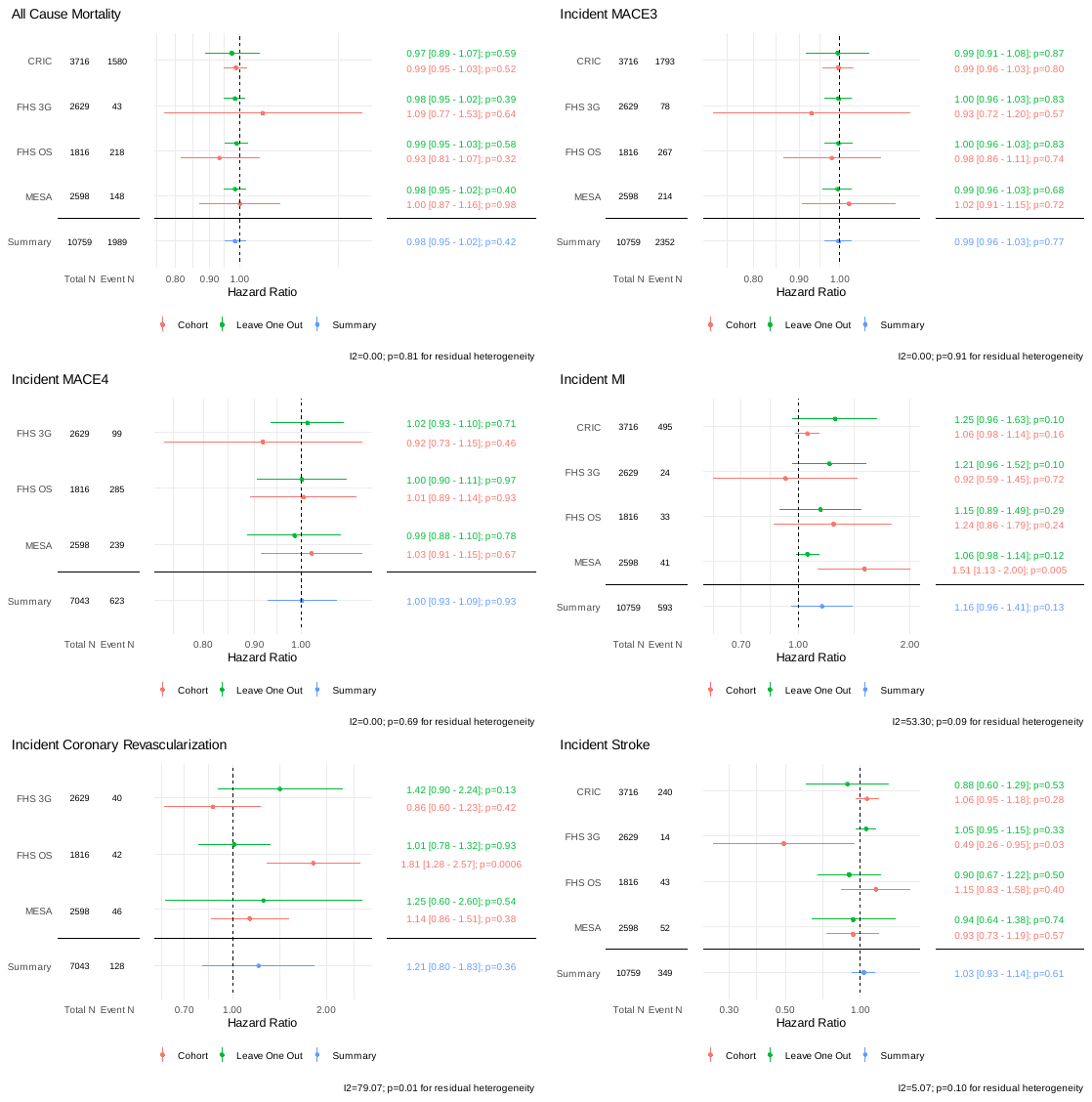

### Immature Granulocyte Count (Unadjusted)

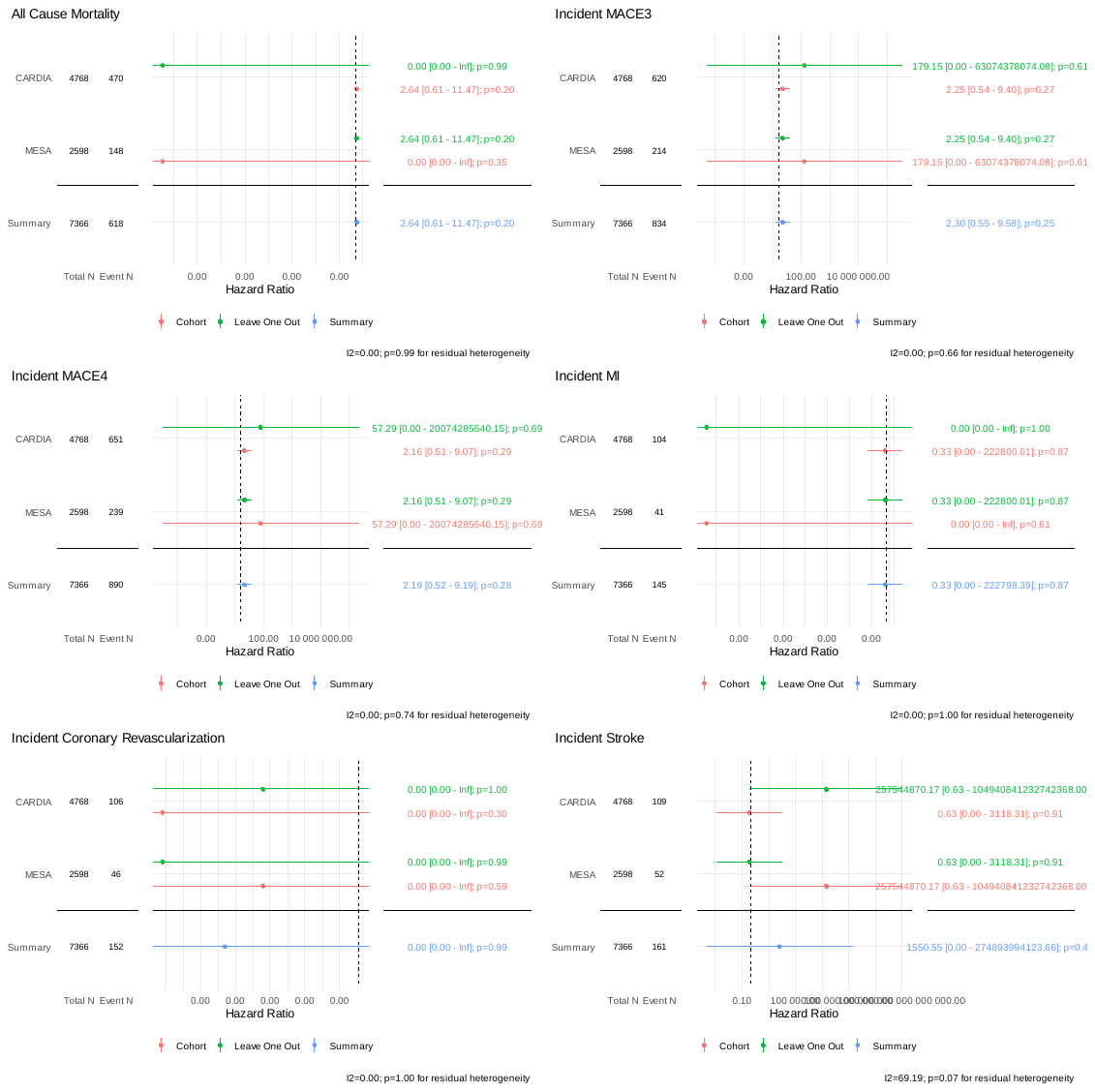

### Immature Granulocyte Count (Age/Sex/Race Adjusted)

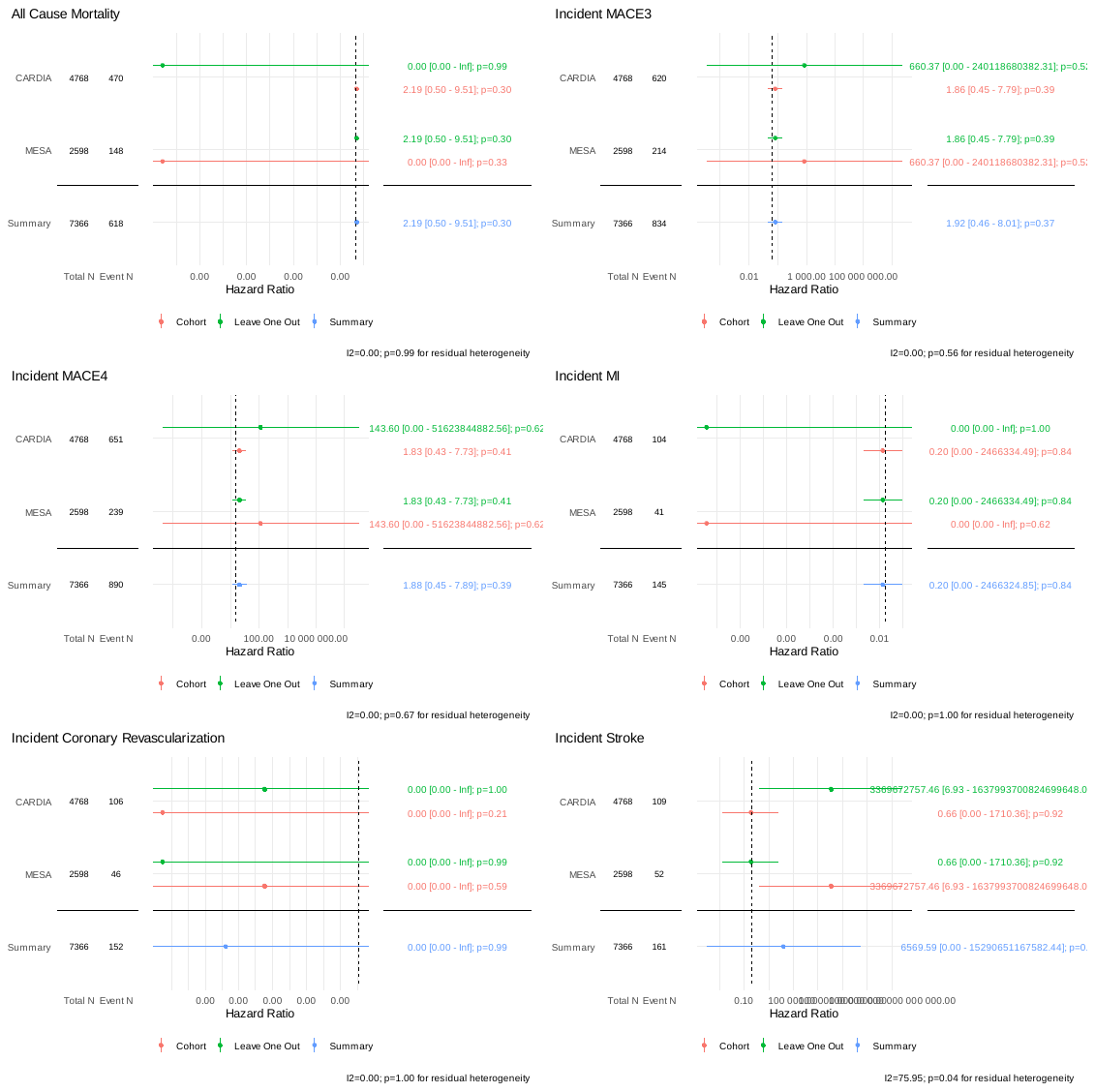

### Immature Granulocyte Count (Fully Adjusted)

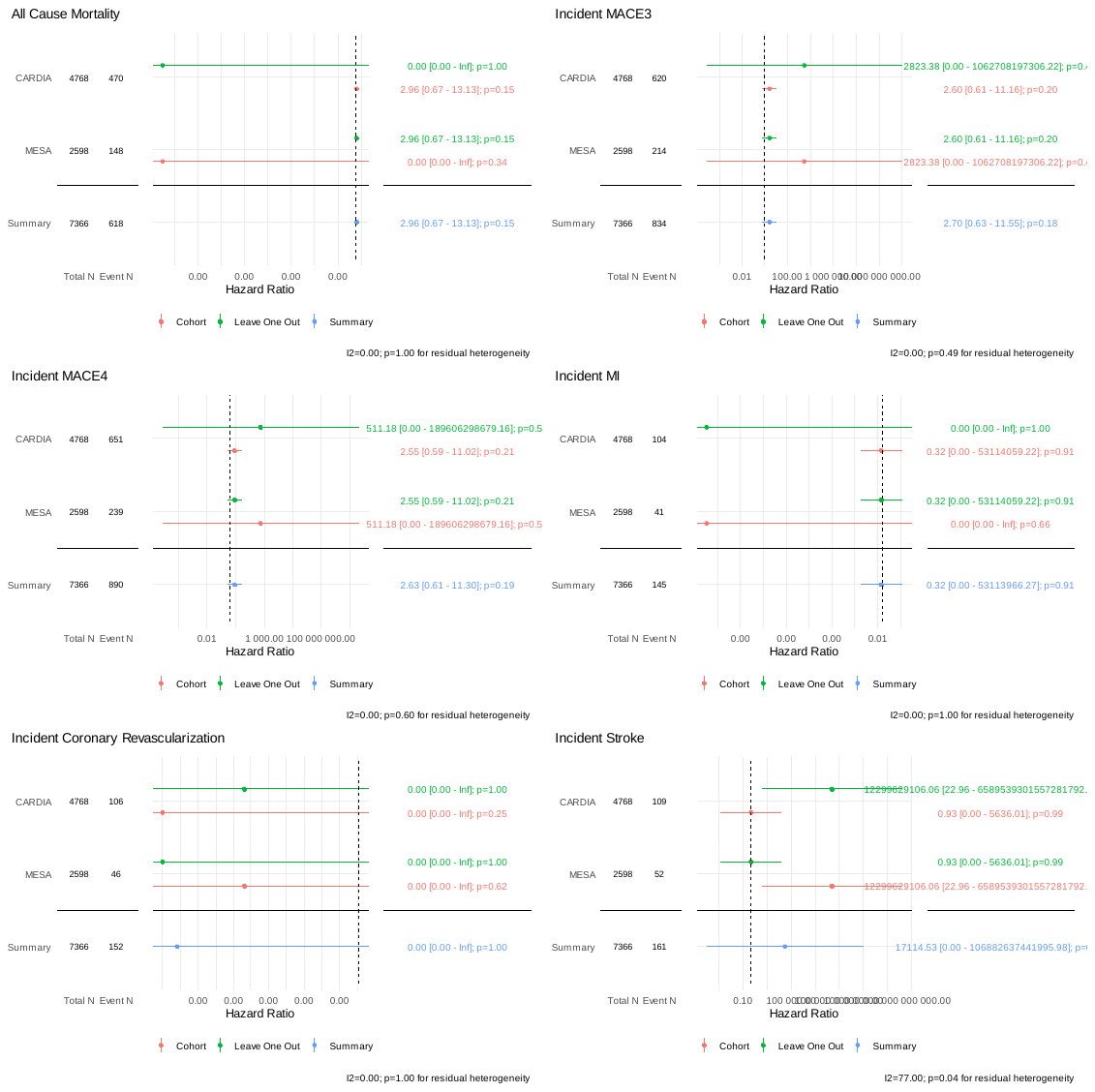

### Immature Granulocytes (%) (Unadjusted)

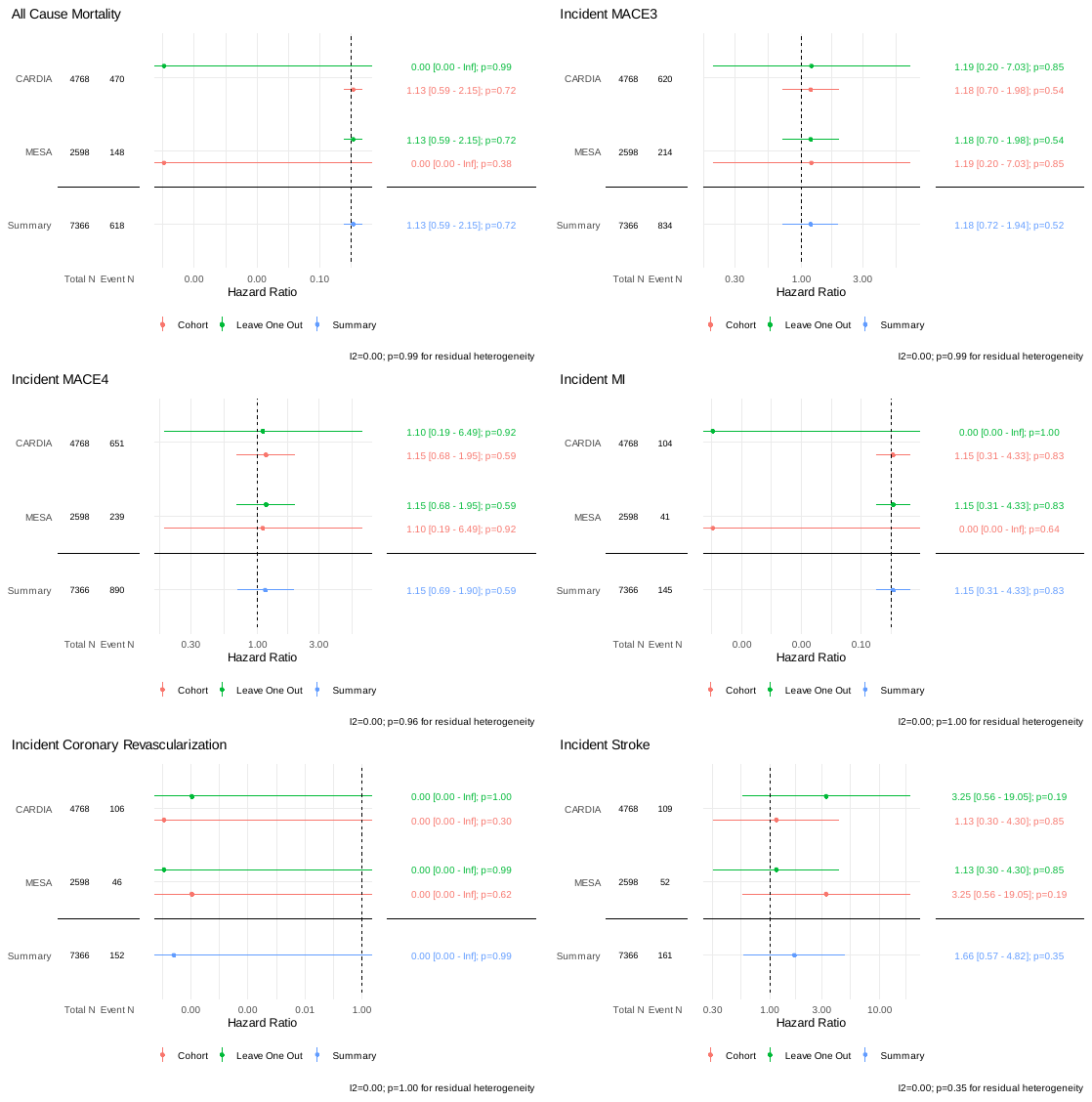

### Immature Granulocytes (%) (Age/Sex/Race Adjusted)

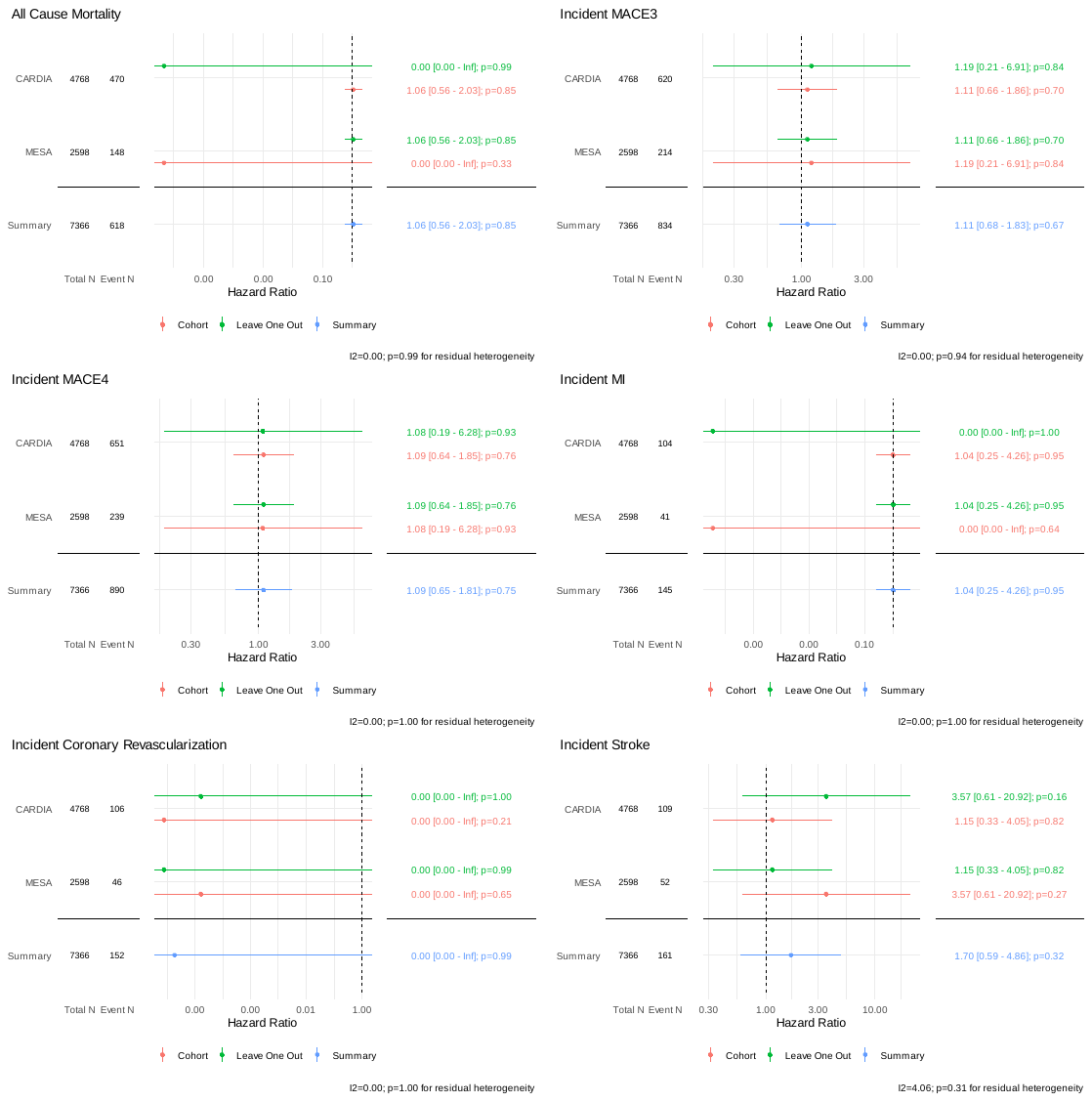

### Immature Granulocytes (%) (Fully Adjusted)

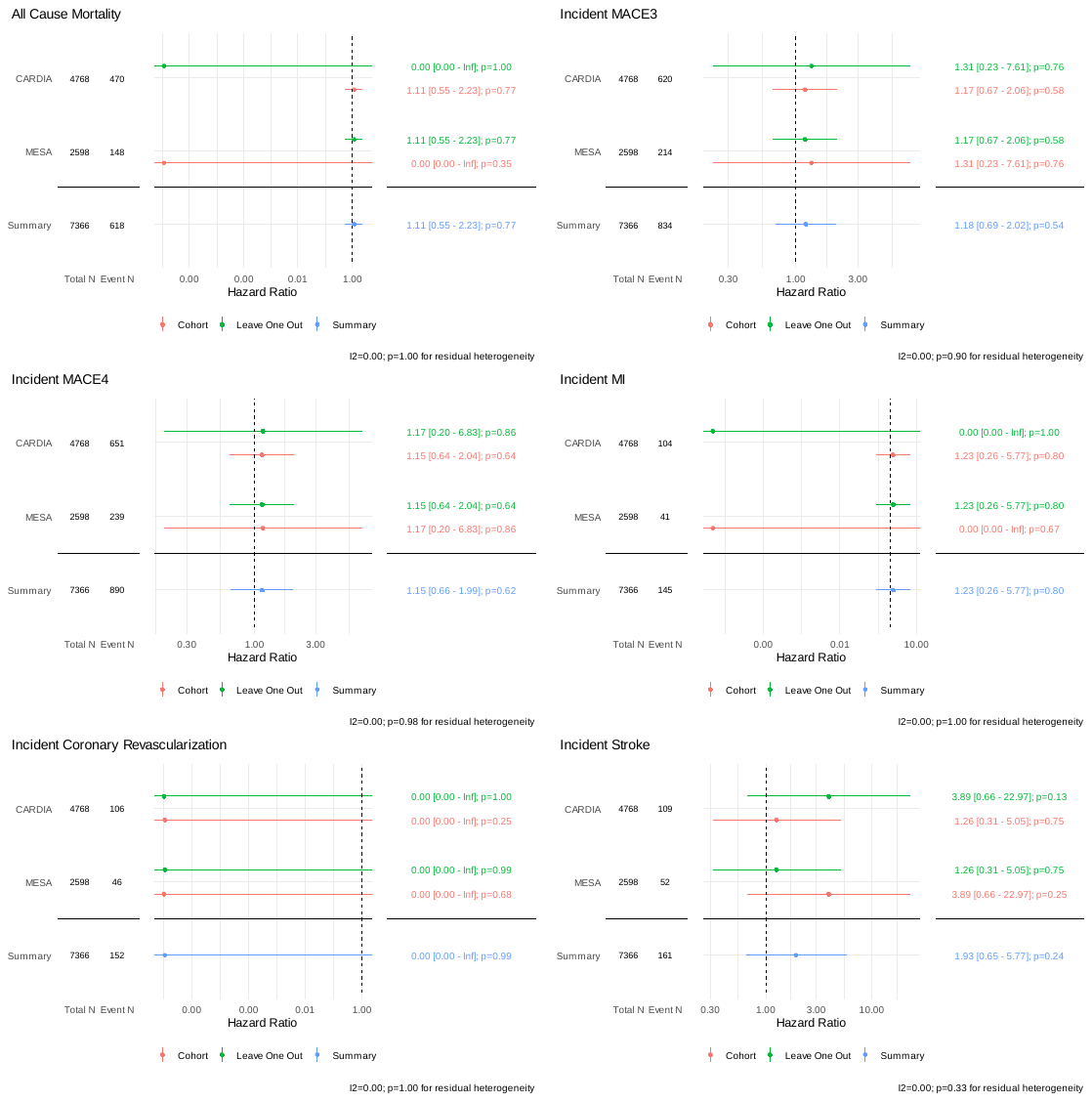

### Hematocrit (Unadjusted)

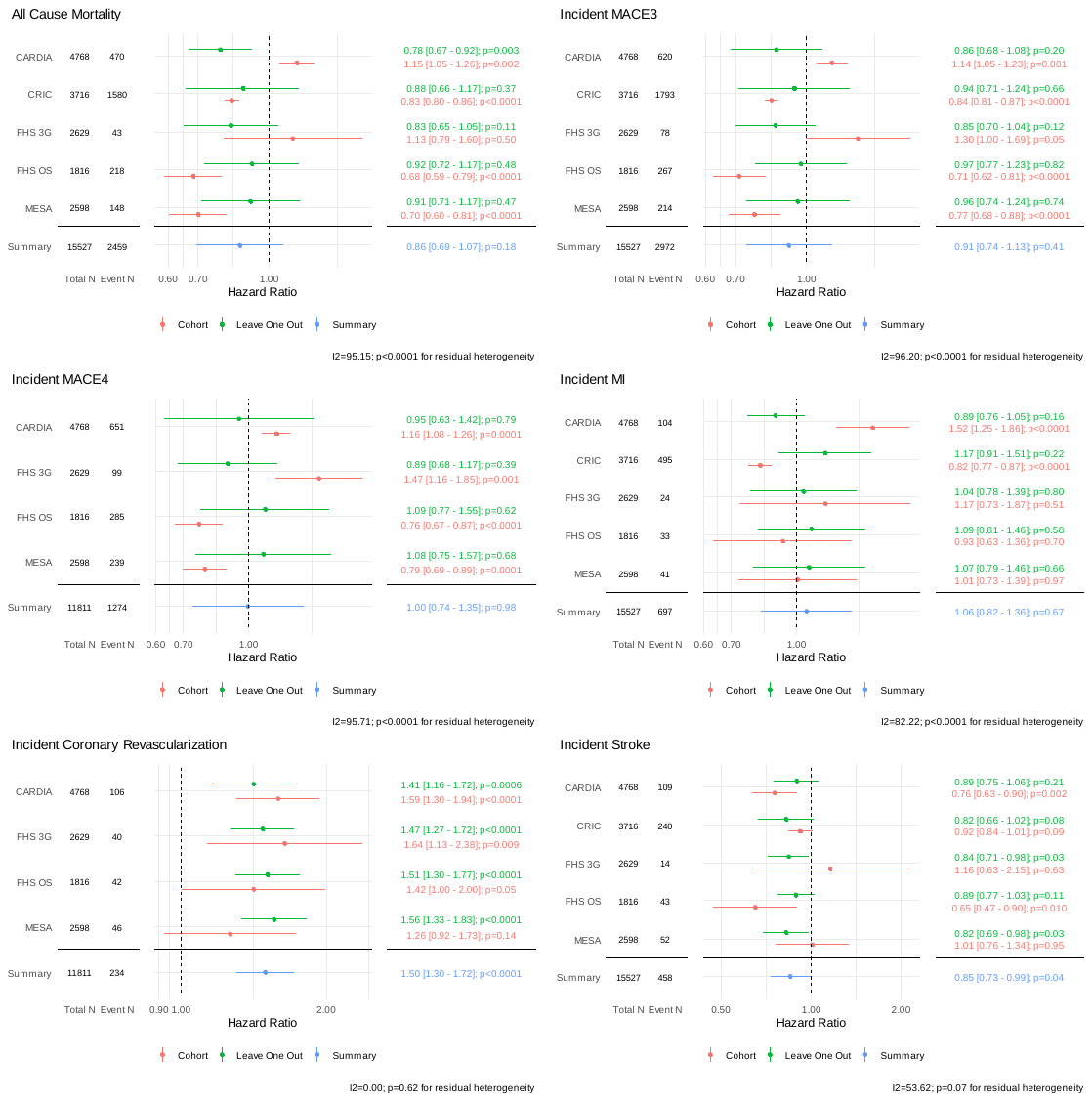

### Hematocrit (Age/Sex/Race Adjusted)

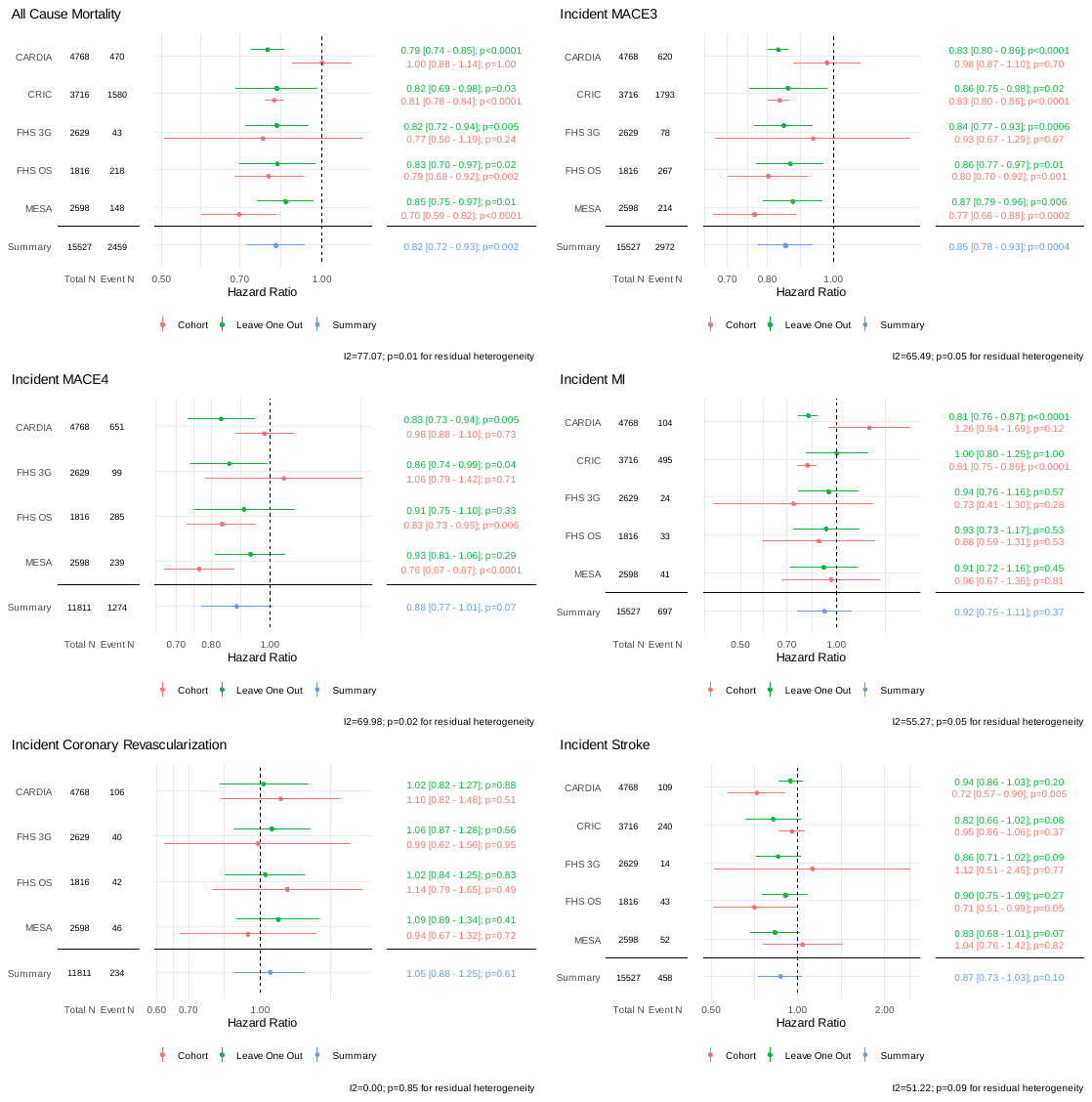

### Hematocrit (Fully Adjusted)

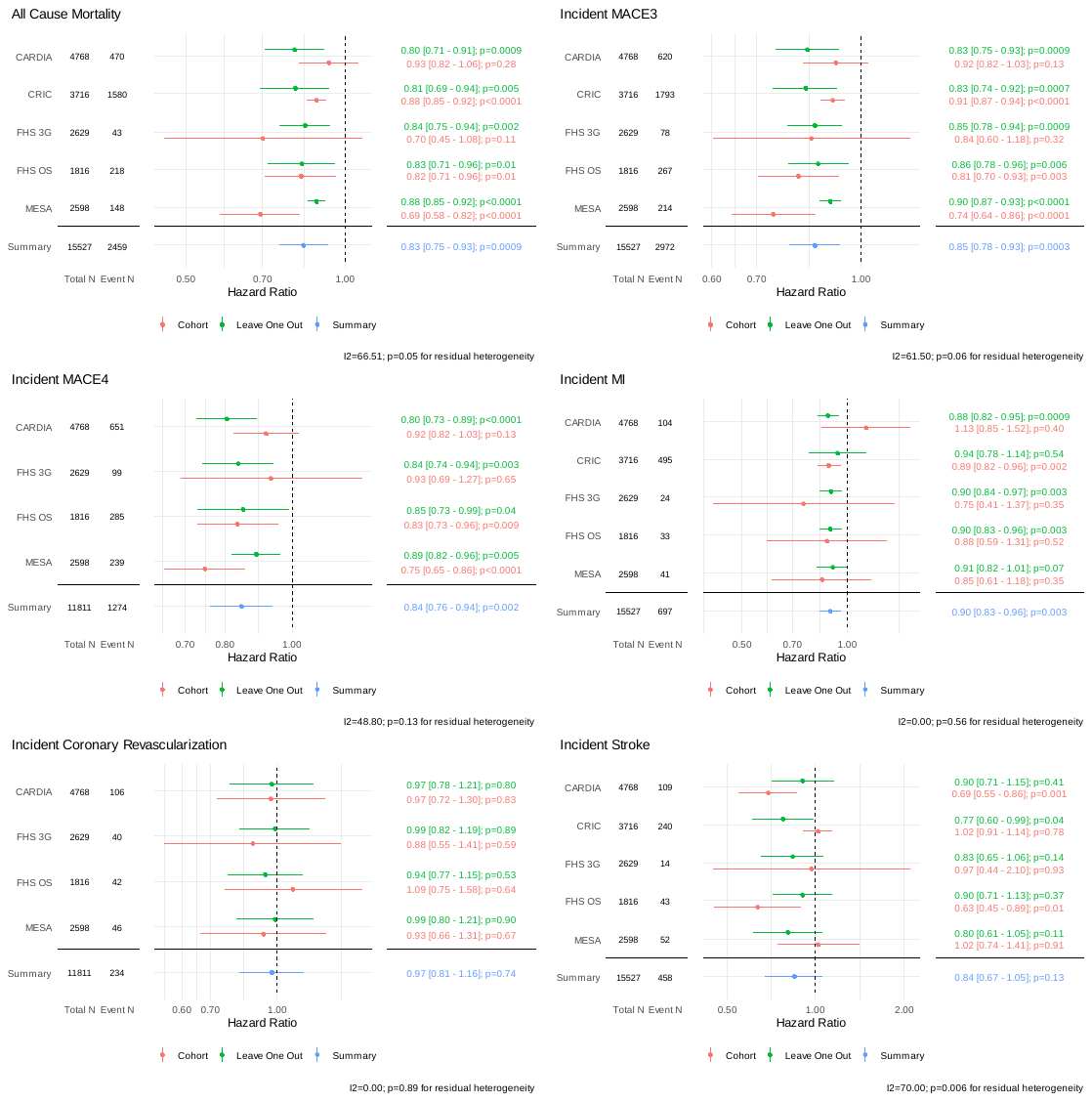

### Hematocrit (Fully Adjusted w/ hsCRP)

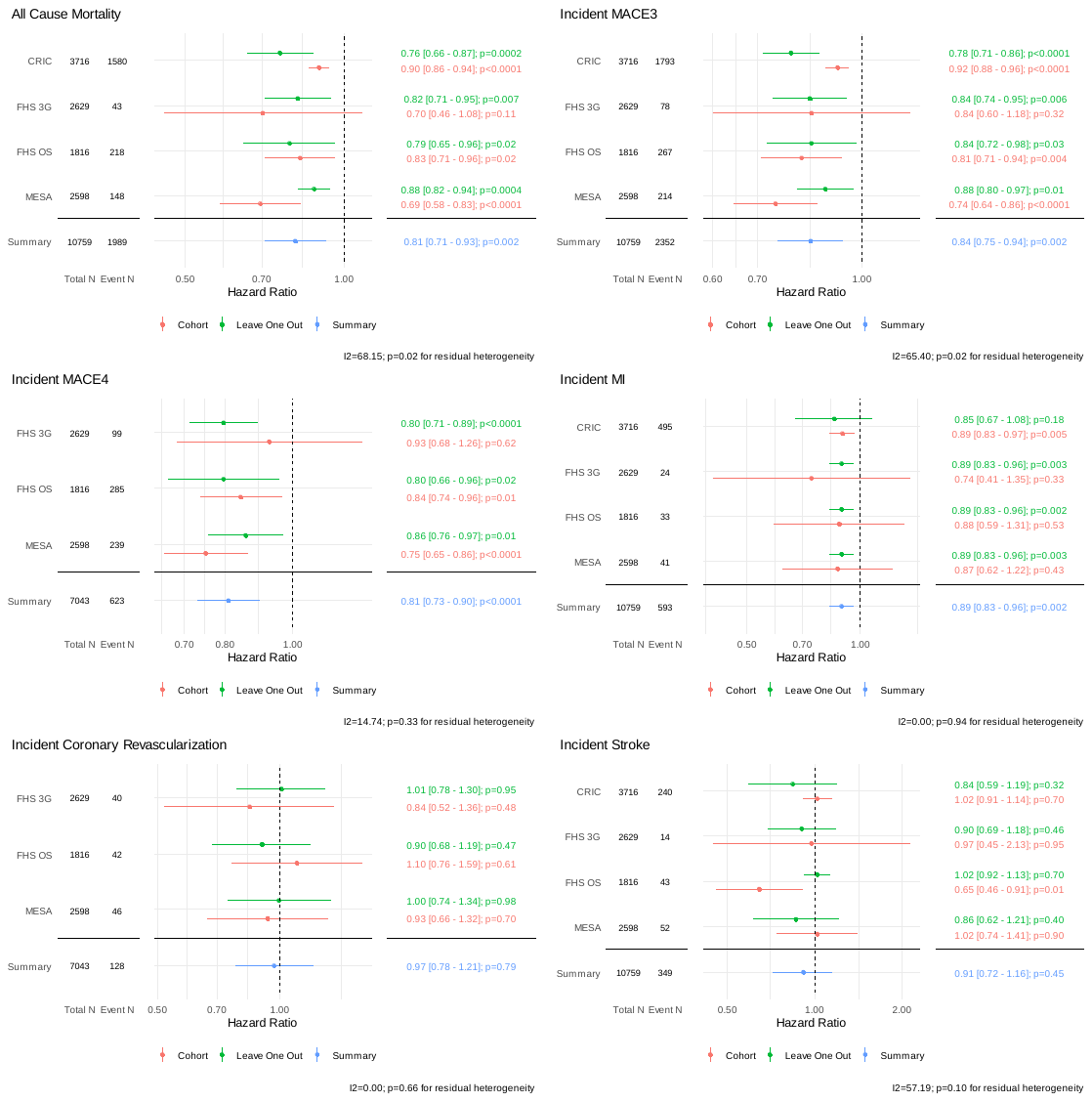

### Hemoglobin (Unadjusted)

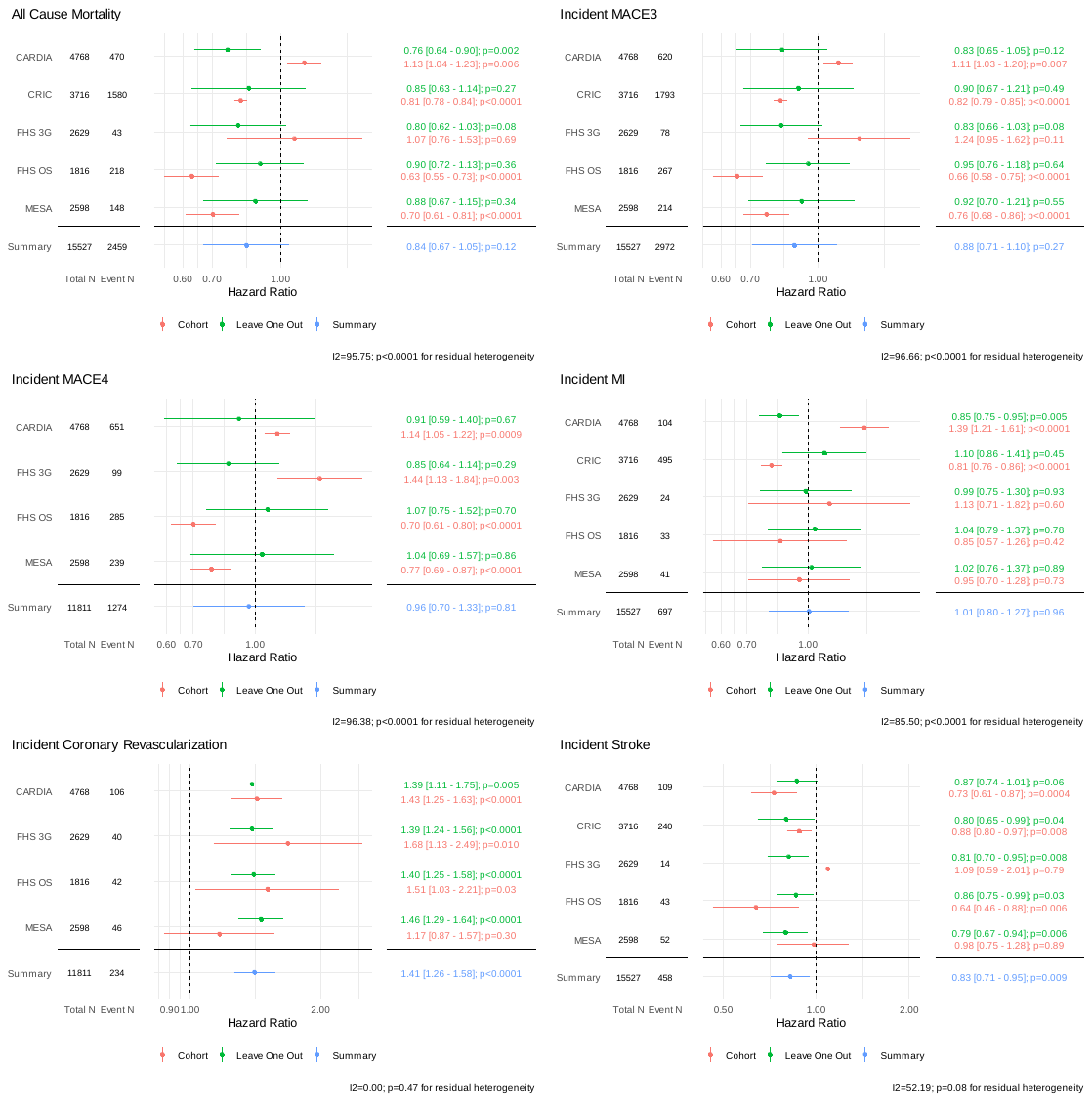

### Hemoglobin (Age/Sex/Race Adjusted)

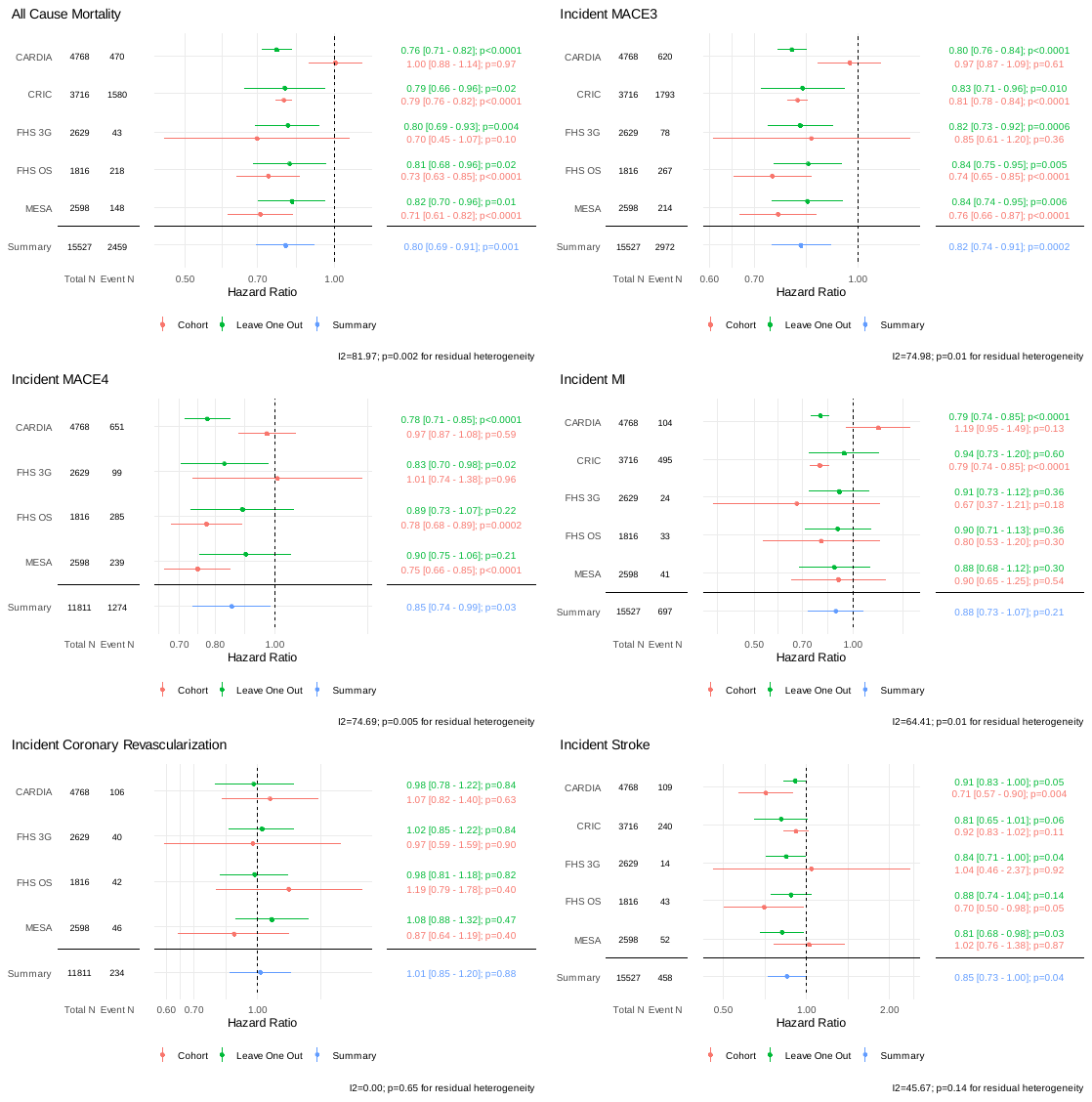

### Hemoglobin (Fully Adjusted)

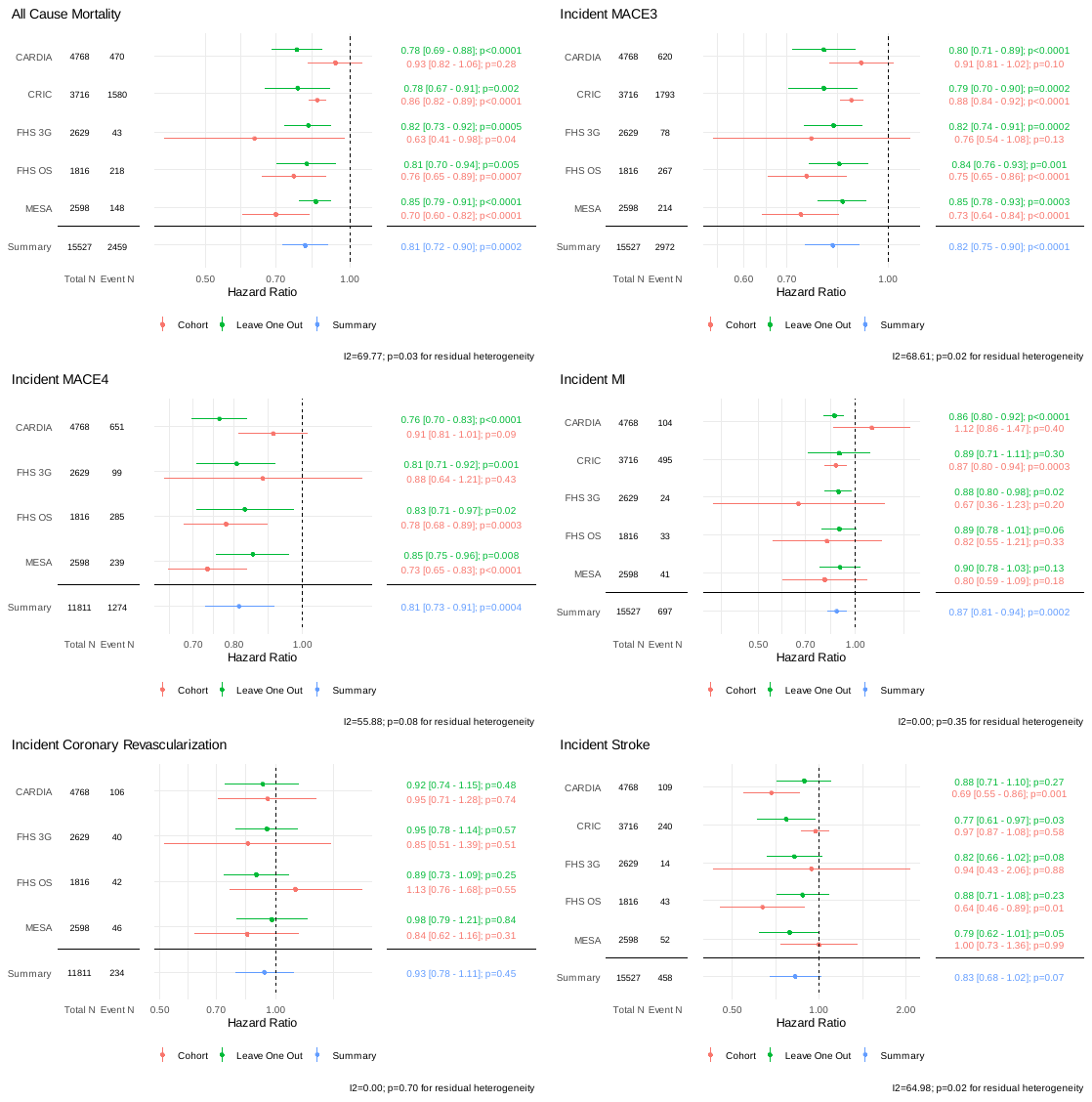

### Hemoglobin (Fully Adjusted w/ hsCRP)

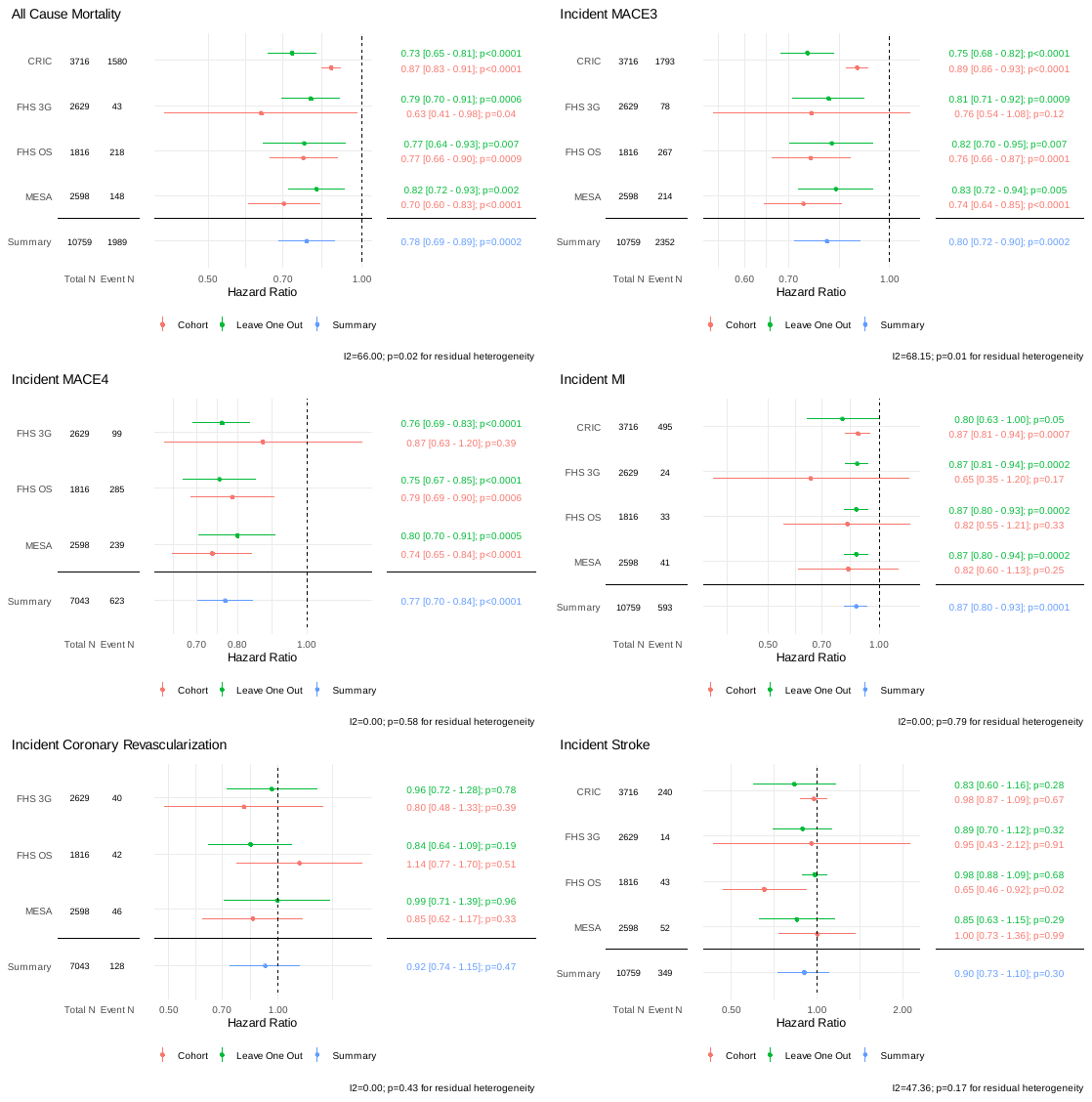

### Lymphocyte Count (Unadjusted)

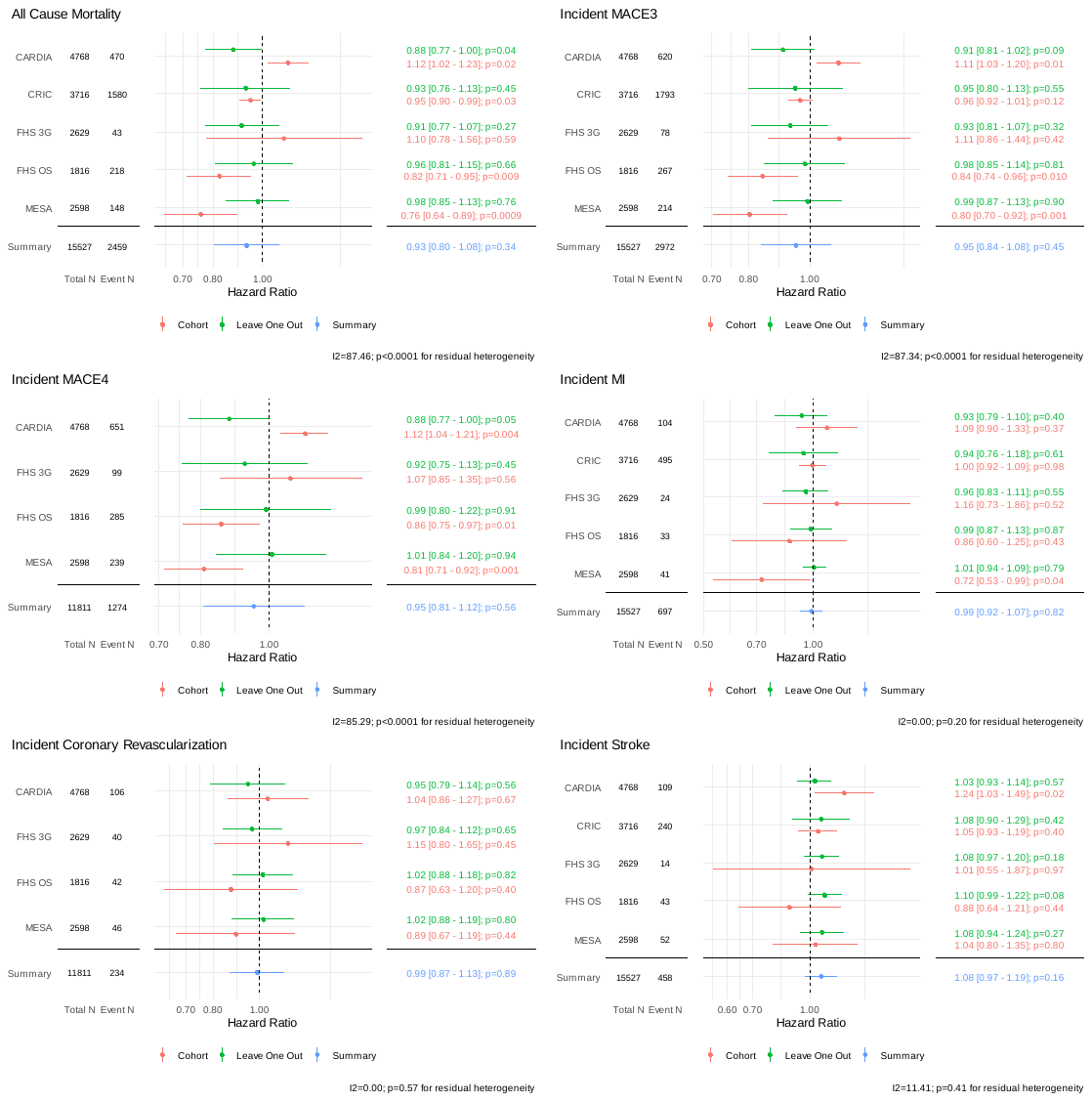

### Lymphocyte Count (Age/Sex/Race Adjusted)

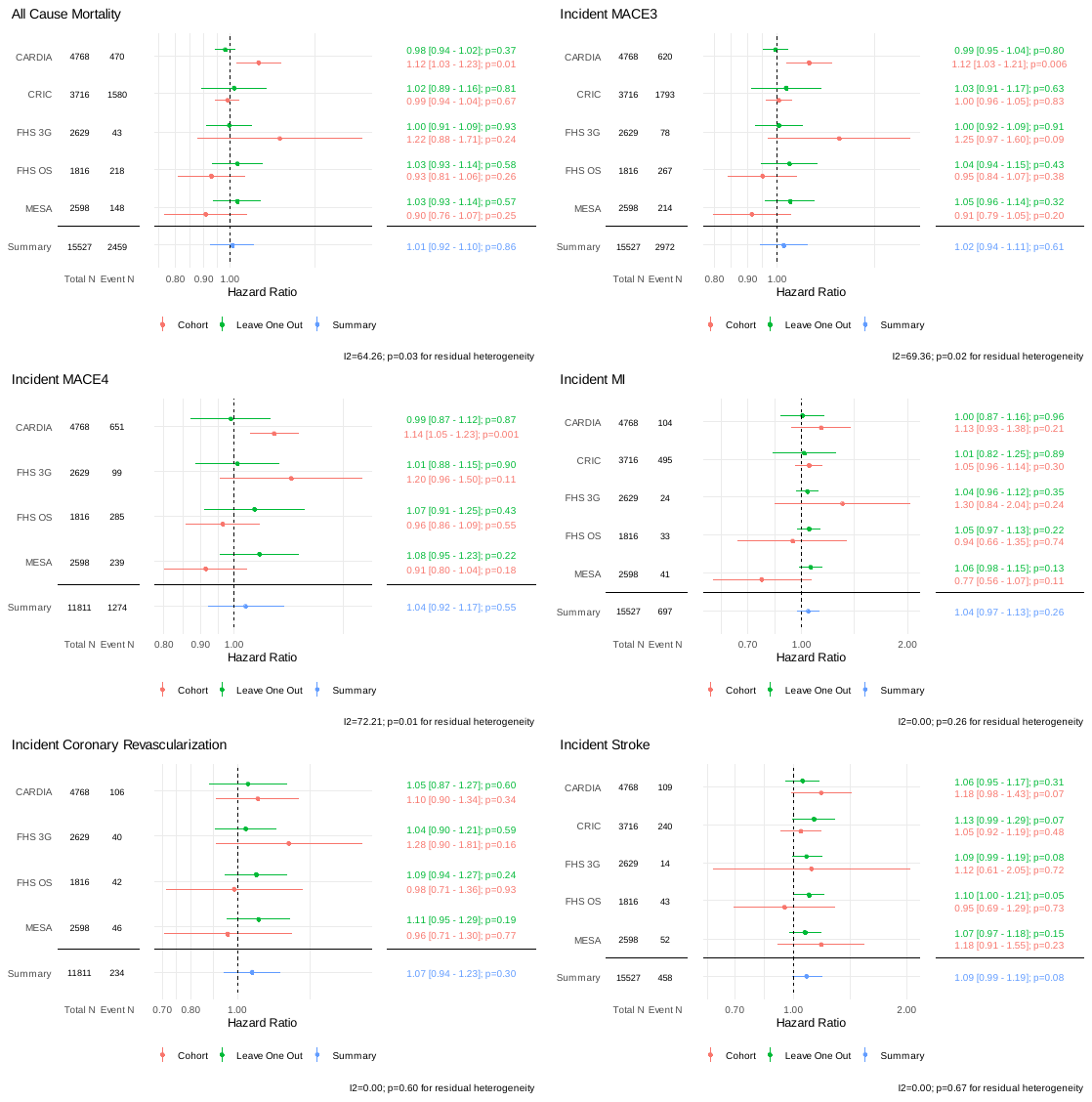

### Lymphocyte Count (Fully Adjusted)

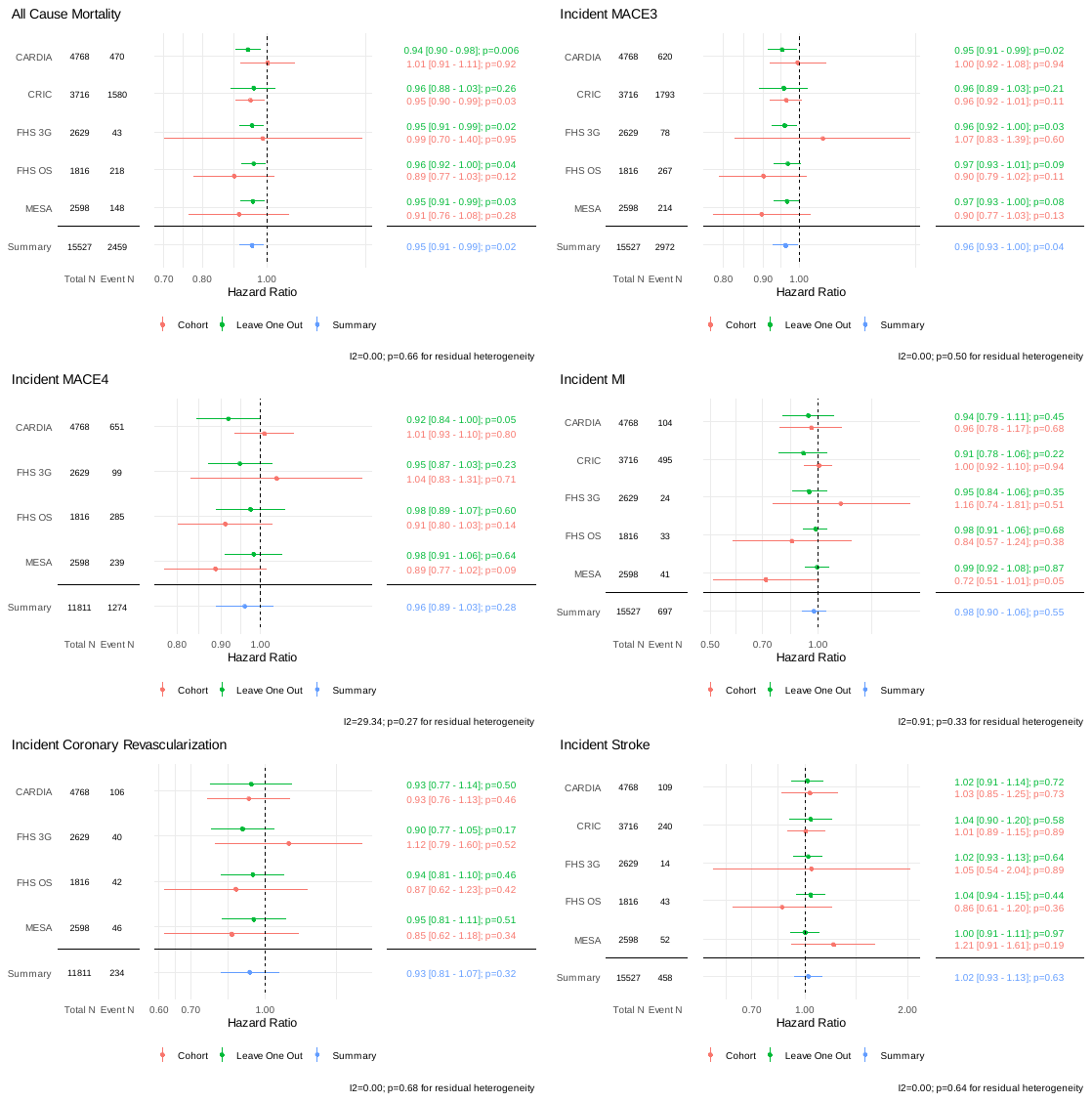

### Lymphocyte Count (Fully Adjusted w/ hsCRP)

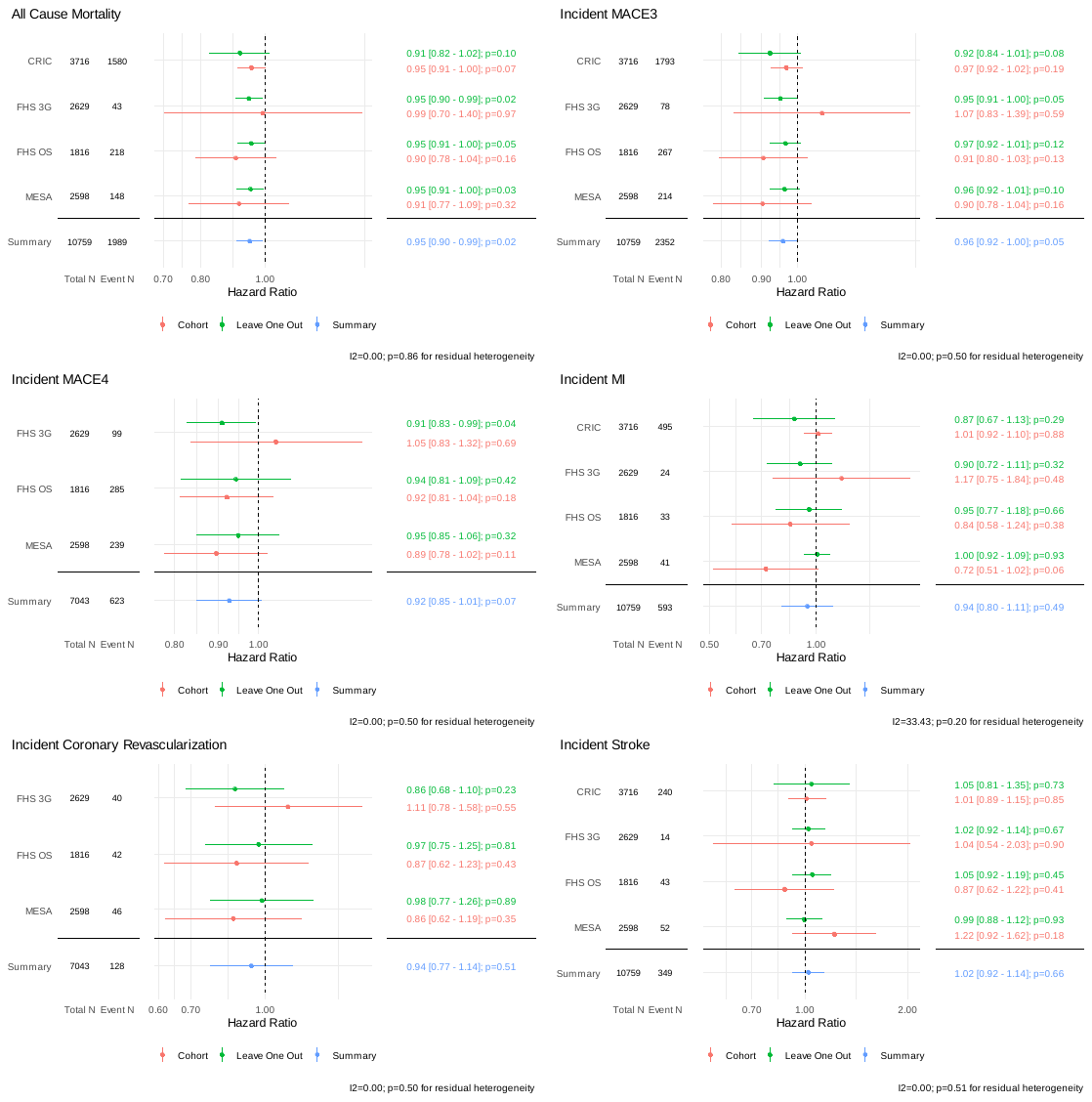

### Lymphocytes (%) (Unadjusted)

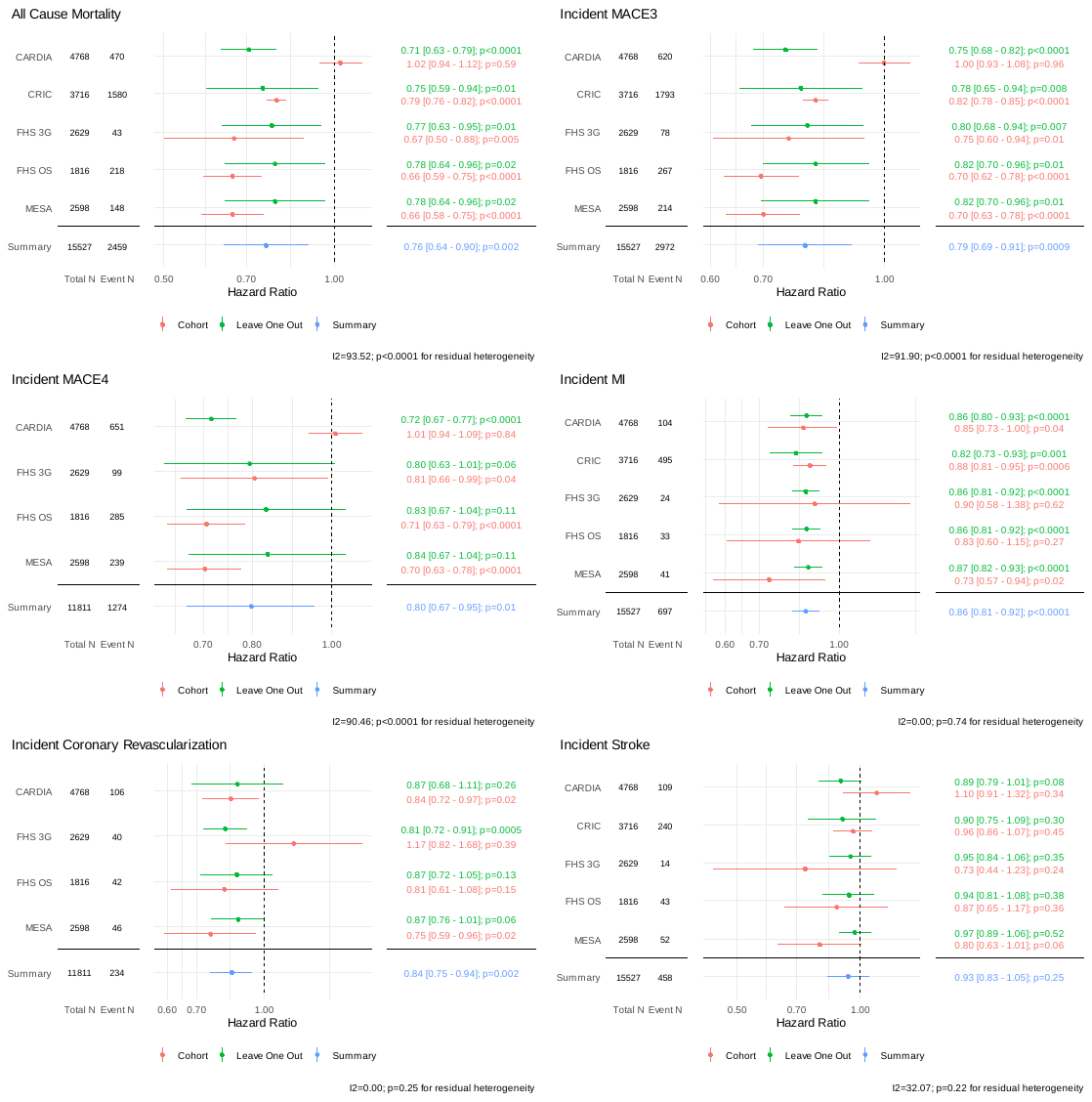

### Lymphocytes (%) (Age/Sex/Race Adjusted)

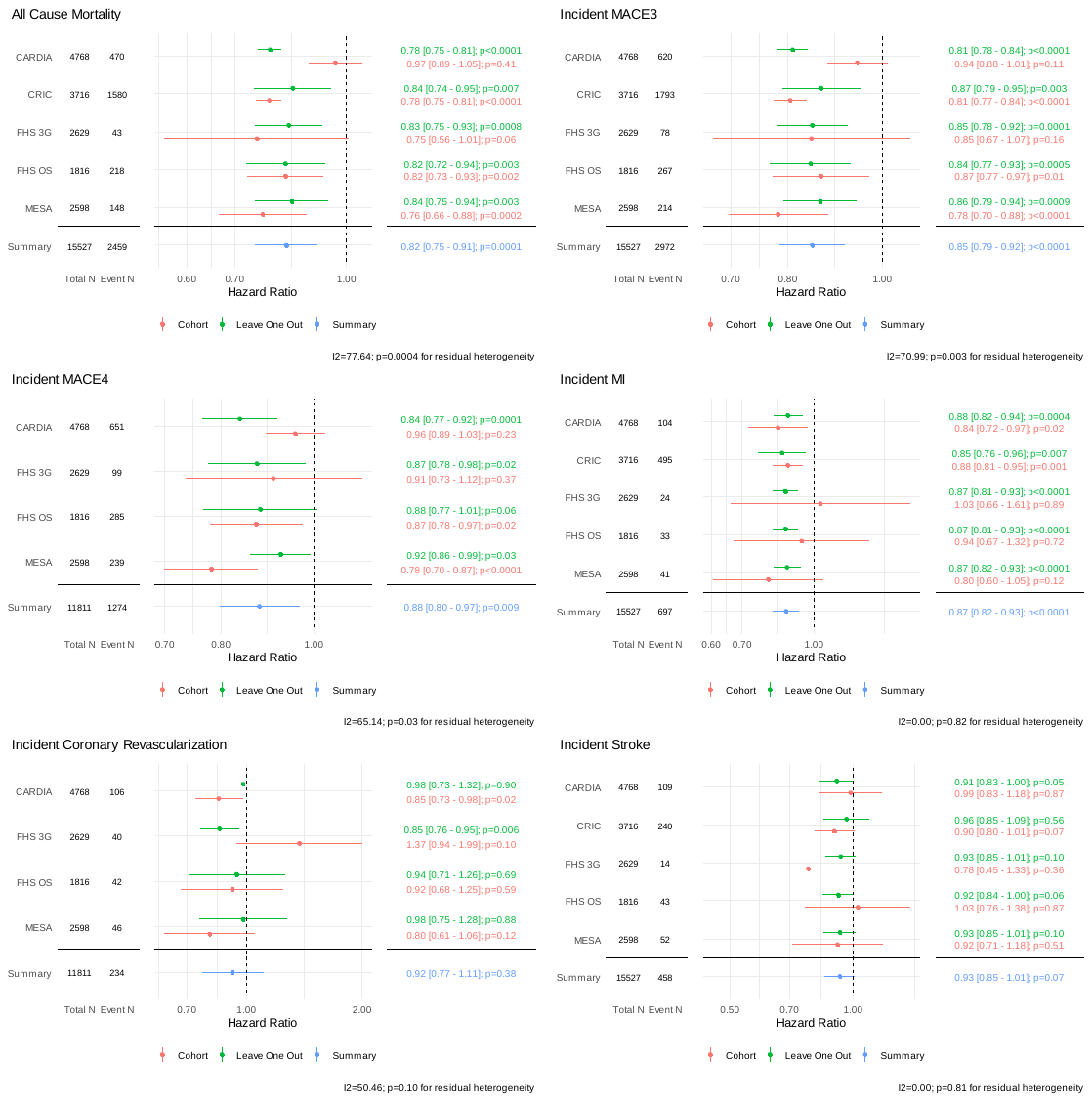

### Lymphocytes (%) (Fully Adjusted)

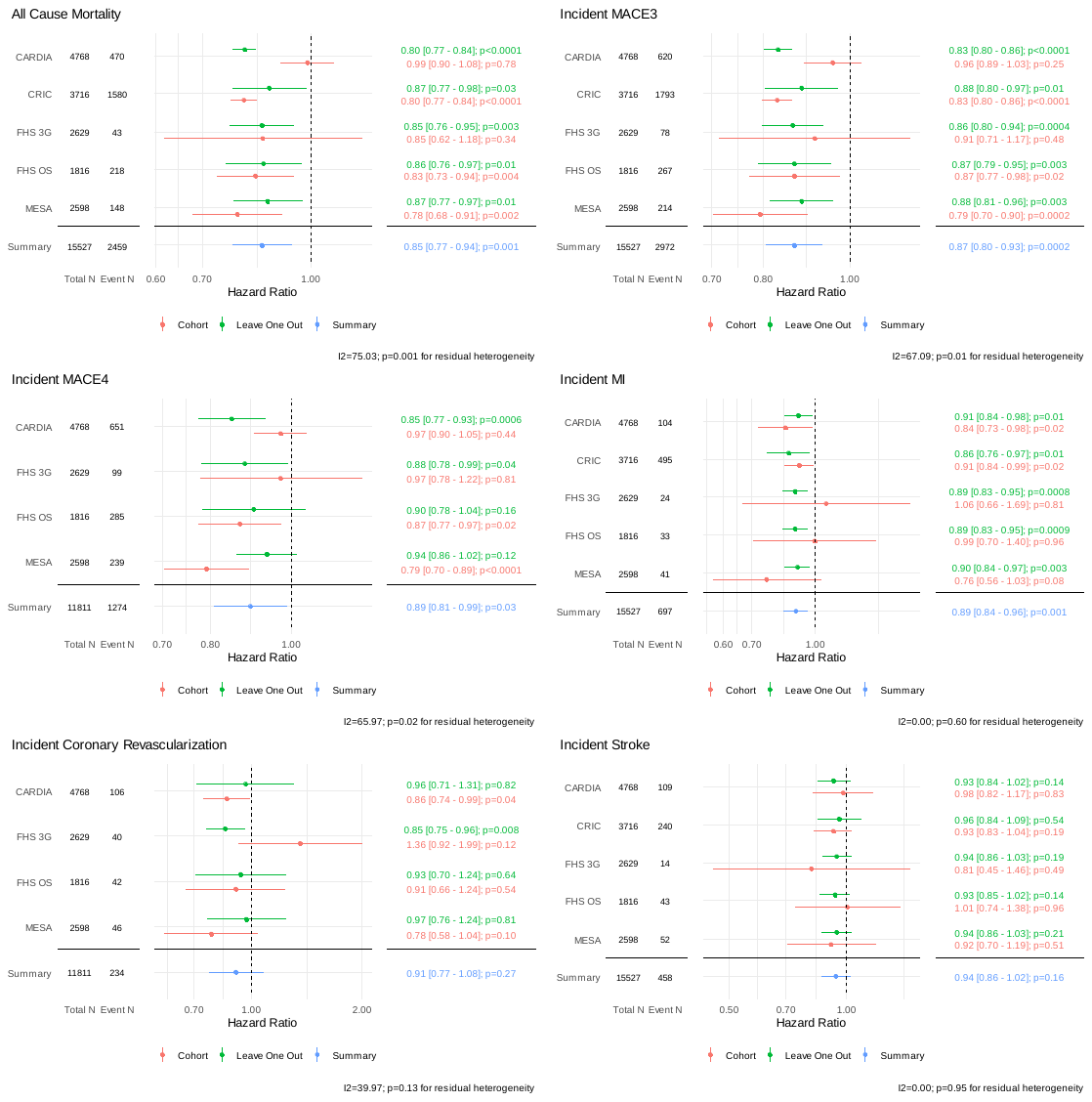

### Lymphocytes (%) (Fully Adjusted w/ hsCRP)

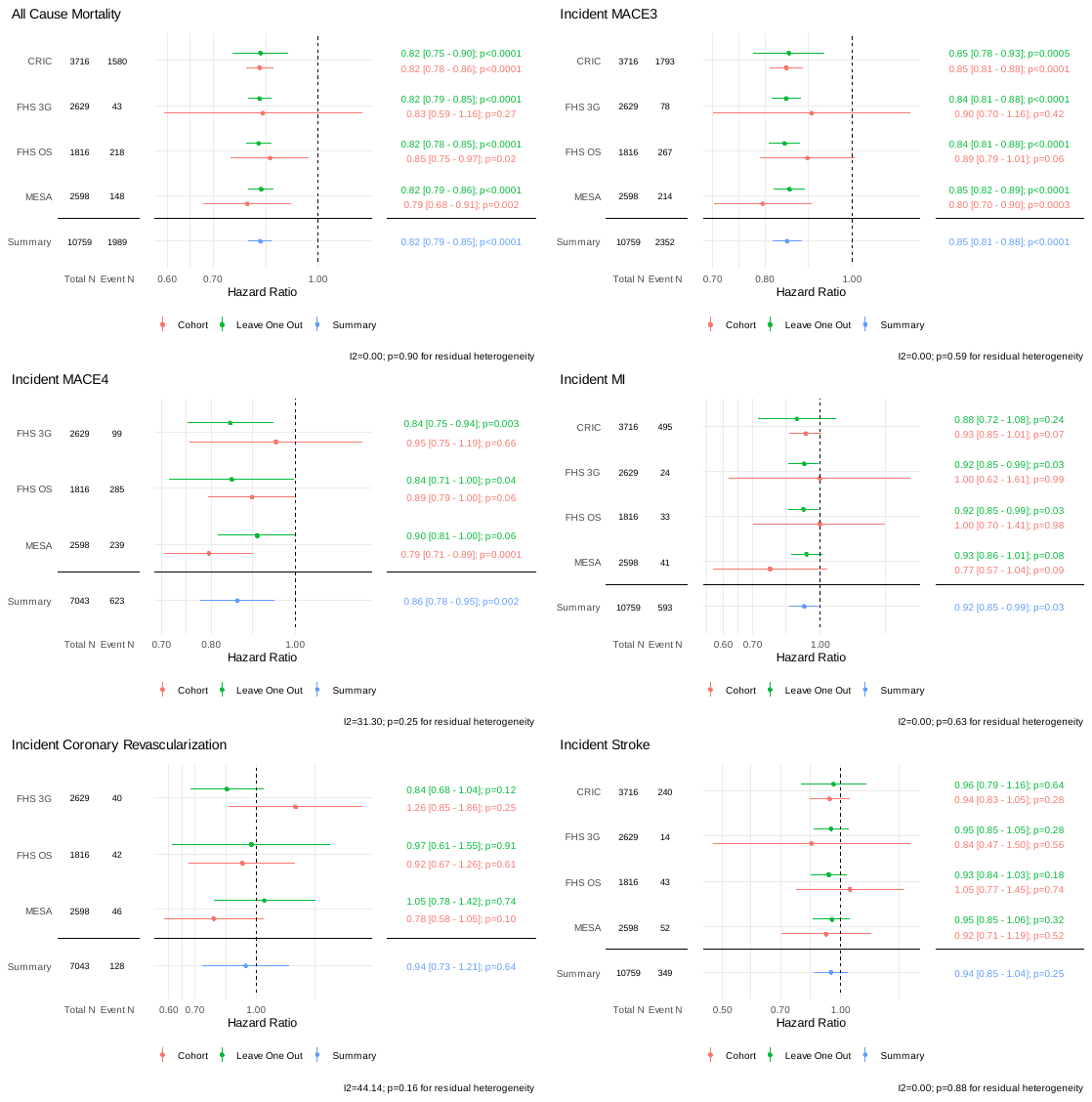

### Mean Corpuscular Hemoglobin (Unadjusted)

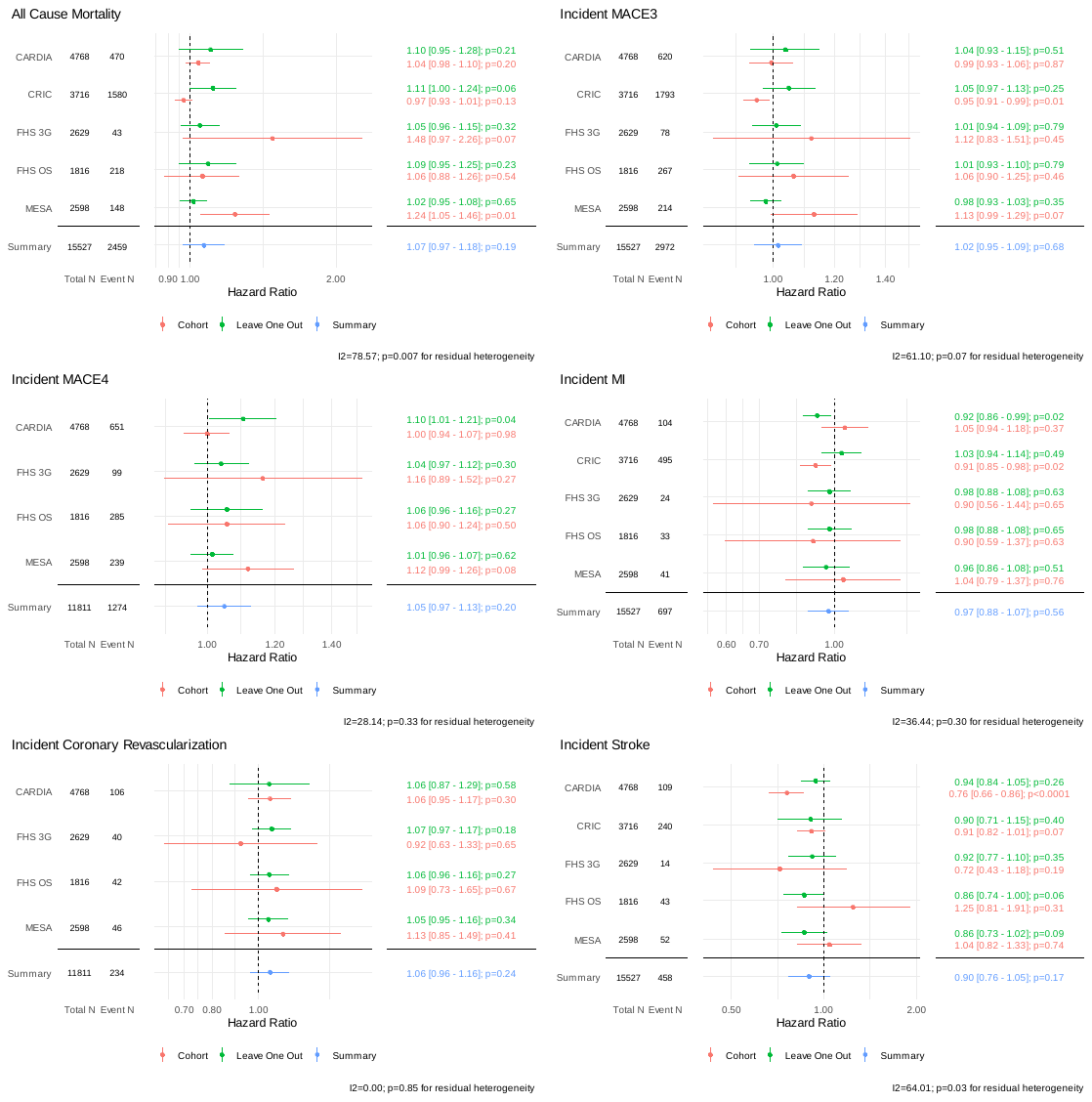

### Mean Corpuscular Hemoglobin (Age/Sex/Race Adjusted)

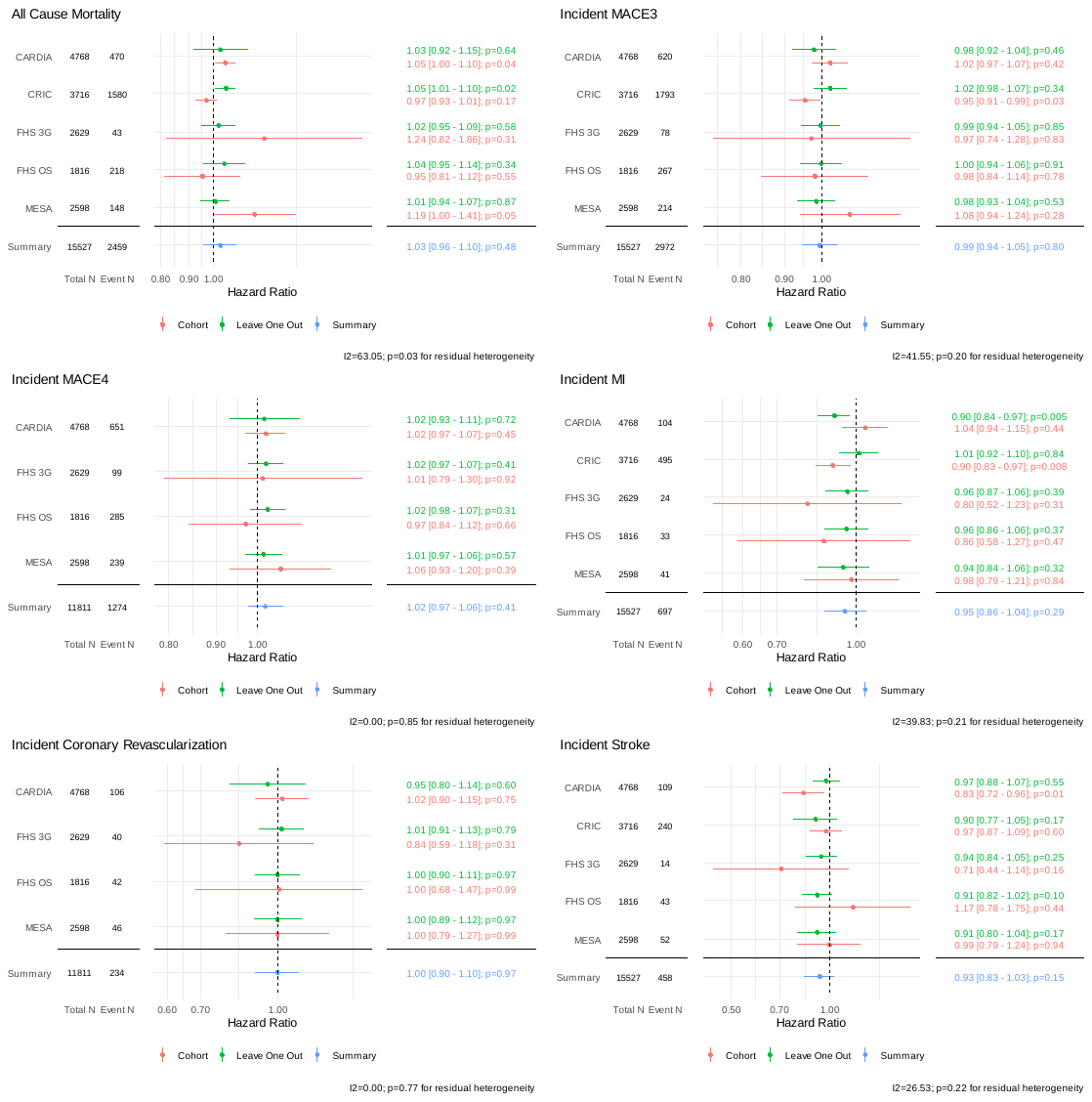

### Mean Corpuscular Hemoglobin (Fully Adjusted)

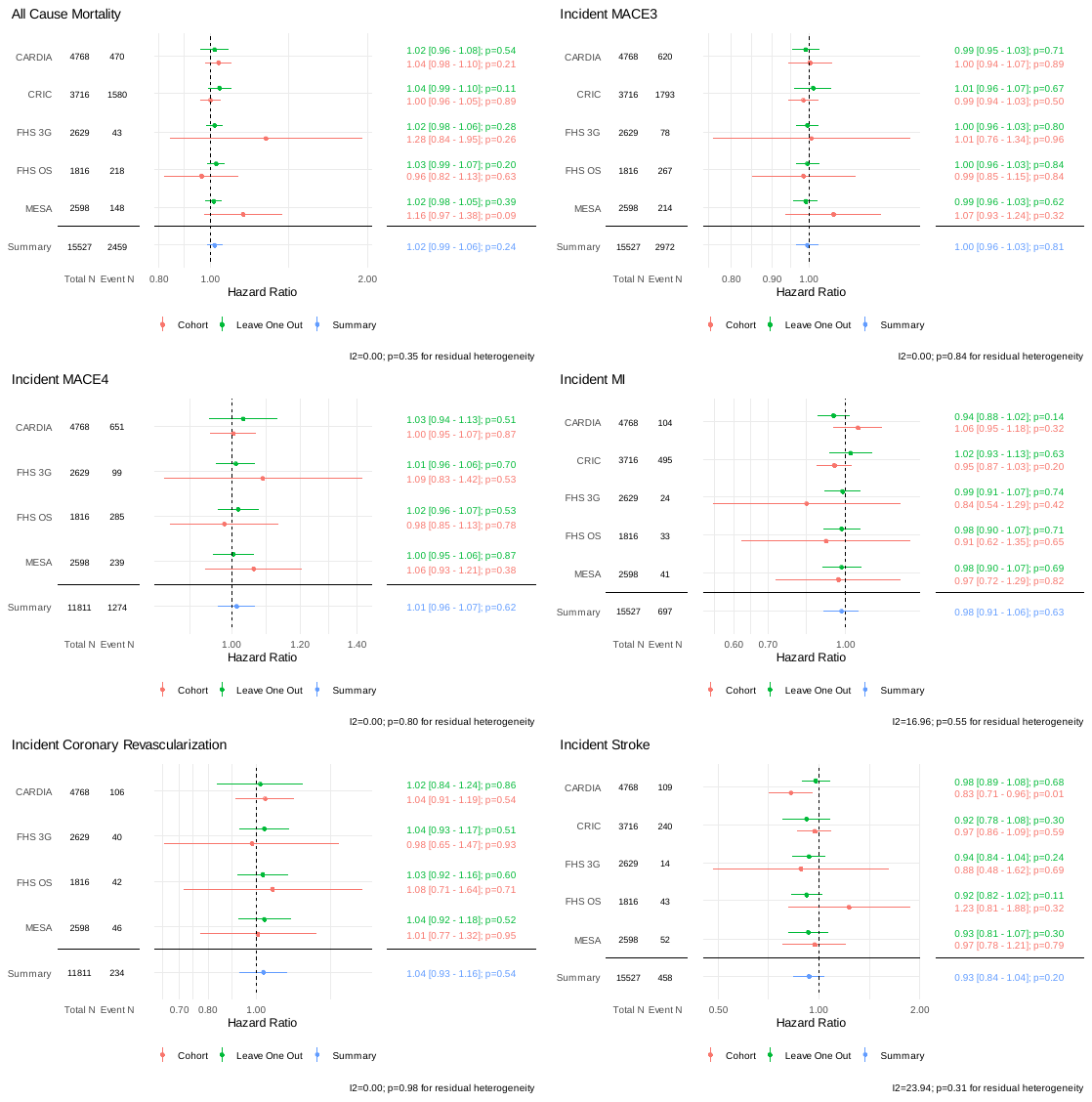

### Mean Corpuscular Hemoglobin (Fully Adjusted w/ hsCRP)

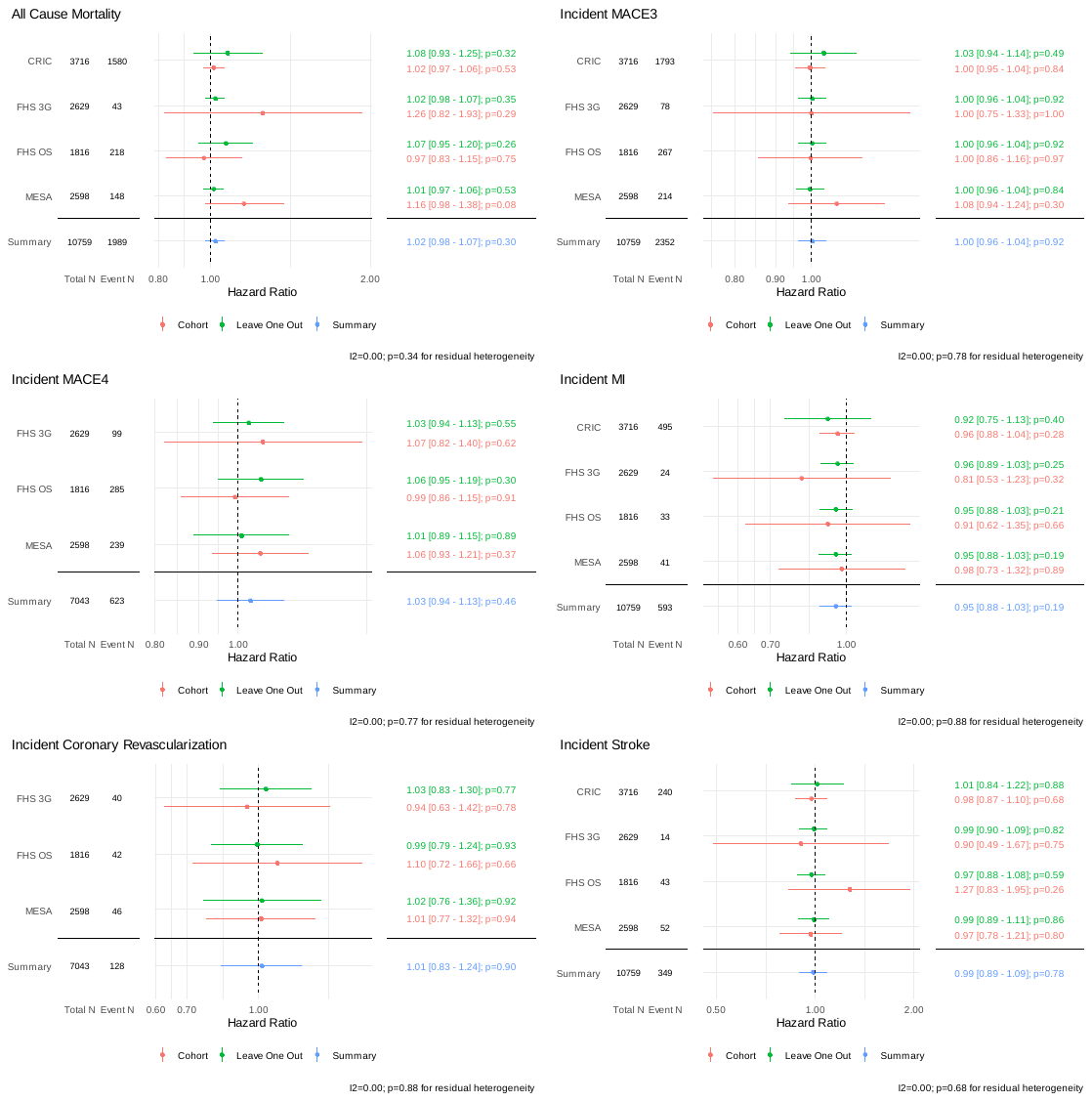

### Mean Corpuscular Hemoglobin Concentration (Unadjusted)

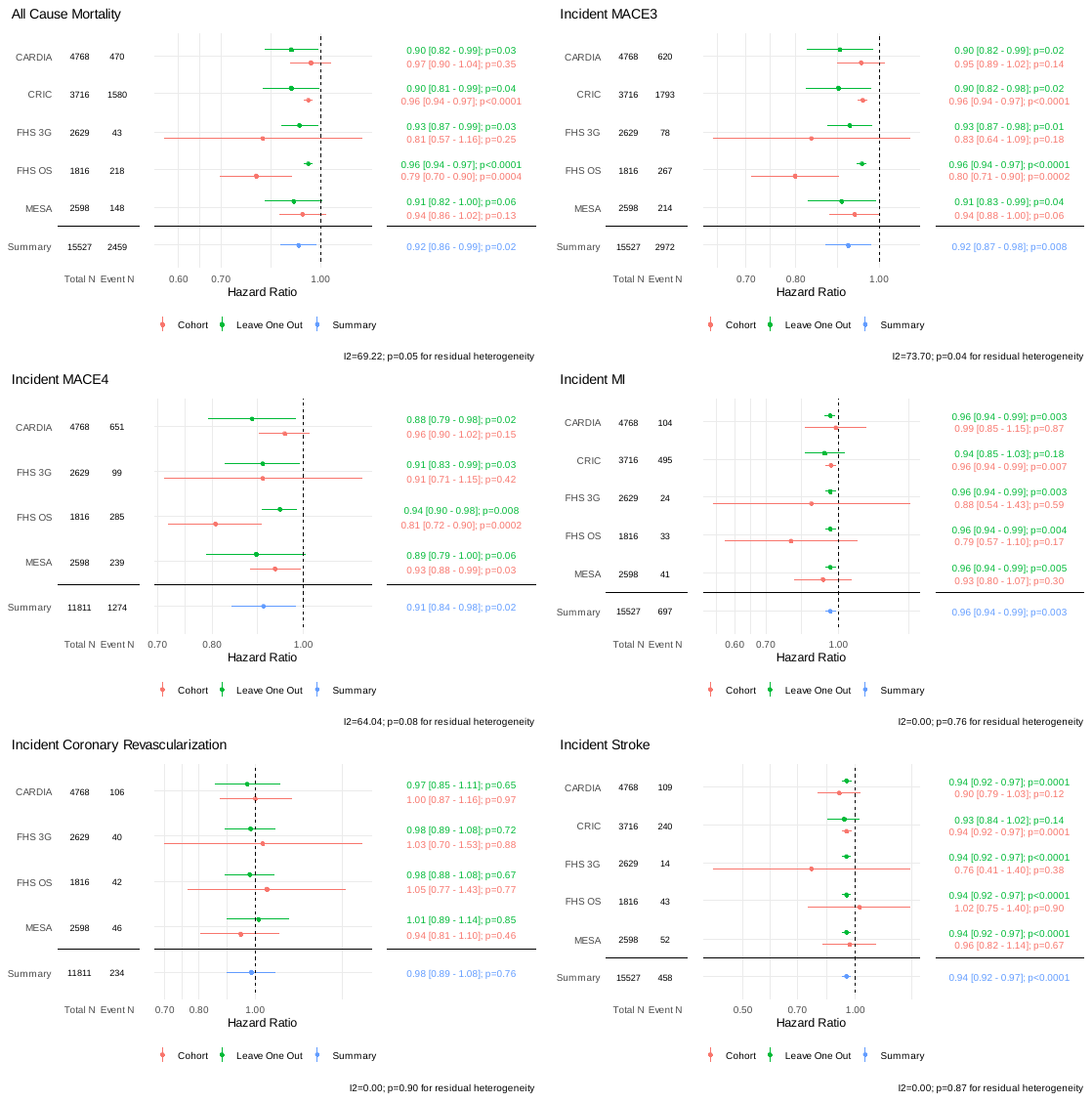

### Mean Corpuscular Hemoglobin Concentration (Age/Sex/Race Adjusted)

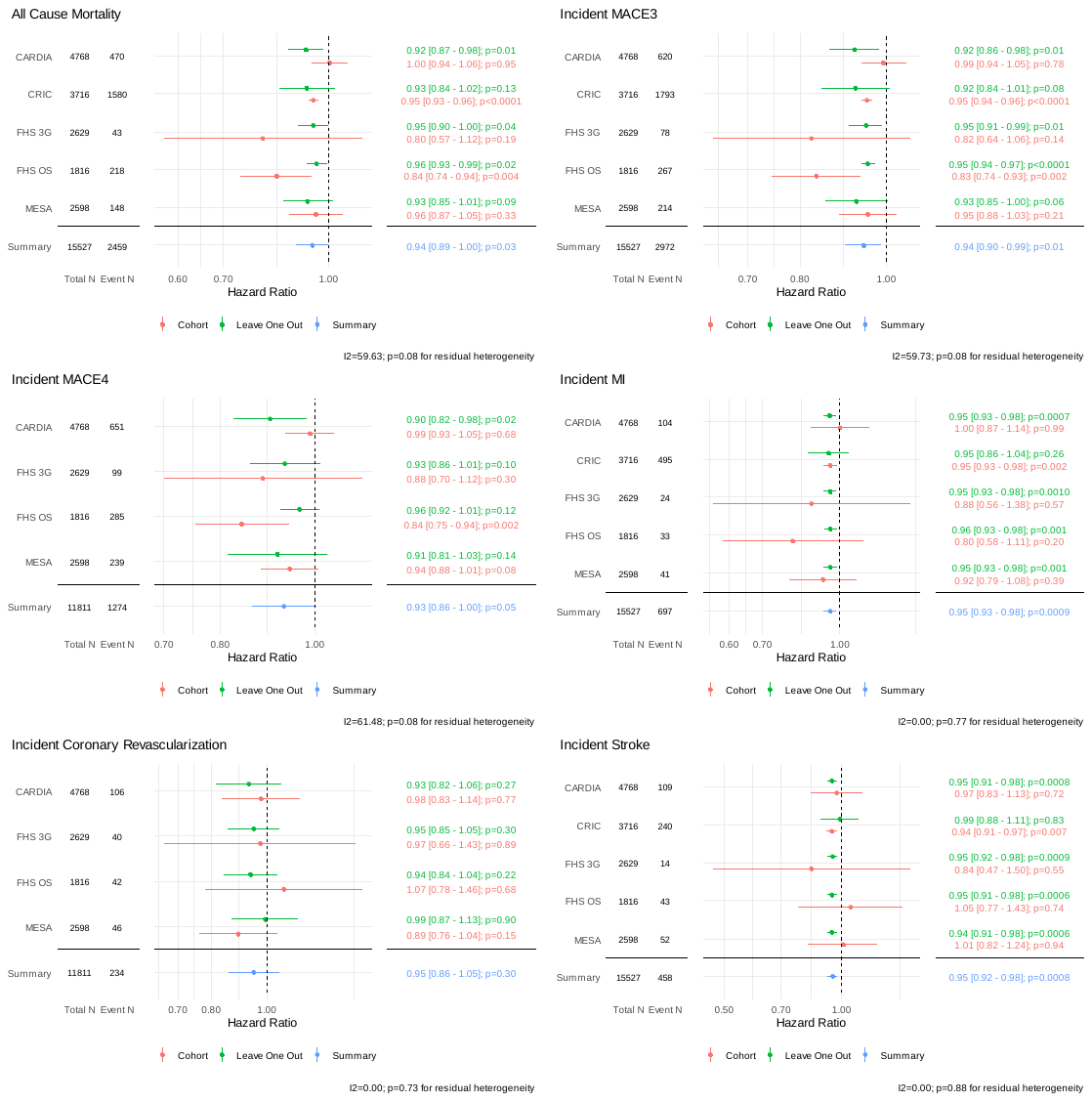

### Mean Corpuscular Hemoglobin Concentration (Fully Adjusted)

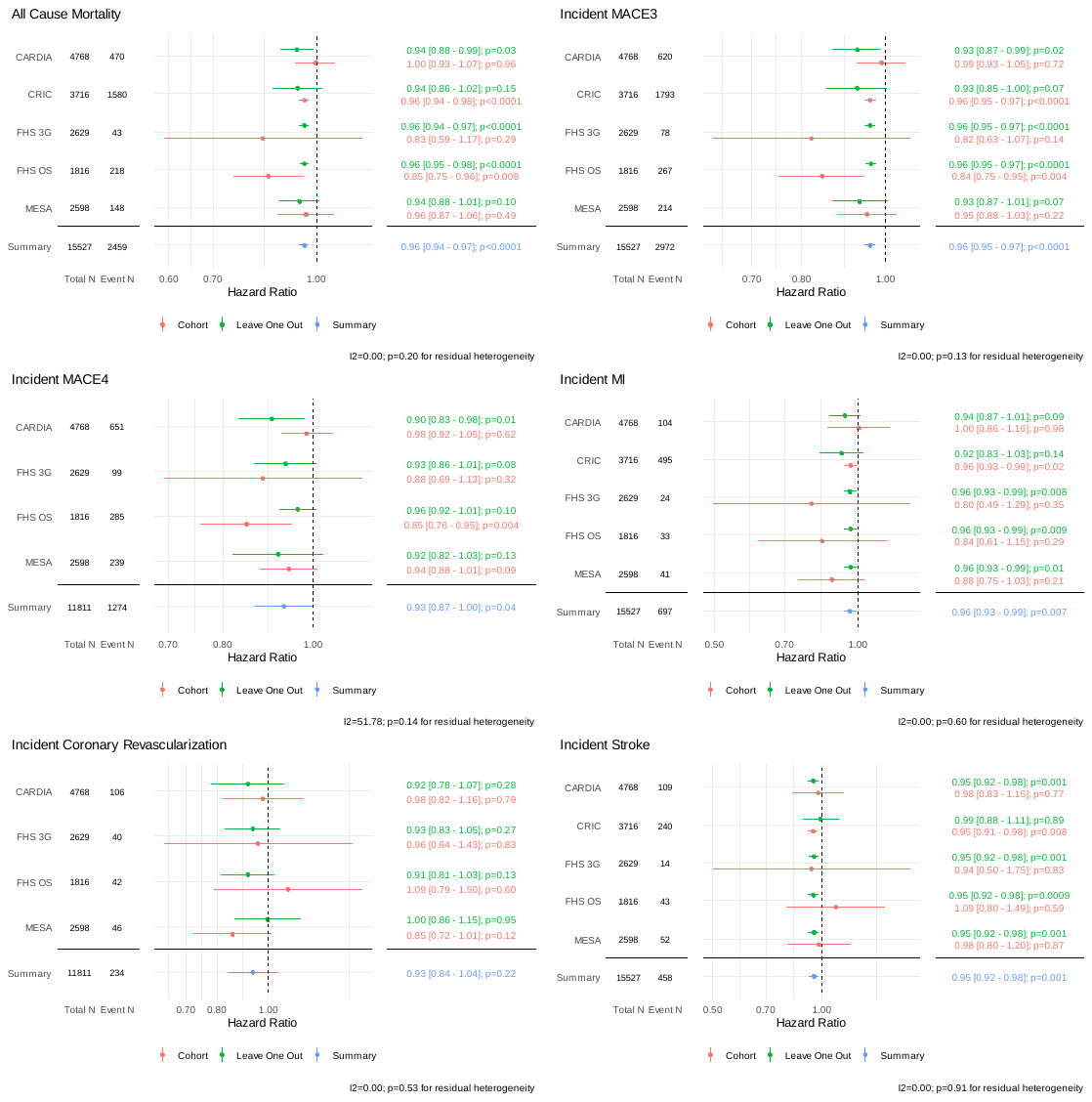

### Mean Corpuscular Hemoglobin Concentration (Fully Adjusted w/ hsCRP)

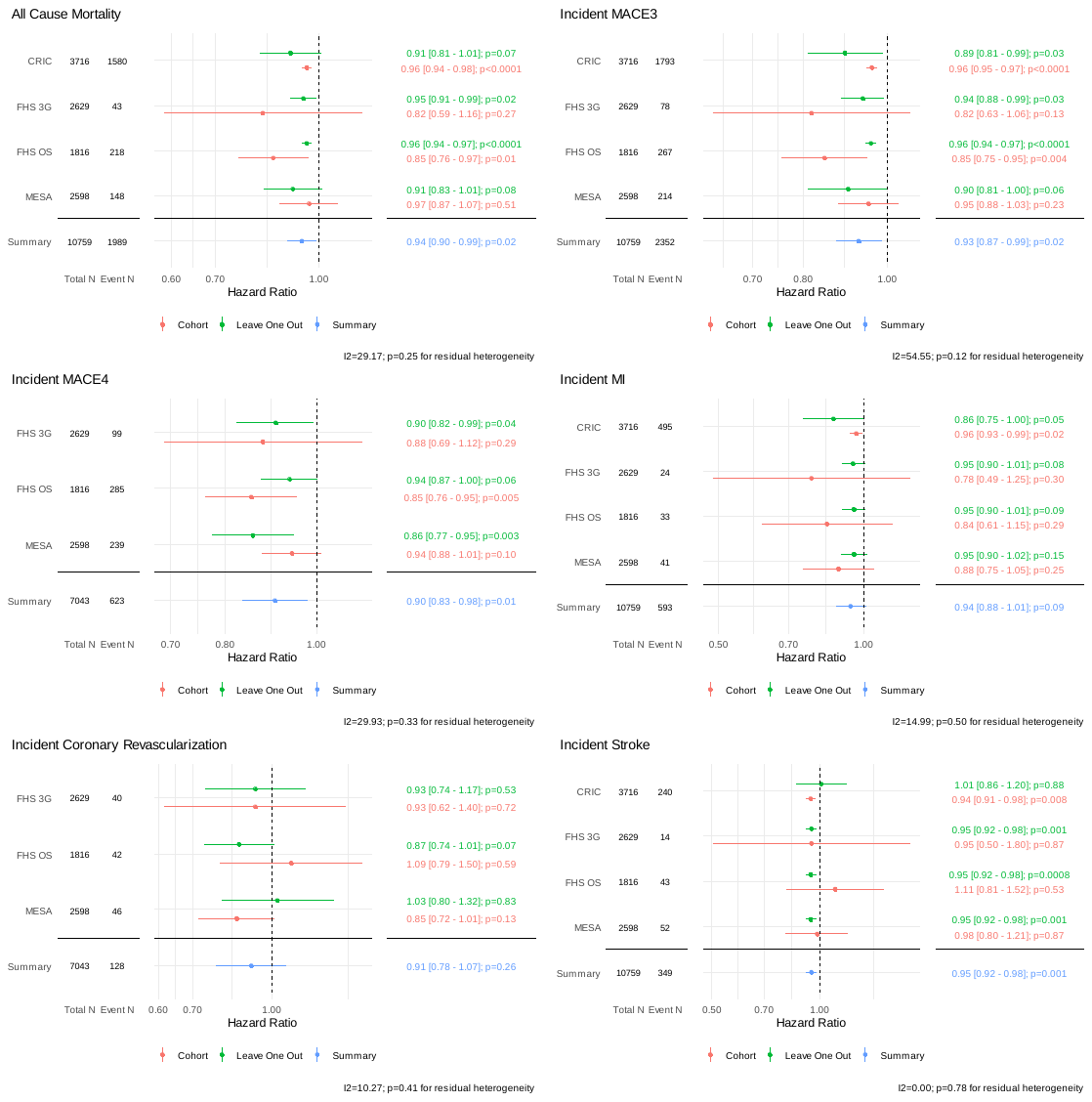

### Mean Corpuscular Volume (Unadjusted)

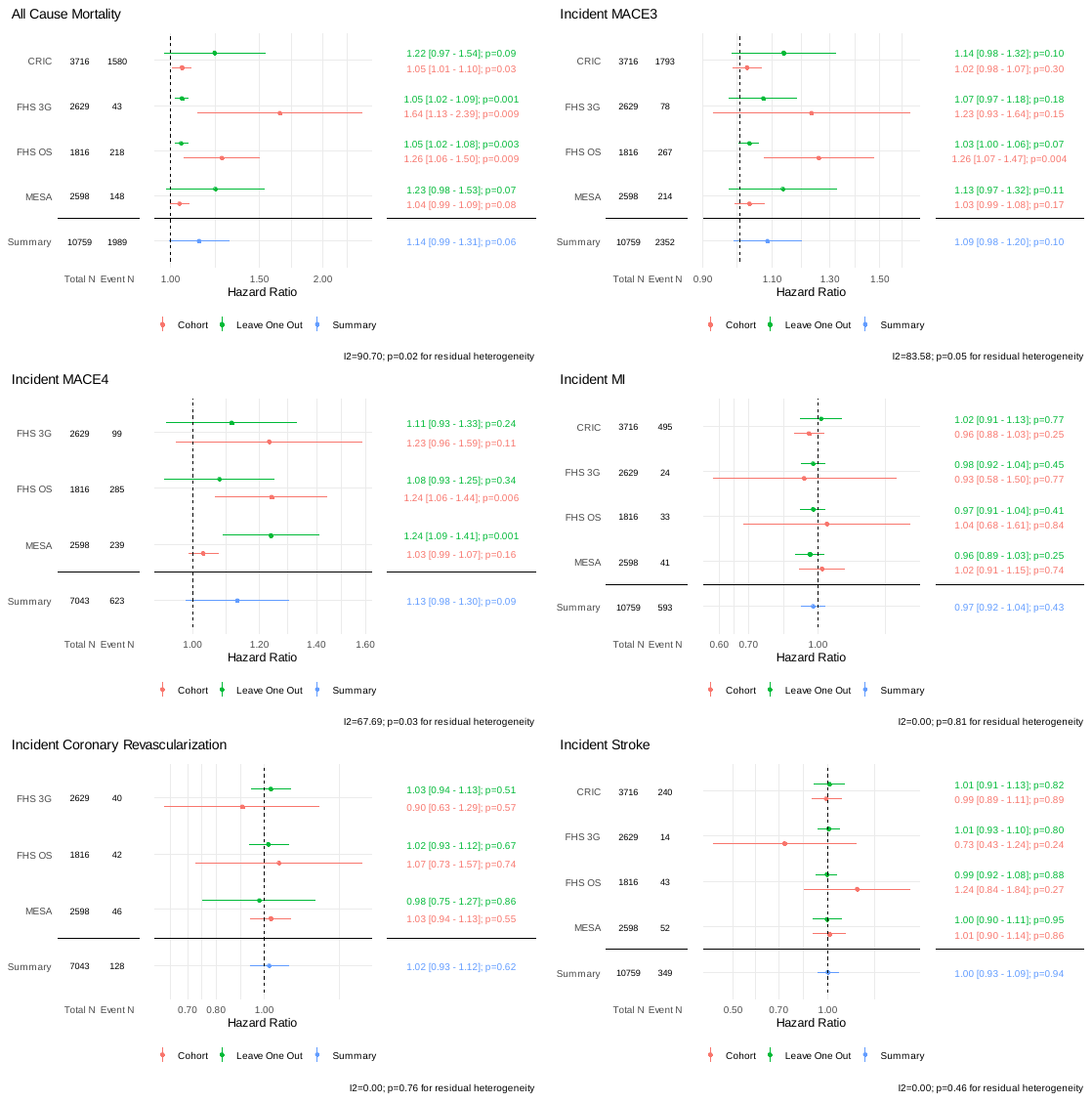

### Mean Corpuscular Volume (Age/Sex/Race Adjusted)

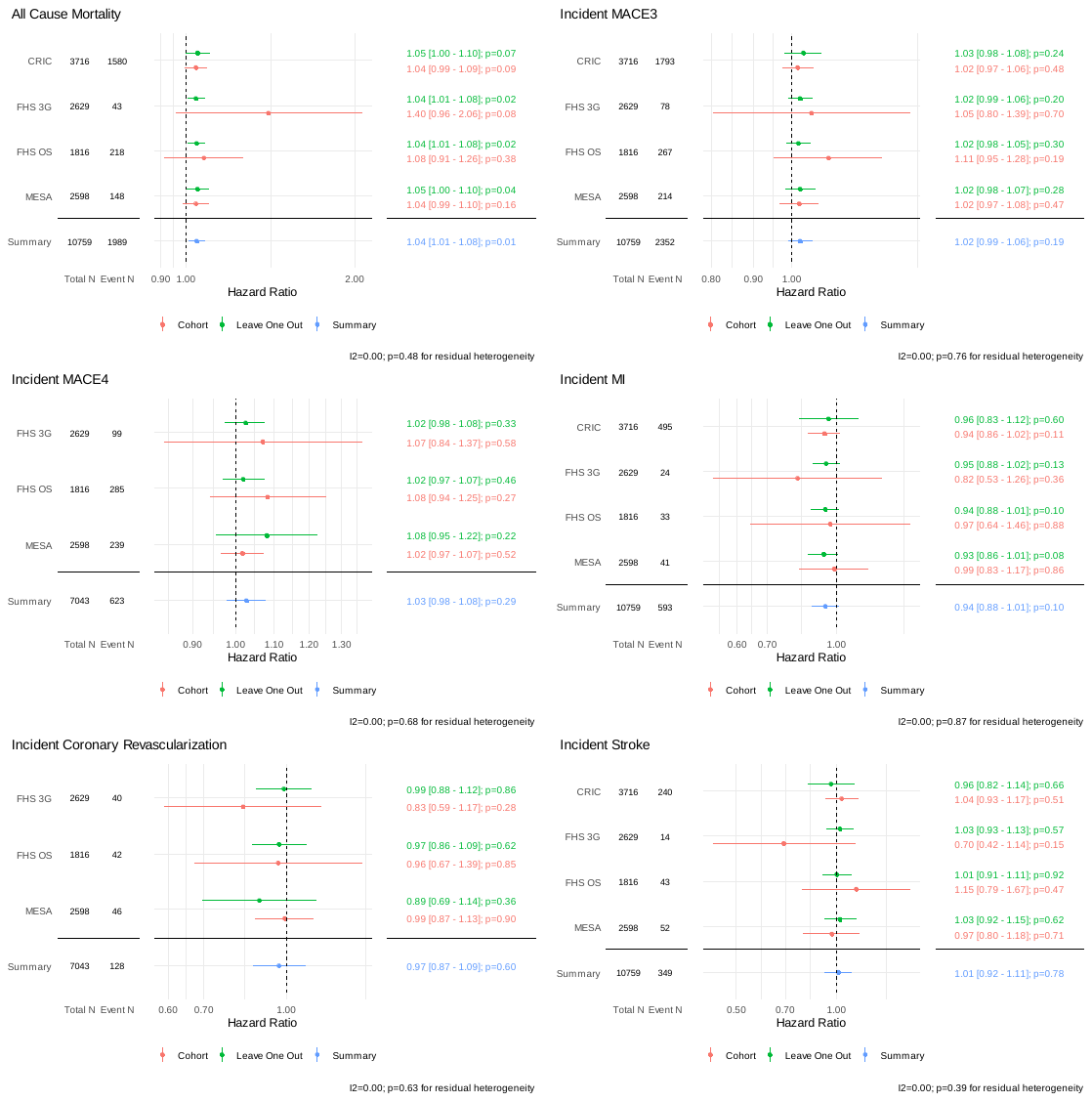

### Mean Corpuscular Volume (Fully Adjusted)

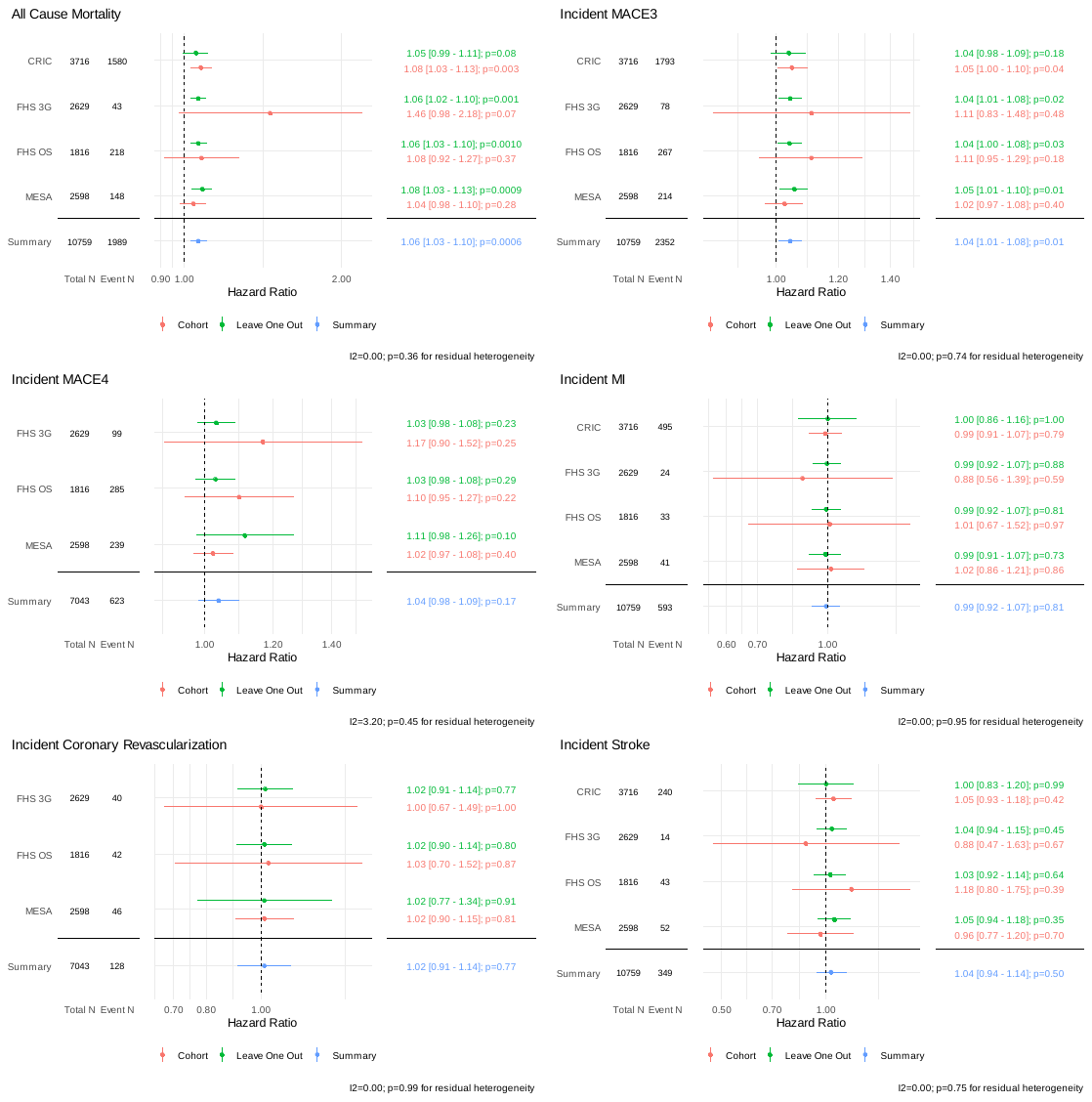

### Mean Corpuscular Volume (Fully Adjusted w/ hsCRP)

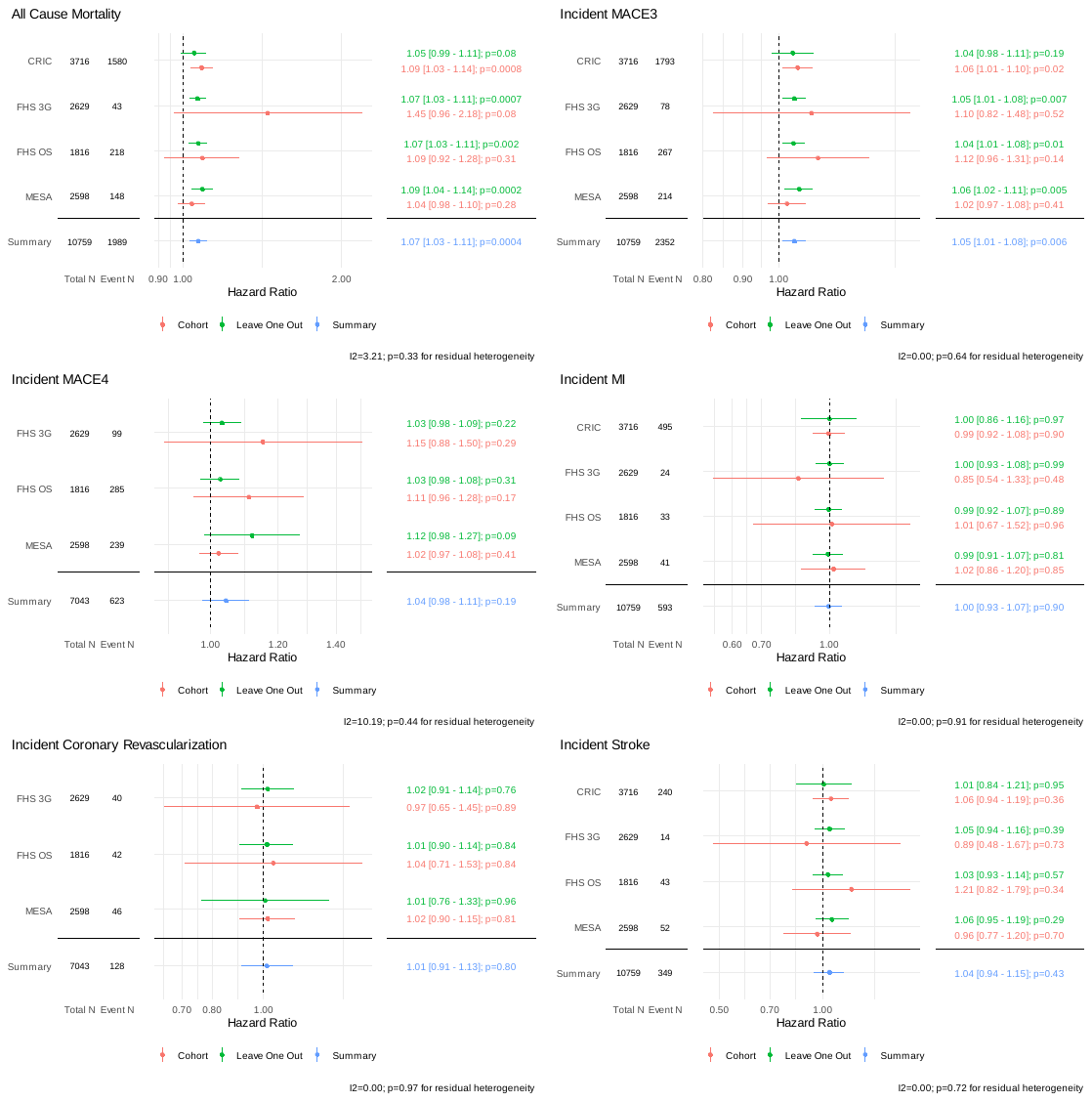

### Monocyte Count (Unadjusted)

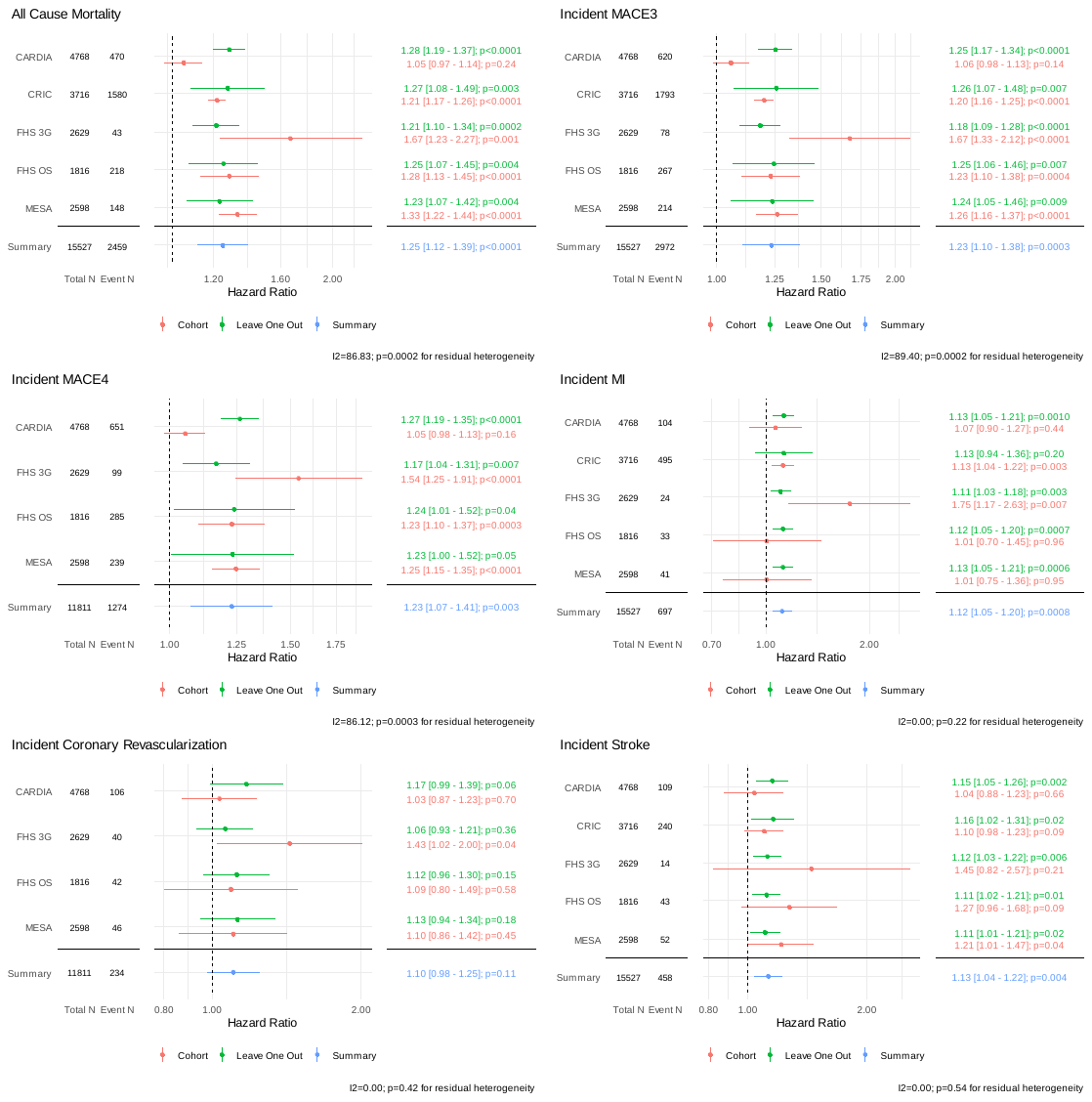

### Monocyte Count (Age/Sex/Race Adjusted)

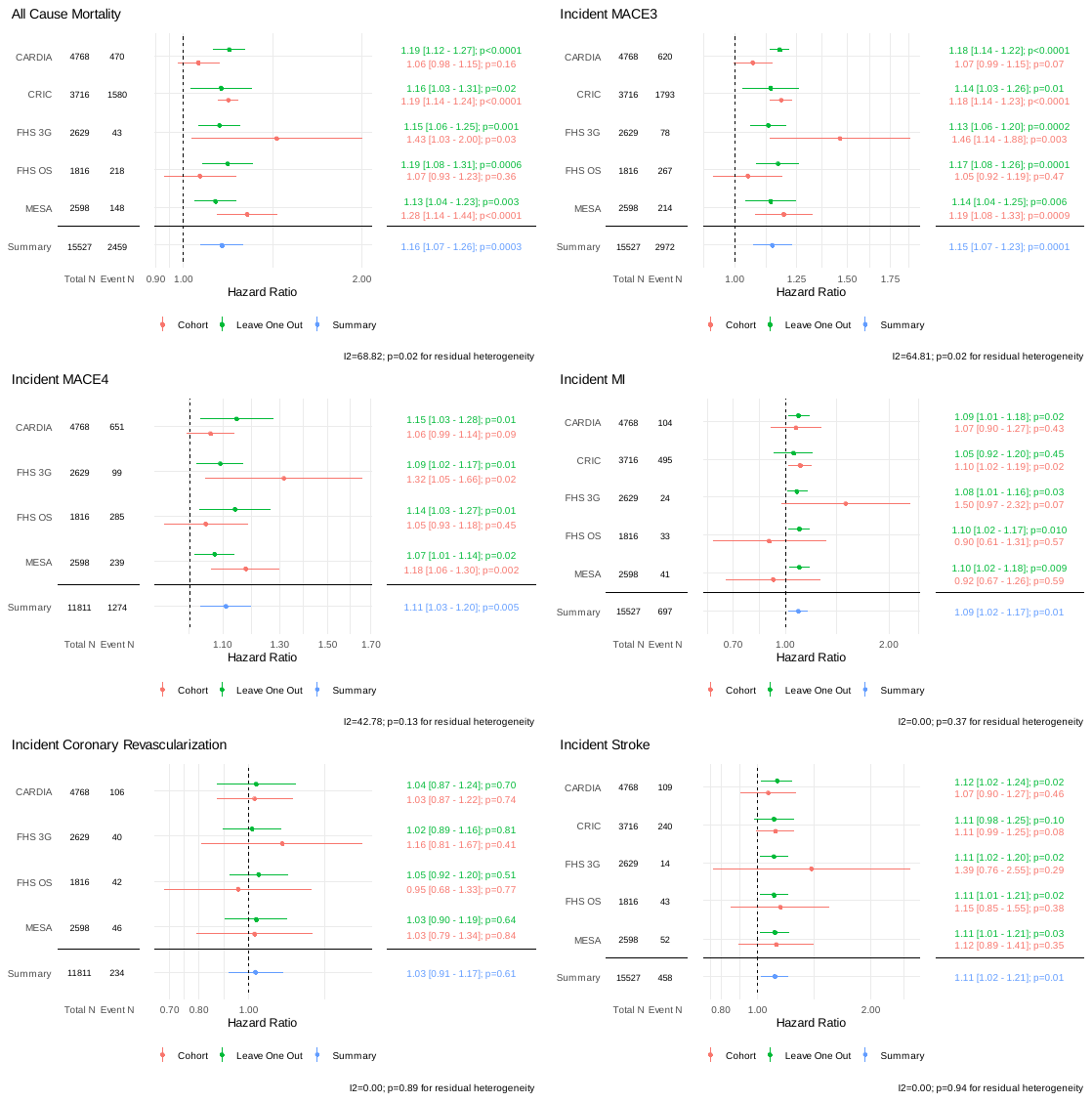

### Monocyte Count (Fully Adjusted)

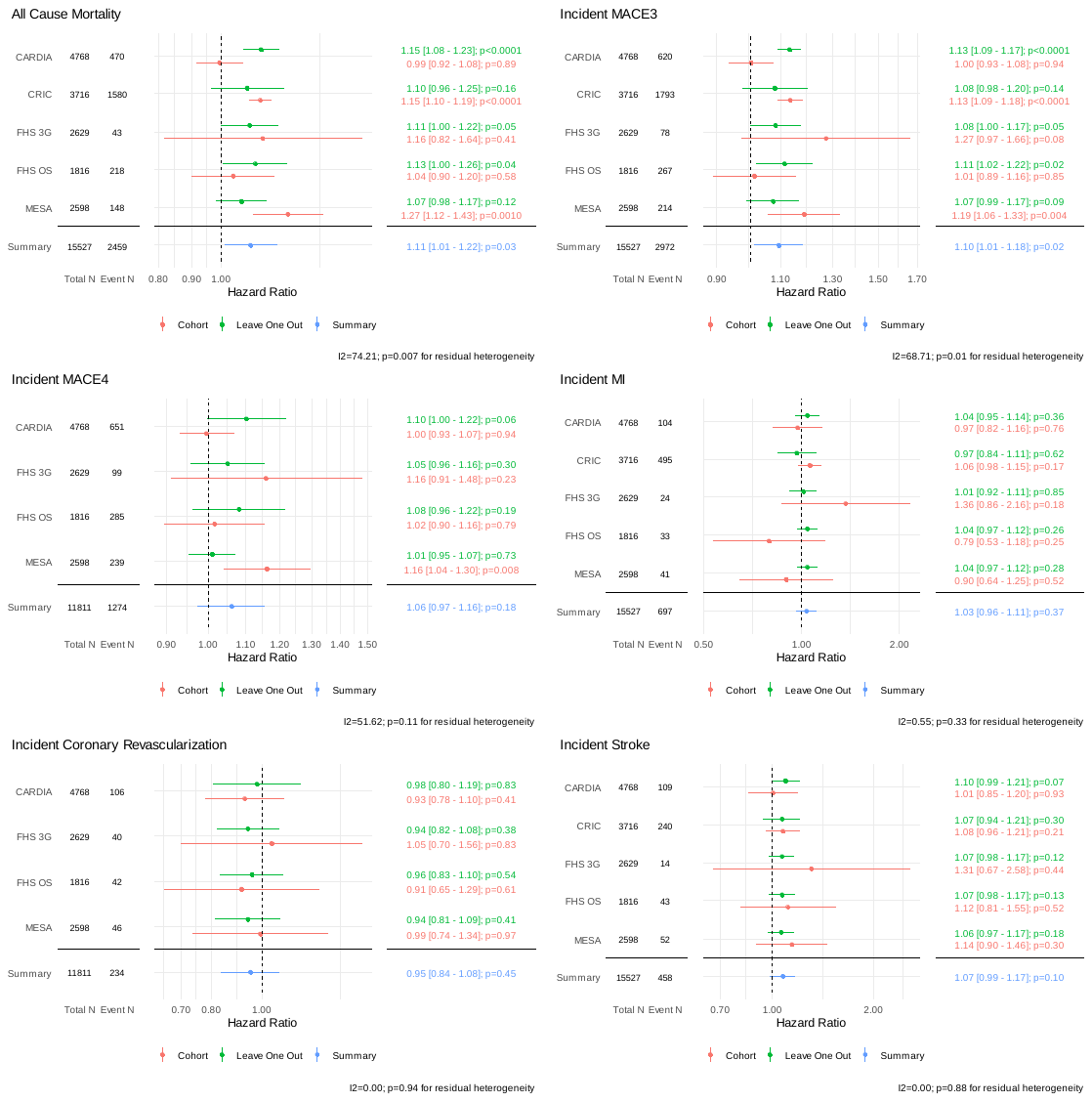

### Monocyte Count (Fully Adjusted w/ hsCRP)

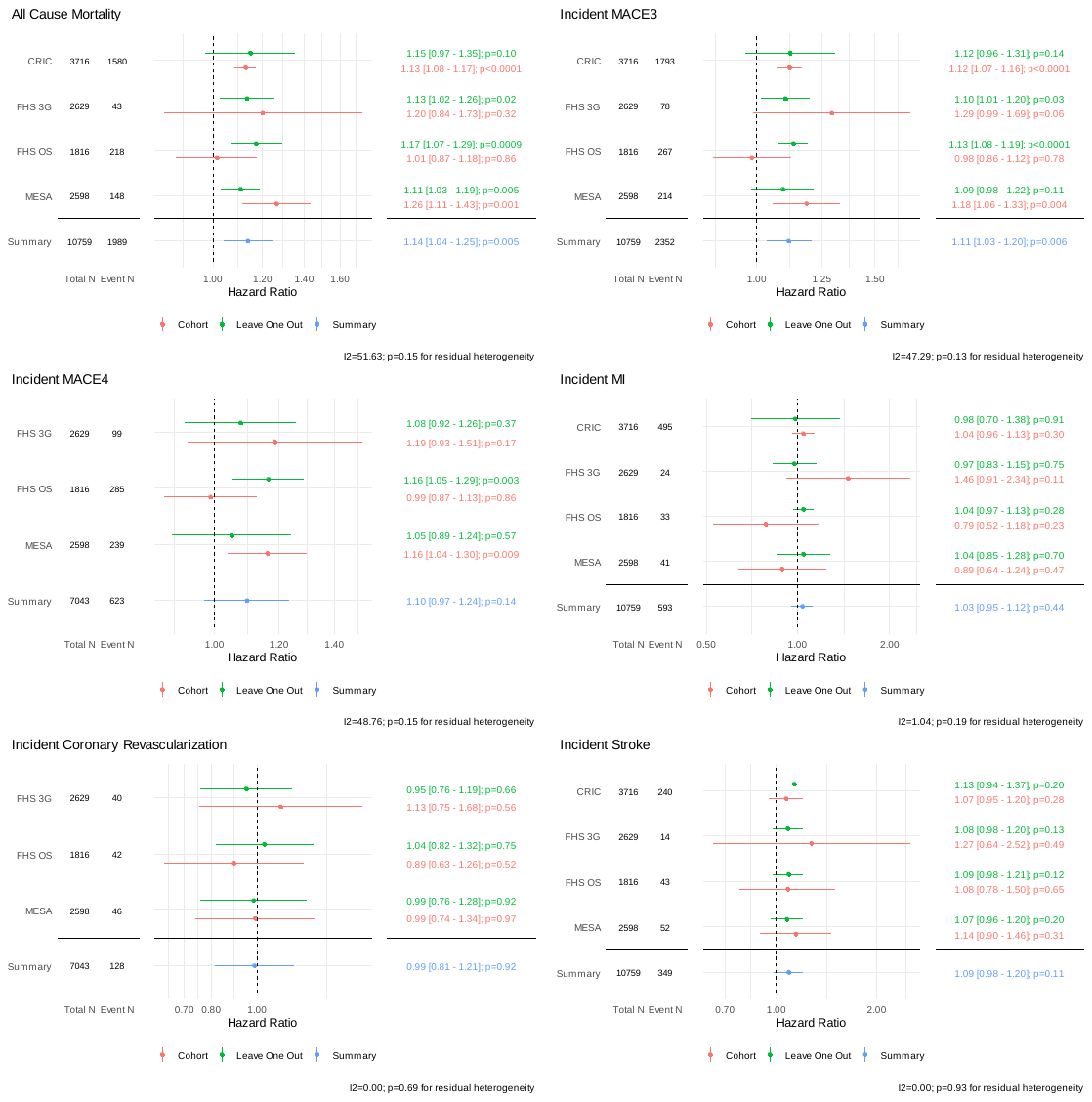

### Monocytes (%) (Unadjusted)

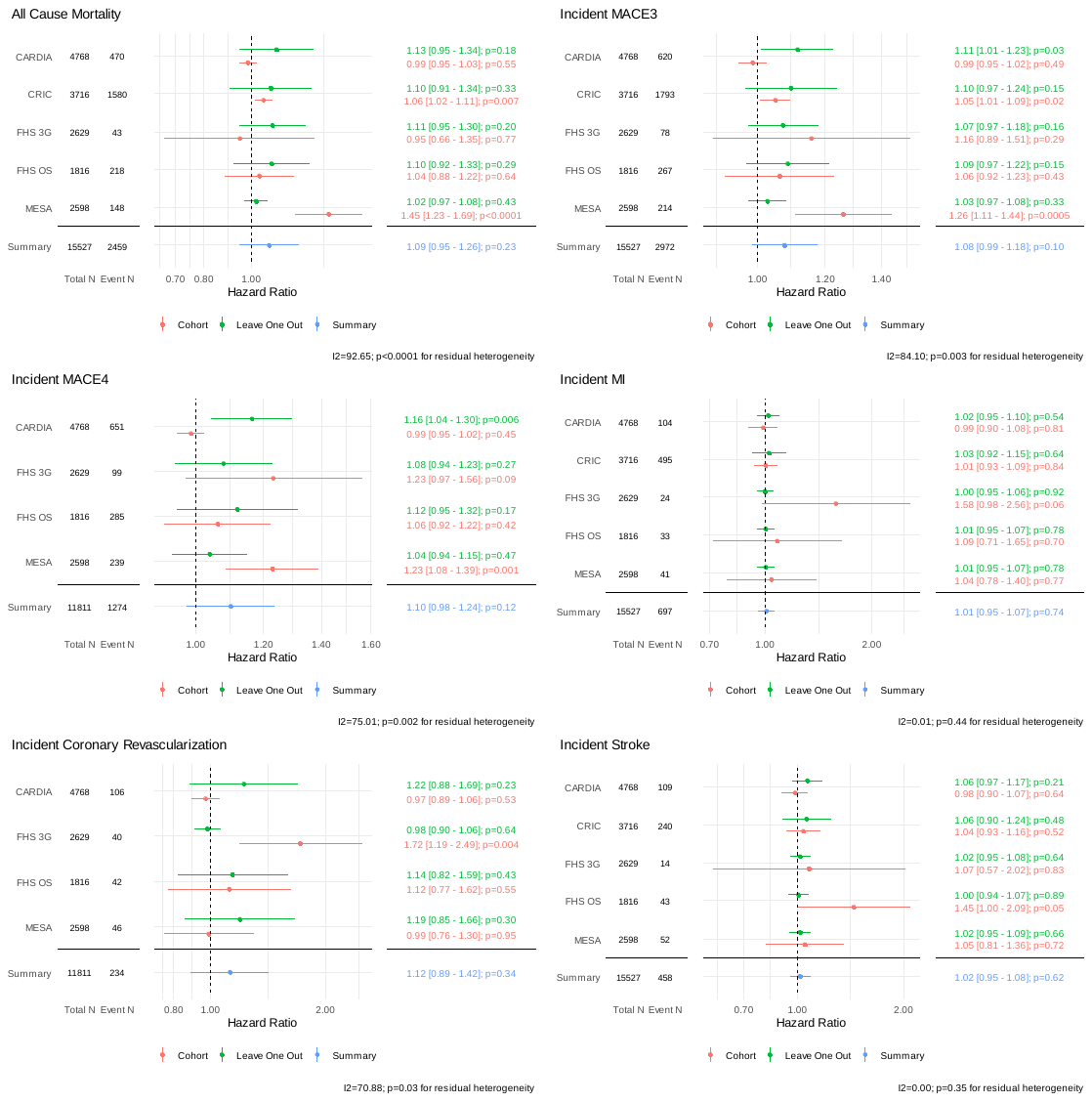

### Monocytes (%) (Age/Sex/Race Adjusted)

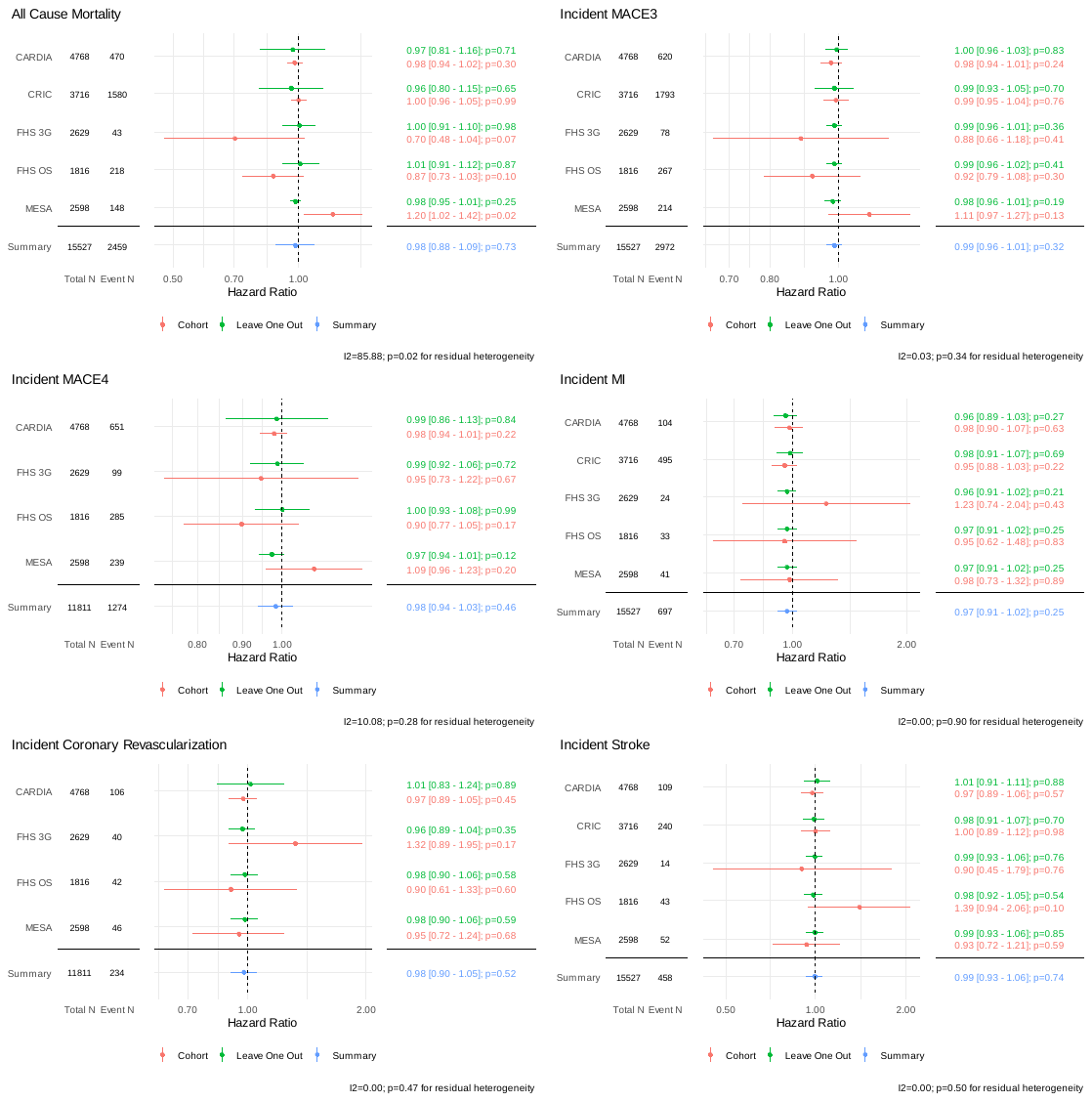

### Monocytes (%) (Fully Adjusted)

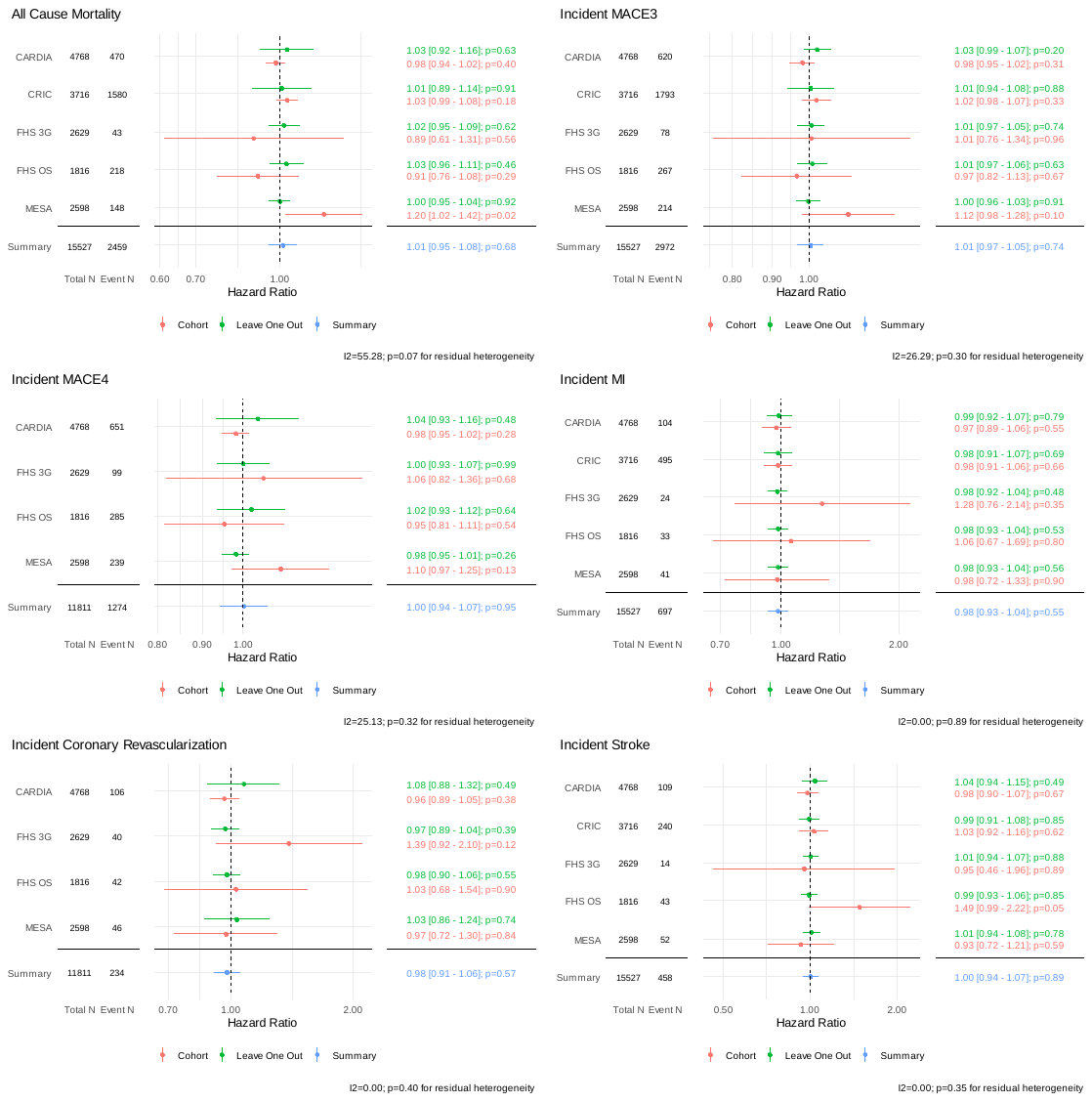

### Monocytes (%) (Fully Adjusted w/ hsCRP)

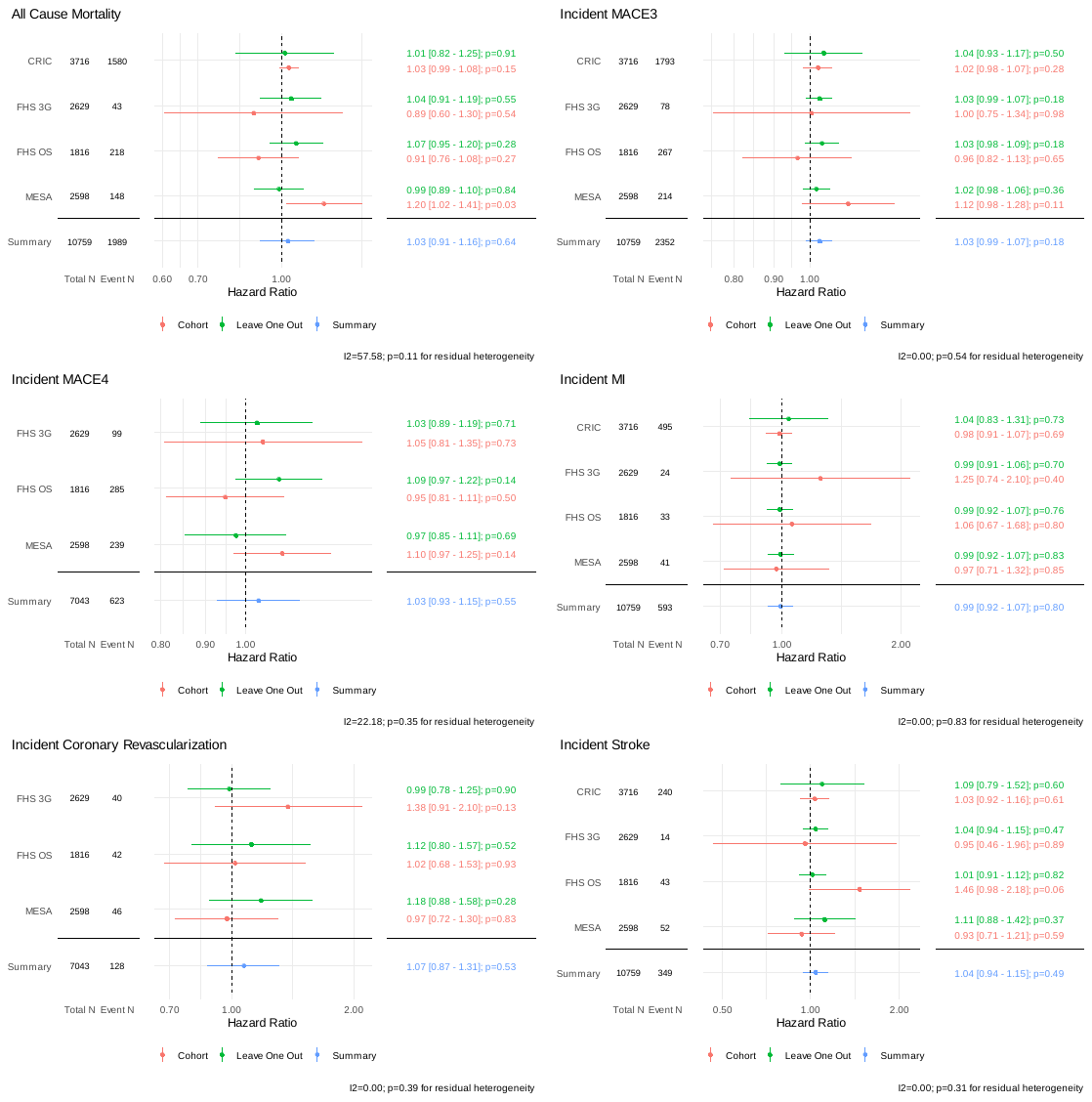

### Neutrophil Count (Unadjusted)

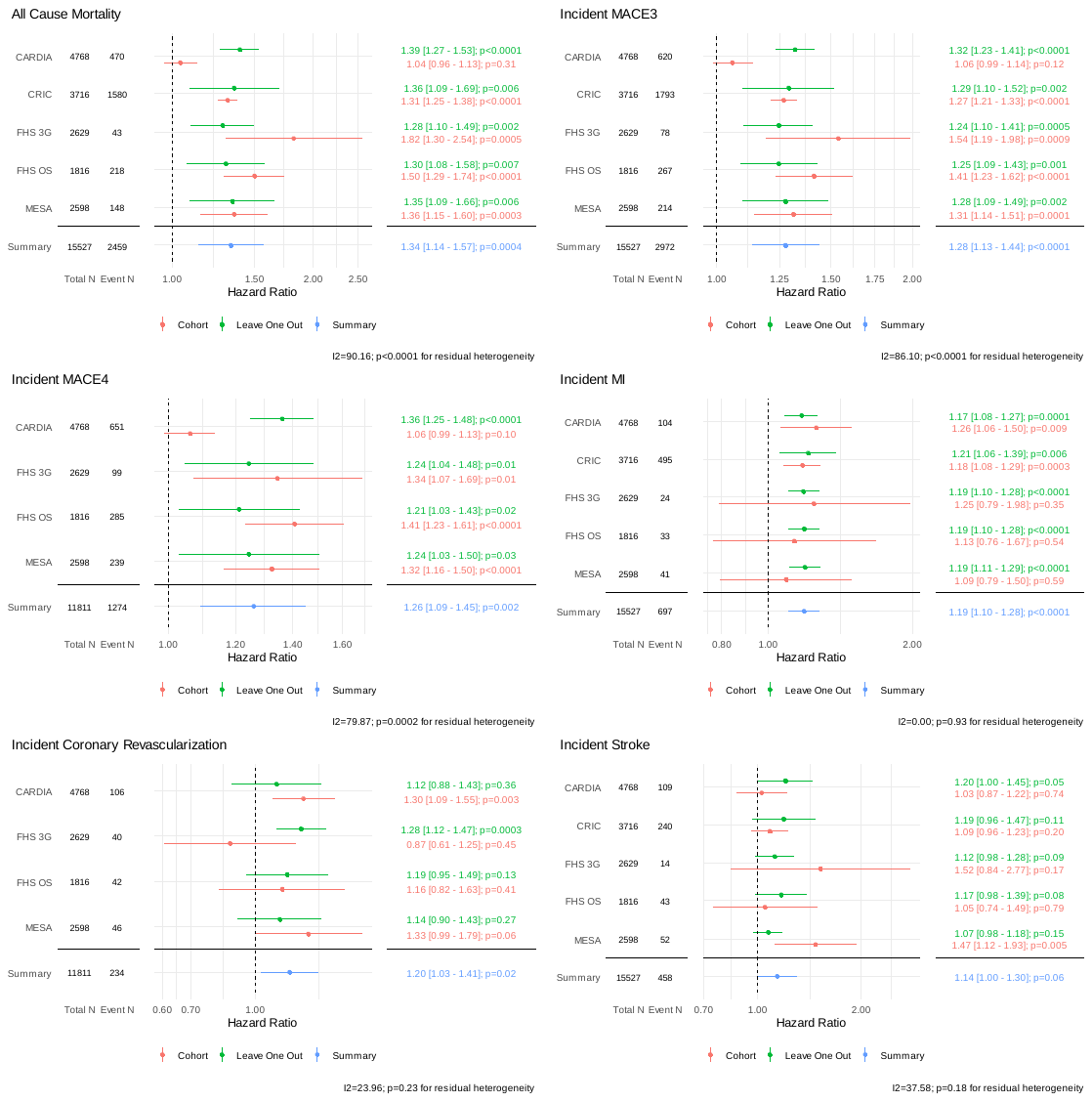

### Neutrophil Count (Age/Sex/Race Adjusted)

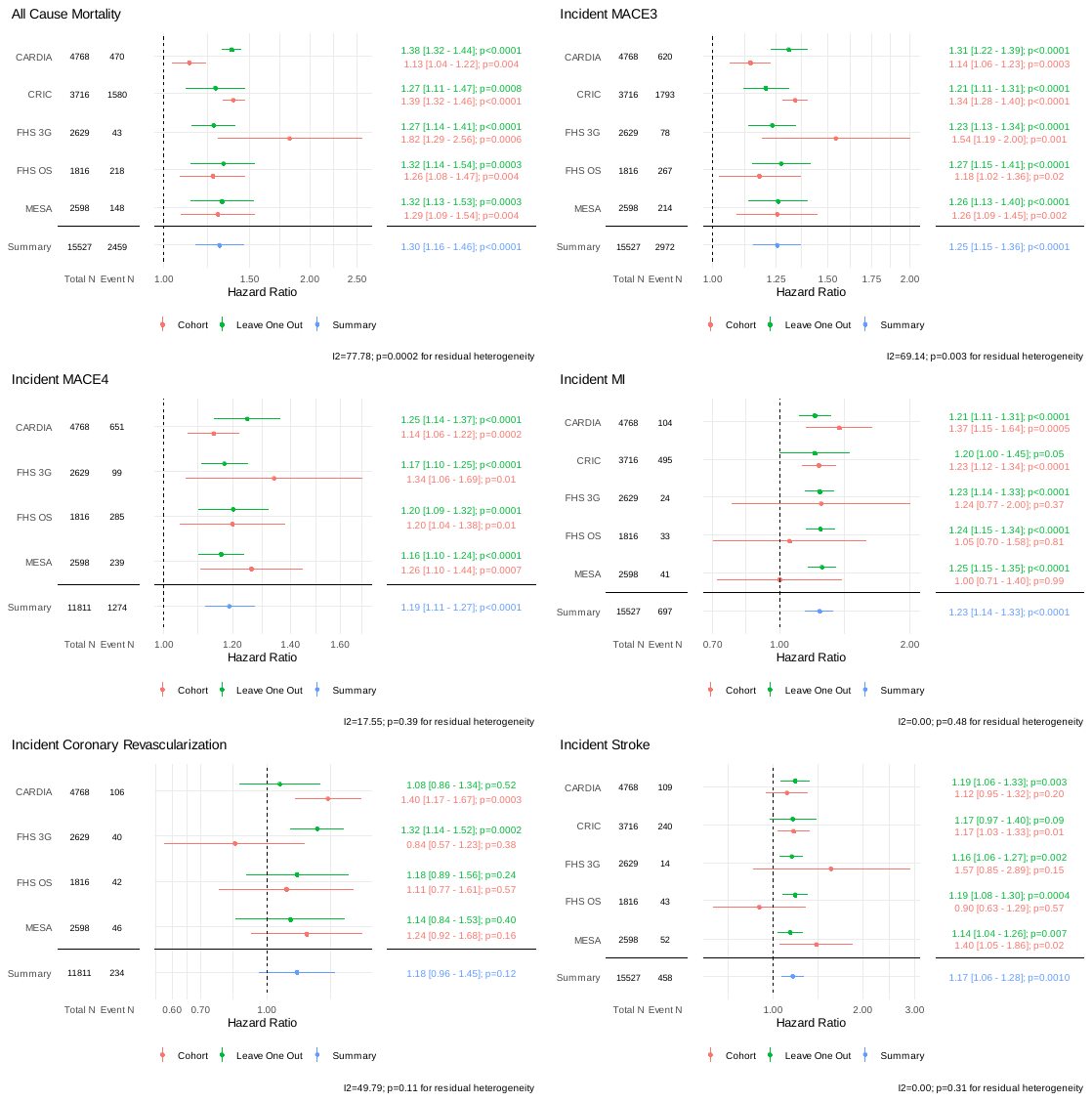

### Neutrophil Count (Fully Adjusted)

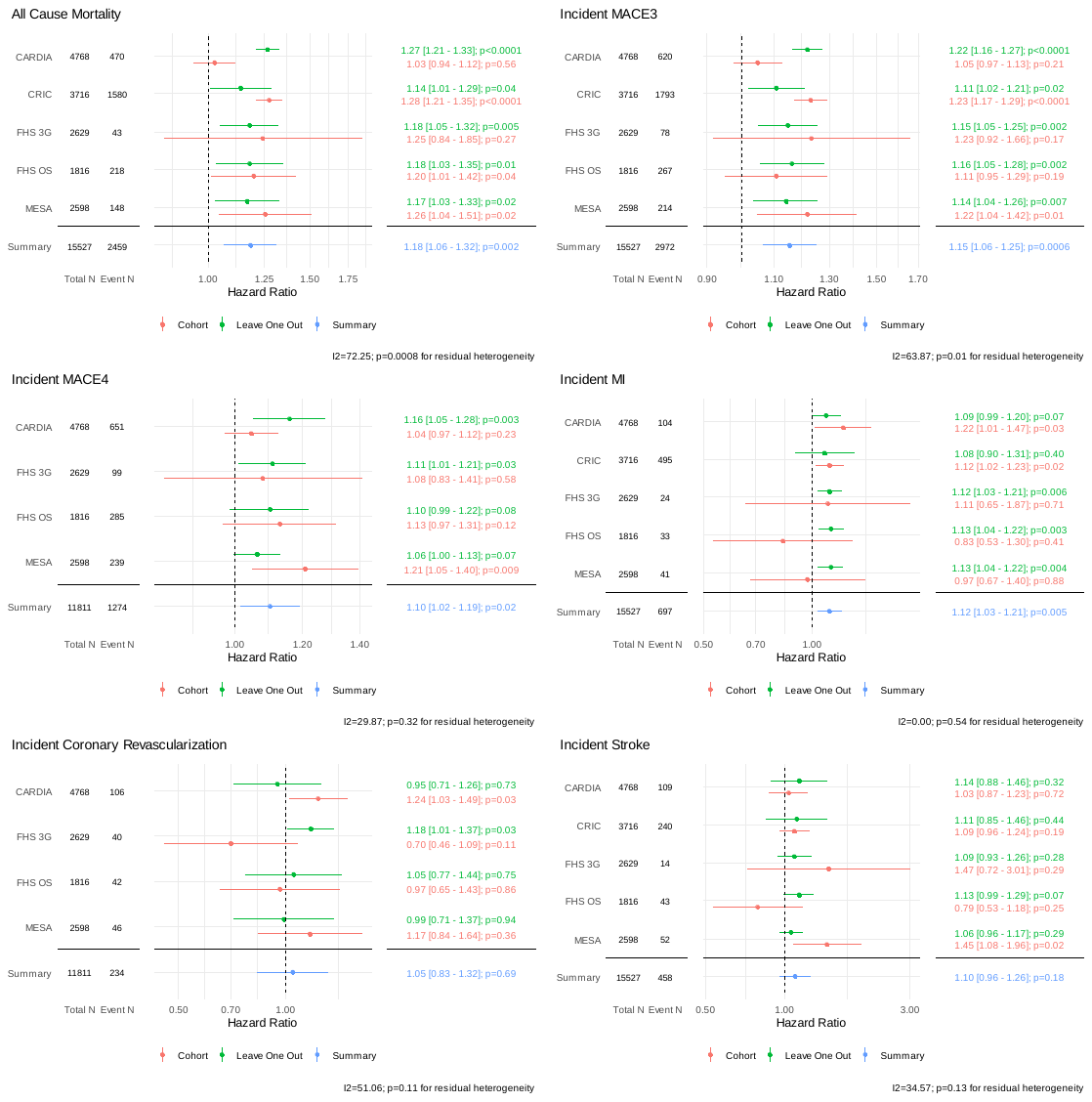

### Neutrophil Count (Fully Adjusted w/ hsCRP)

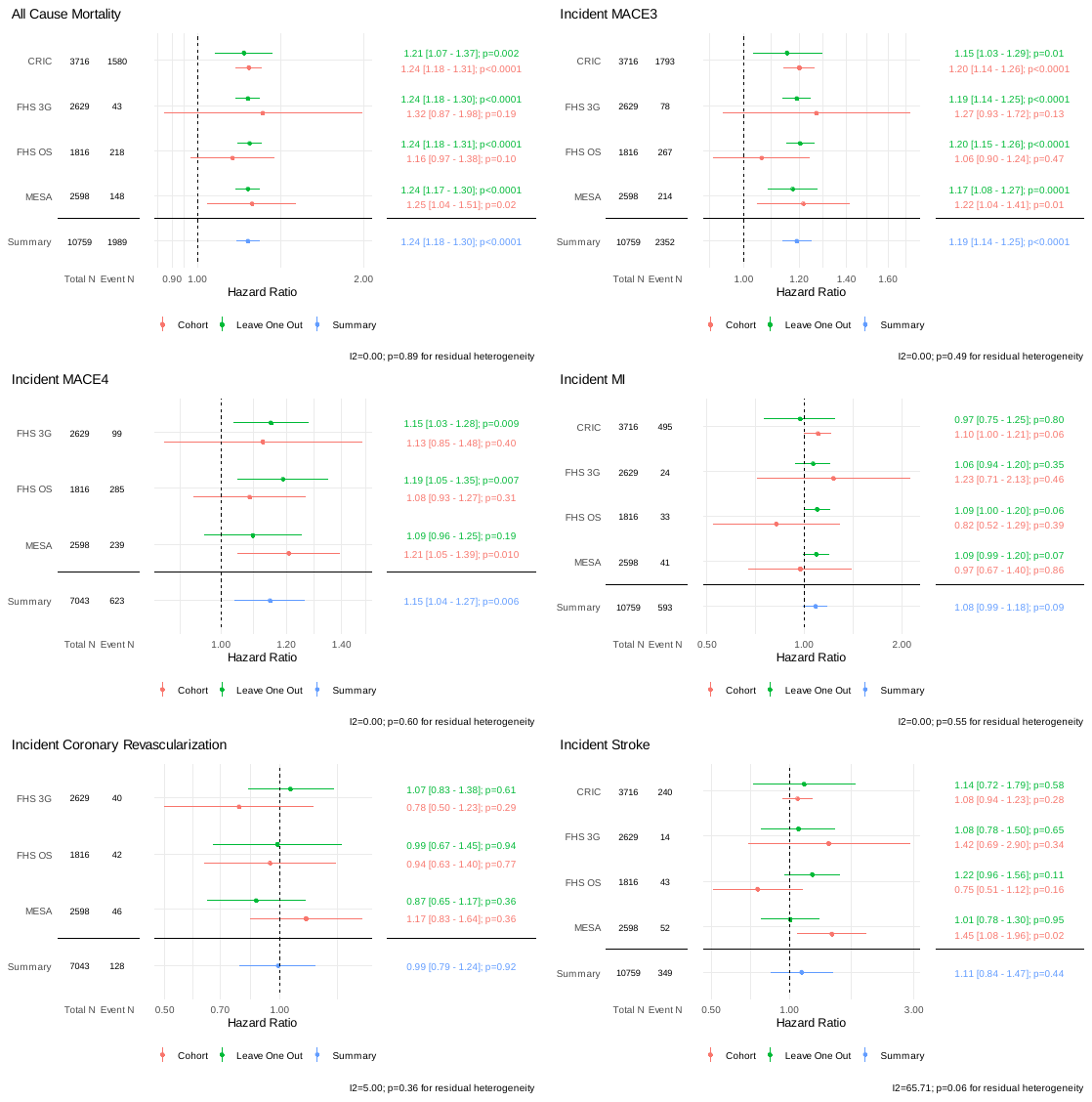

### Neutrophil:Lymphocyte Ratio (Unadjusted)

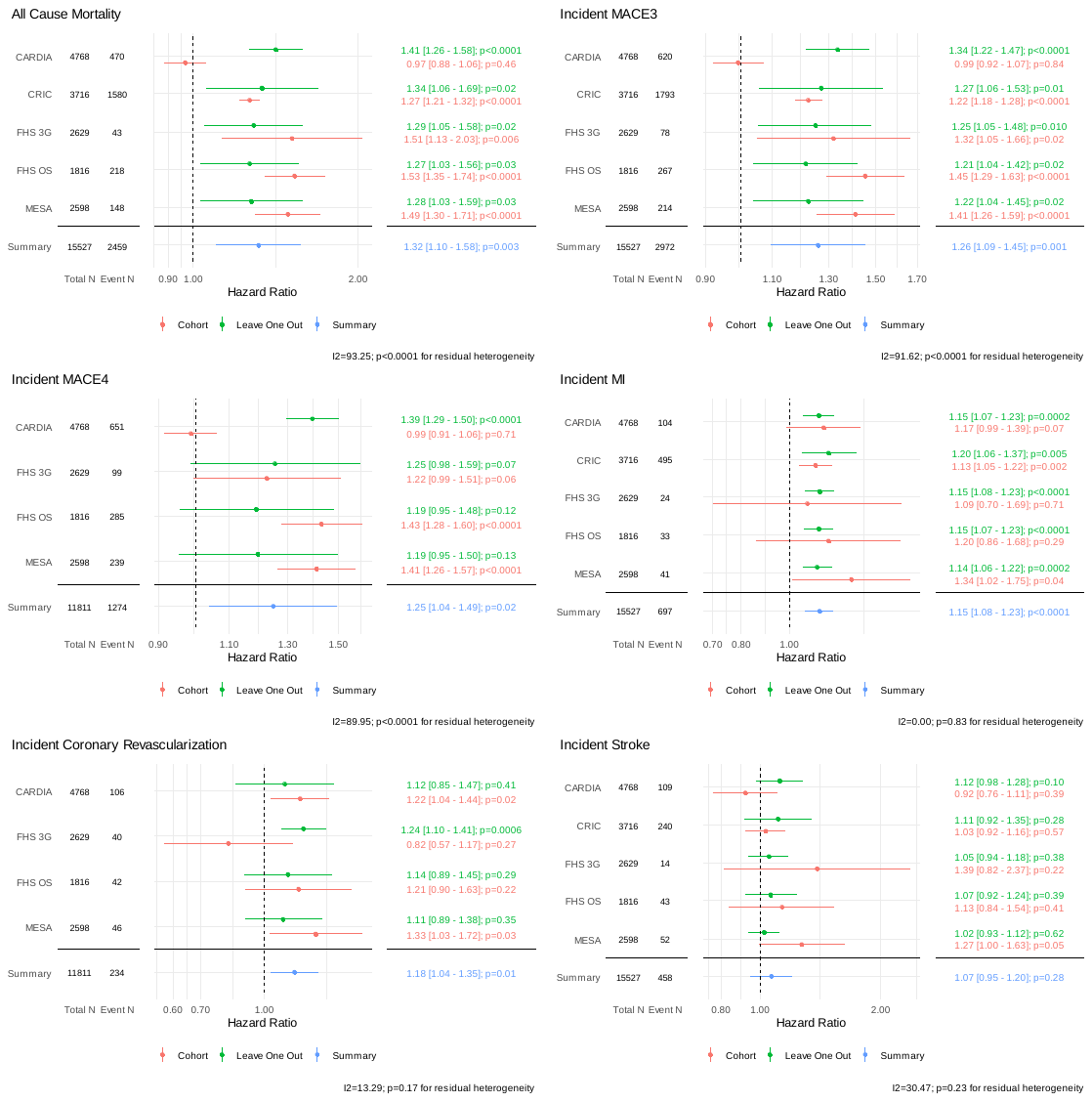

### Neutrophil:Lymphocyte Ratio (Age/Sex/Race Adjusted)

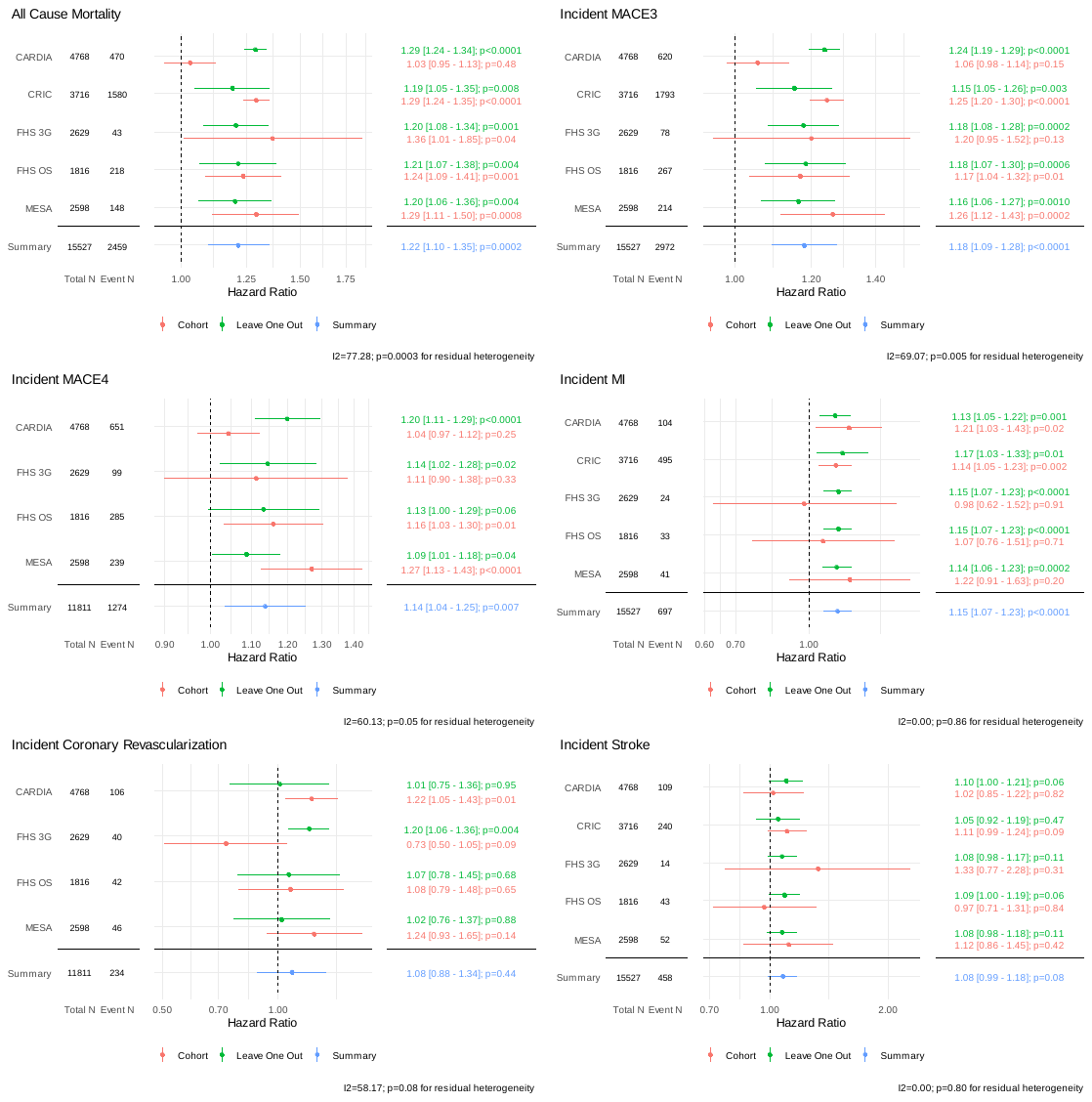

### Neutrophil:Lymphocyte Ratio (Fully Adjusted)

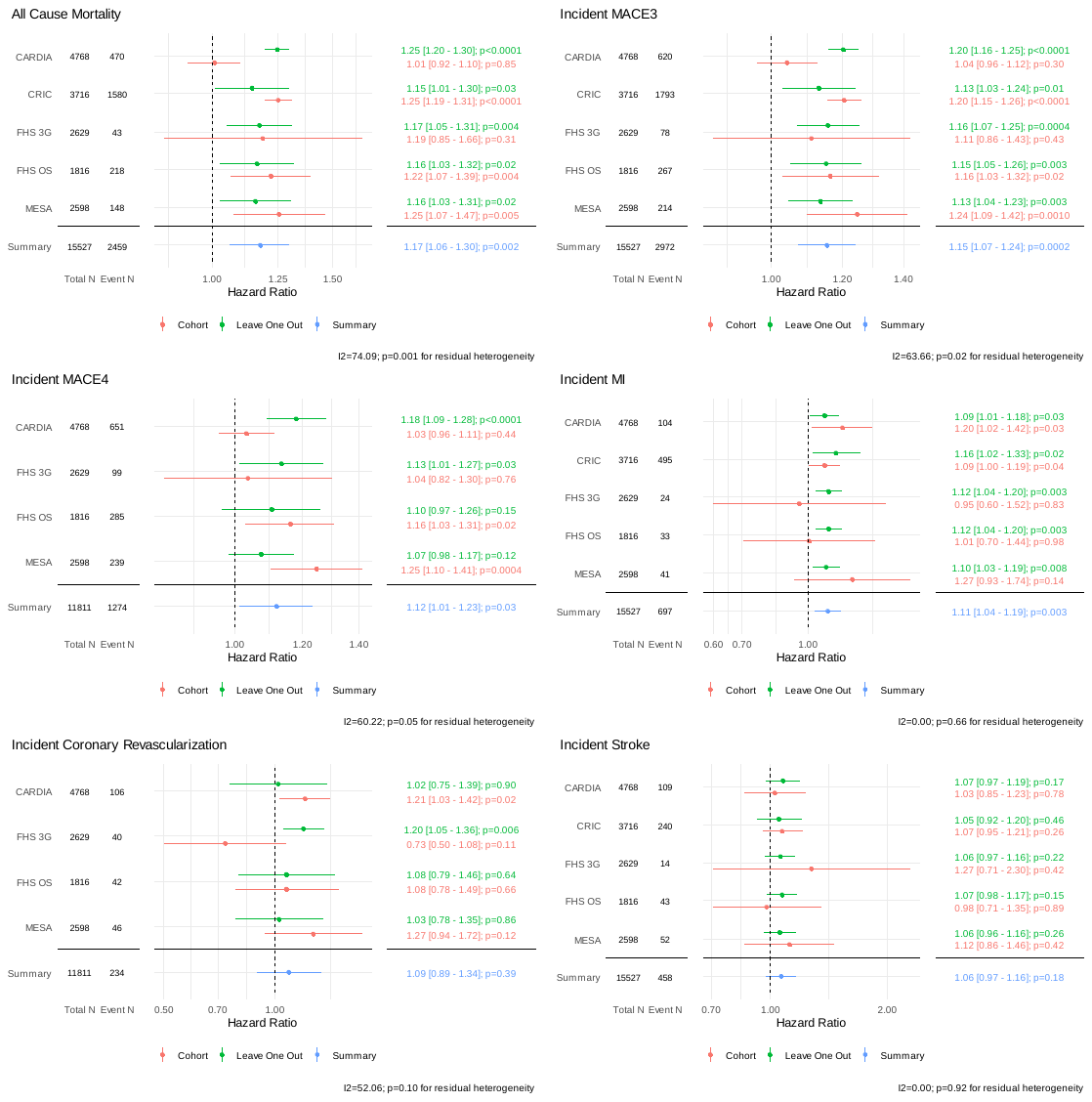

### Neutrophil:Lymphocyte Ratio (Fully Adjusted w/ hsCRP)

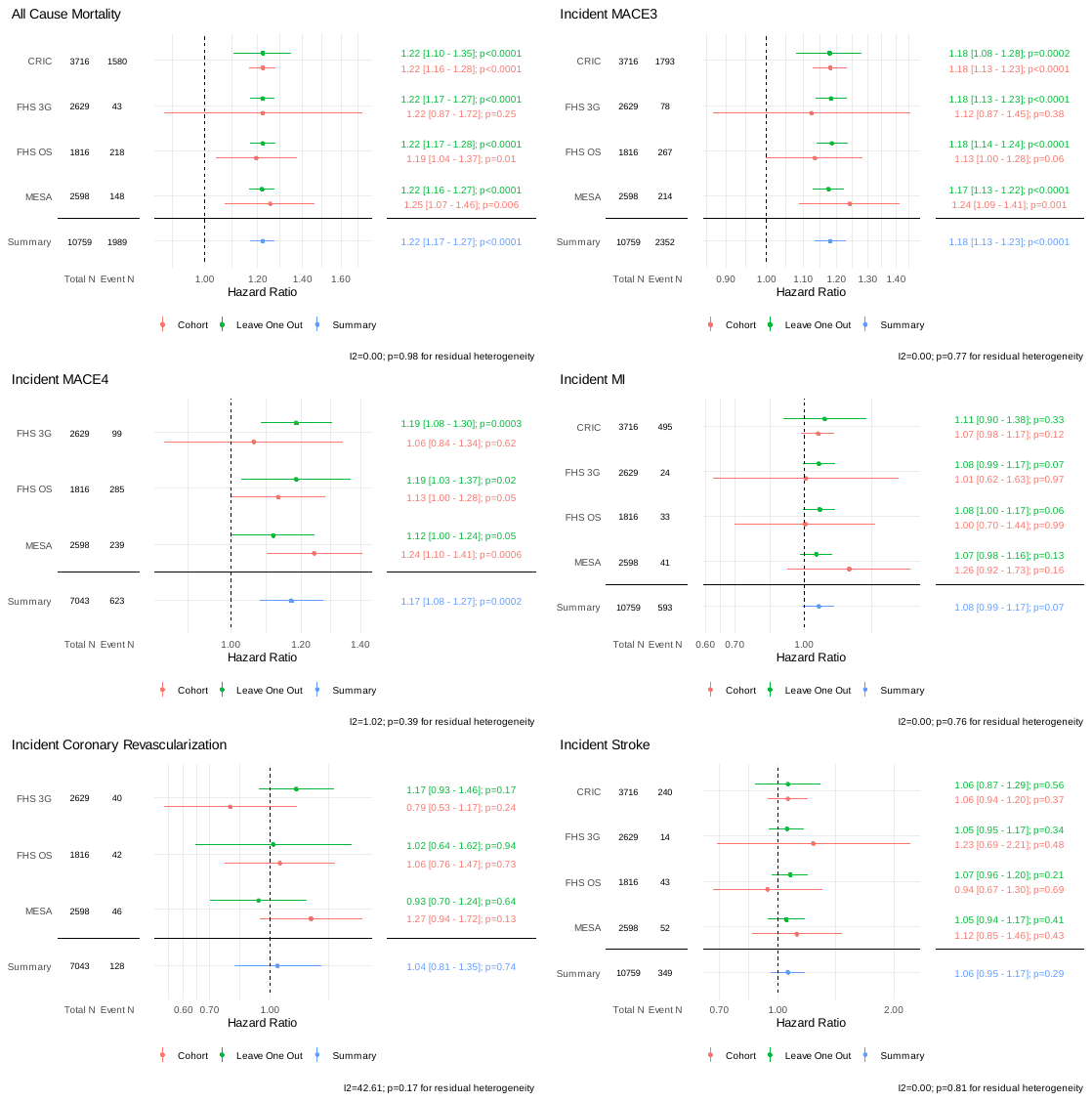

### Neutrophils (%) (Unadjusted)

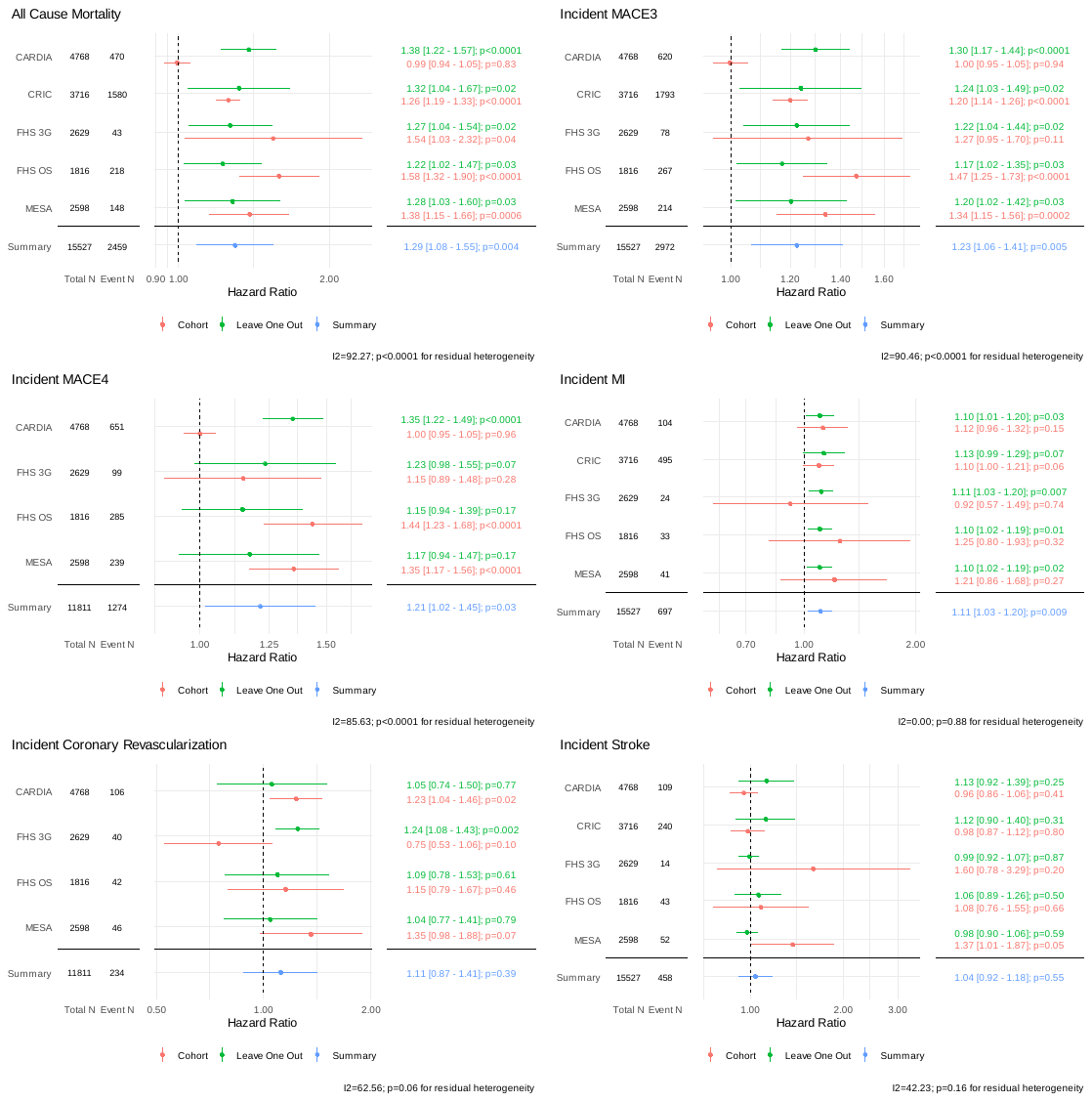

### Neutrophils (%) (Age/Sex/Race Adjusted)

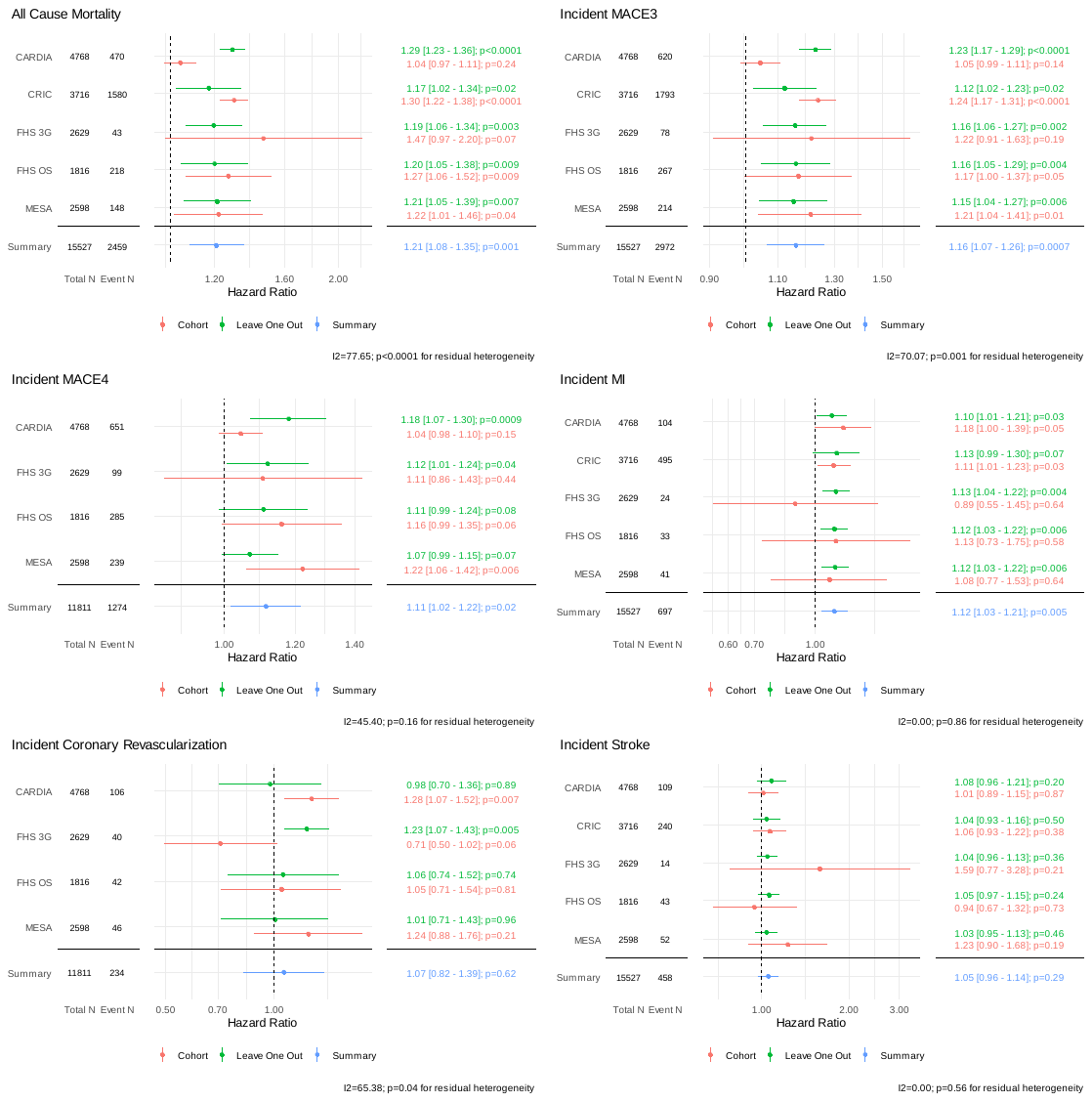

### Neutrophils (%) (Fully Adjusted)

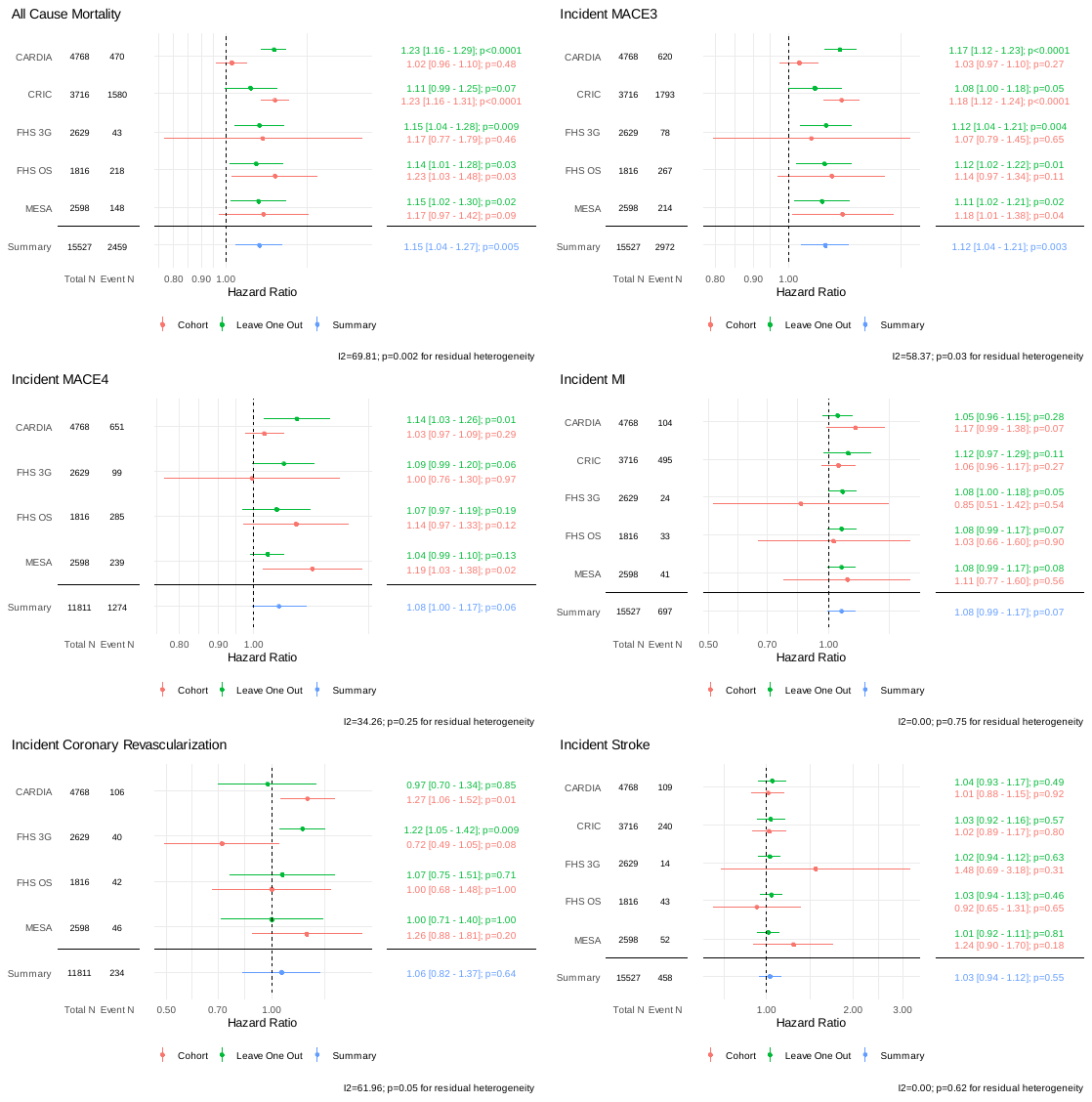

### Neutrophils (%) (Fully Adjusted w/ hsCRP)

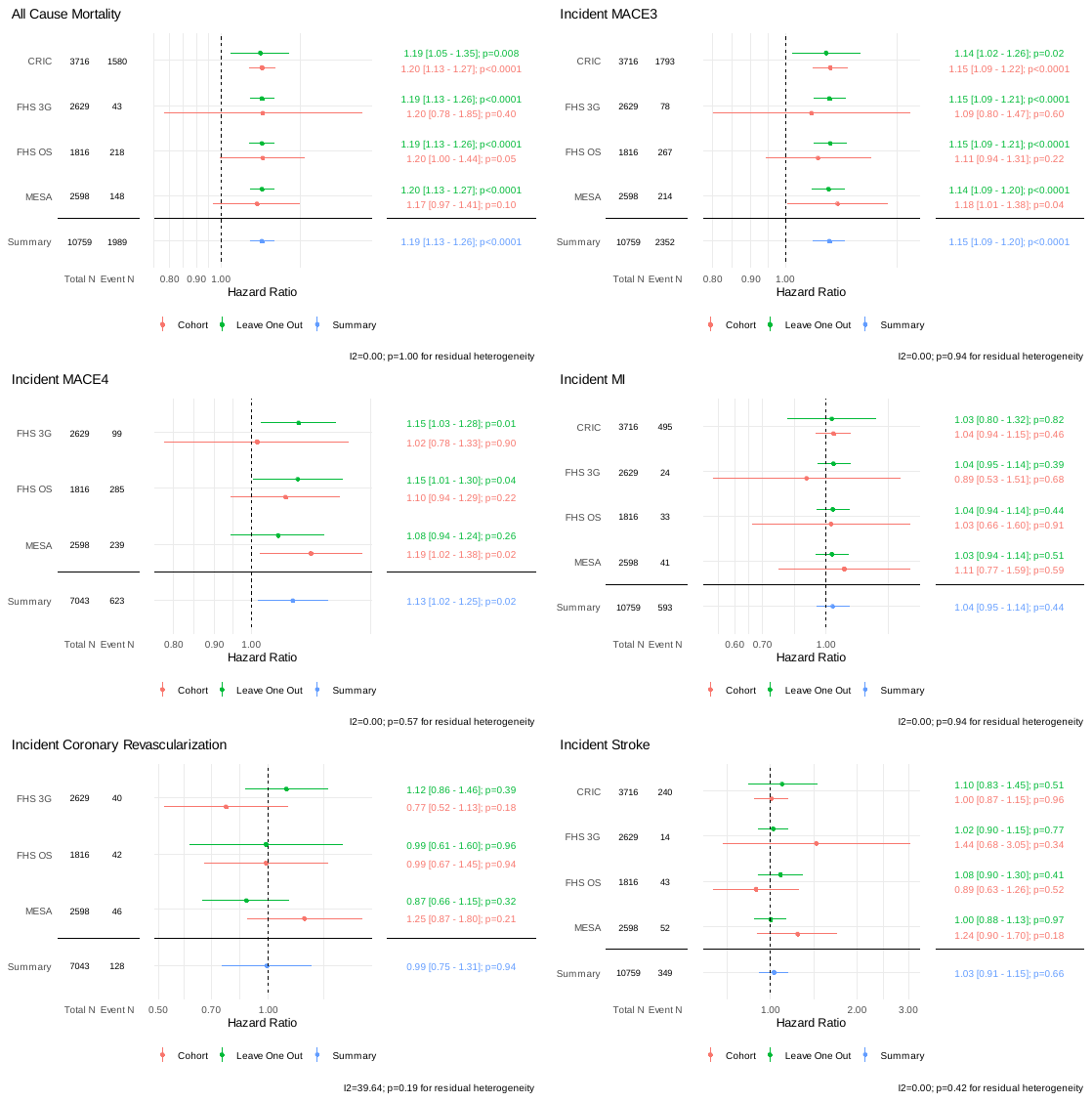

### Platelet Count (Unadjusted)

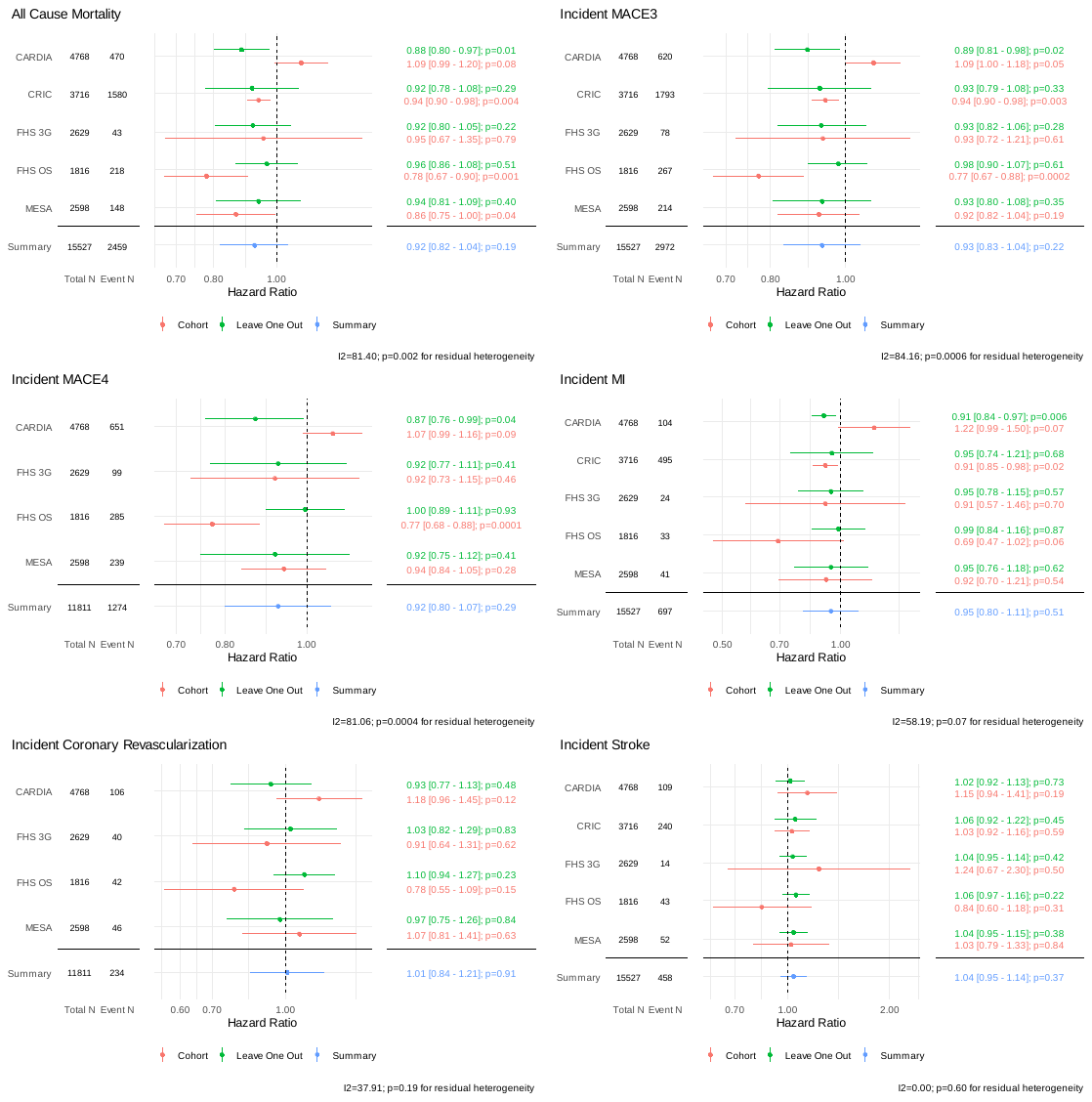

### Platelet Count (Age/Sex/Race Adjusted)

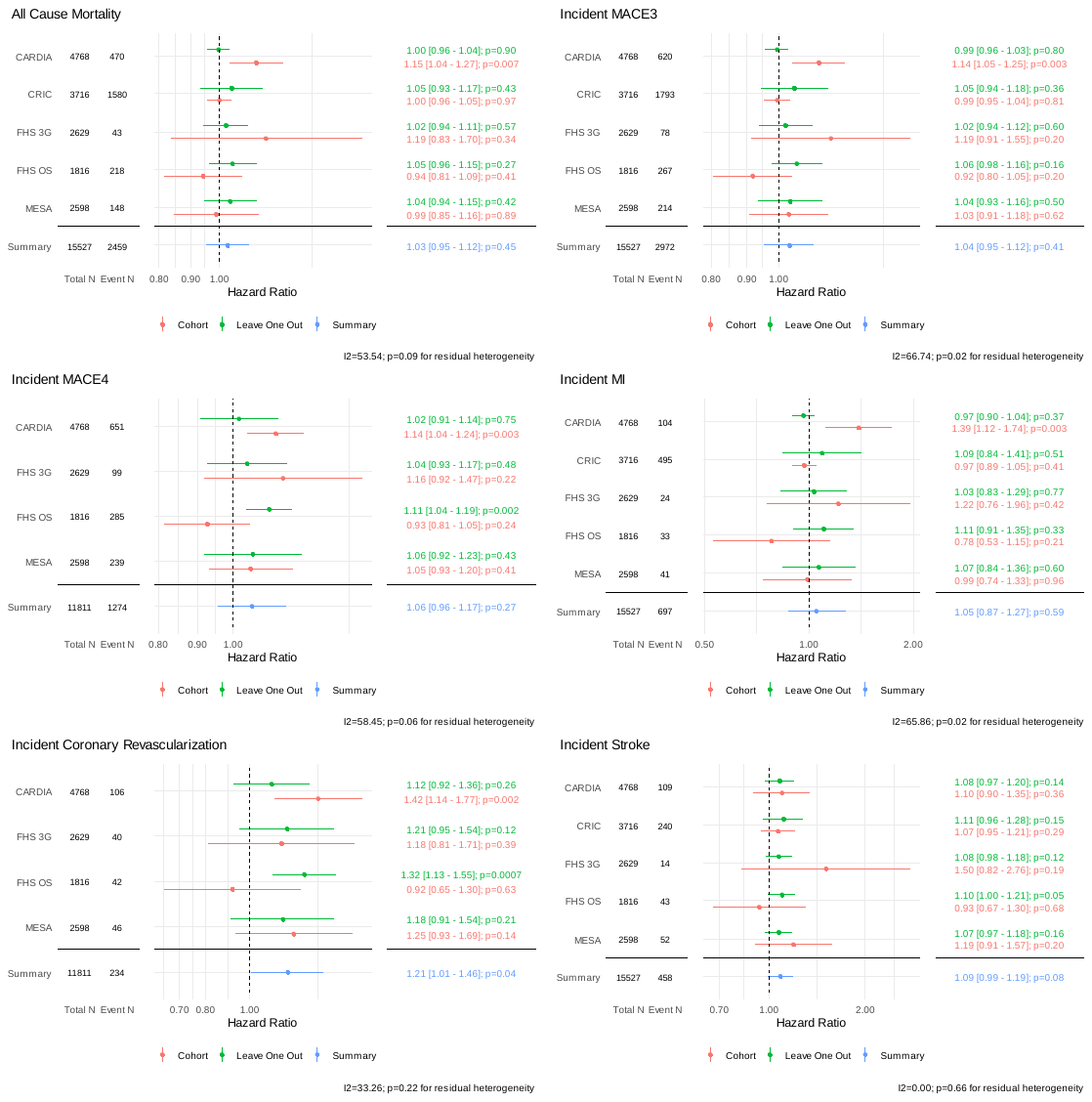

### Platelet Count (Fully Adjusted)

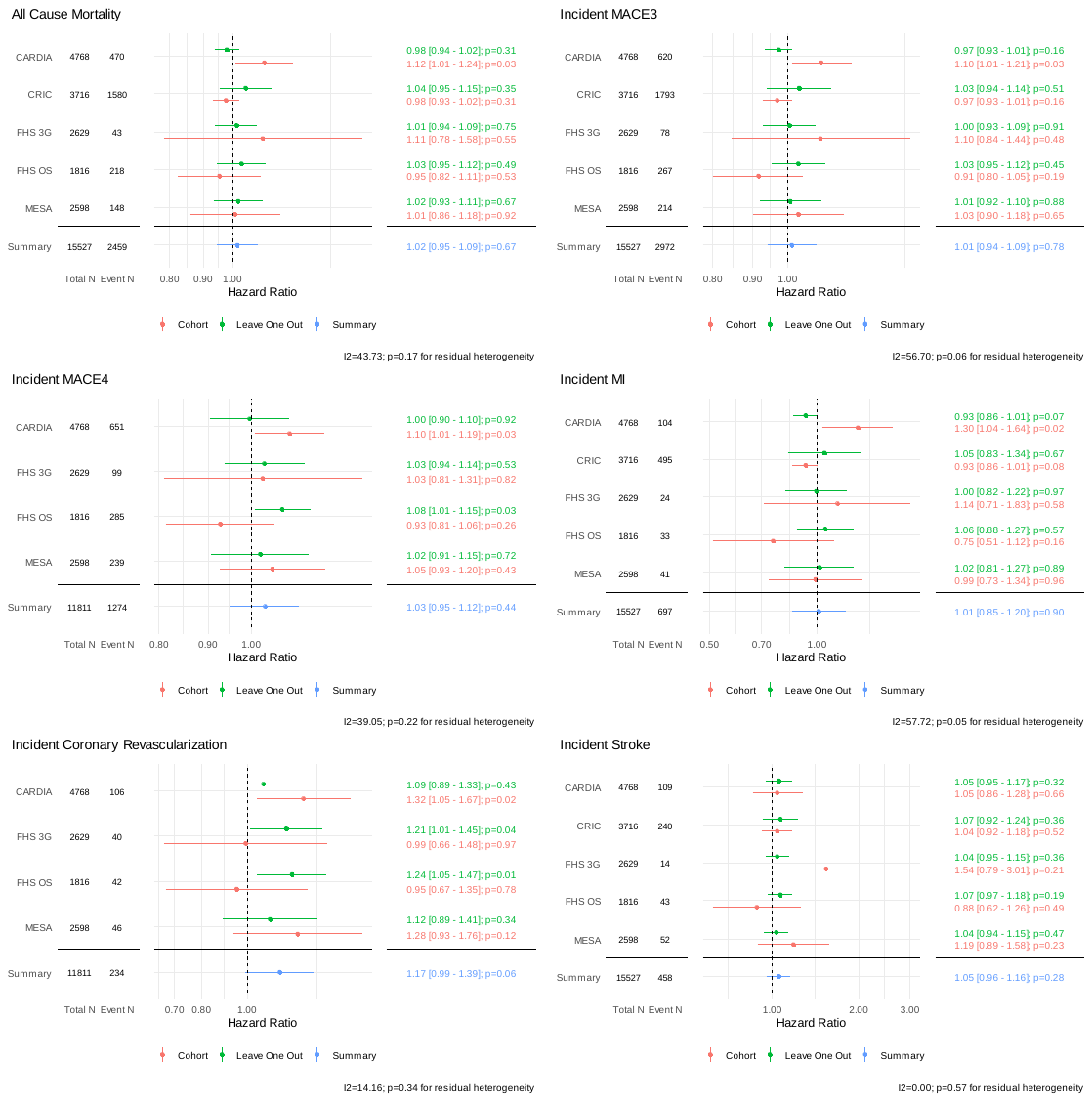

### Platelet Count (Fully Adjusted w/ hsCRP)

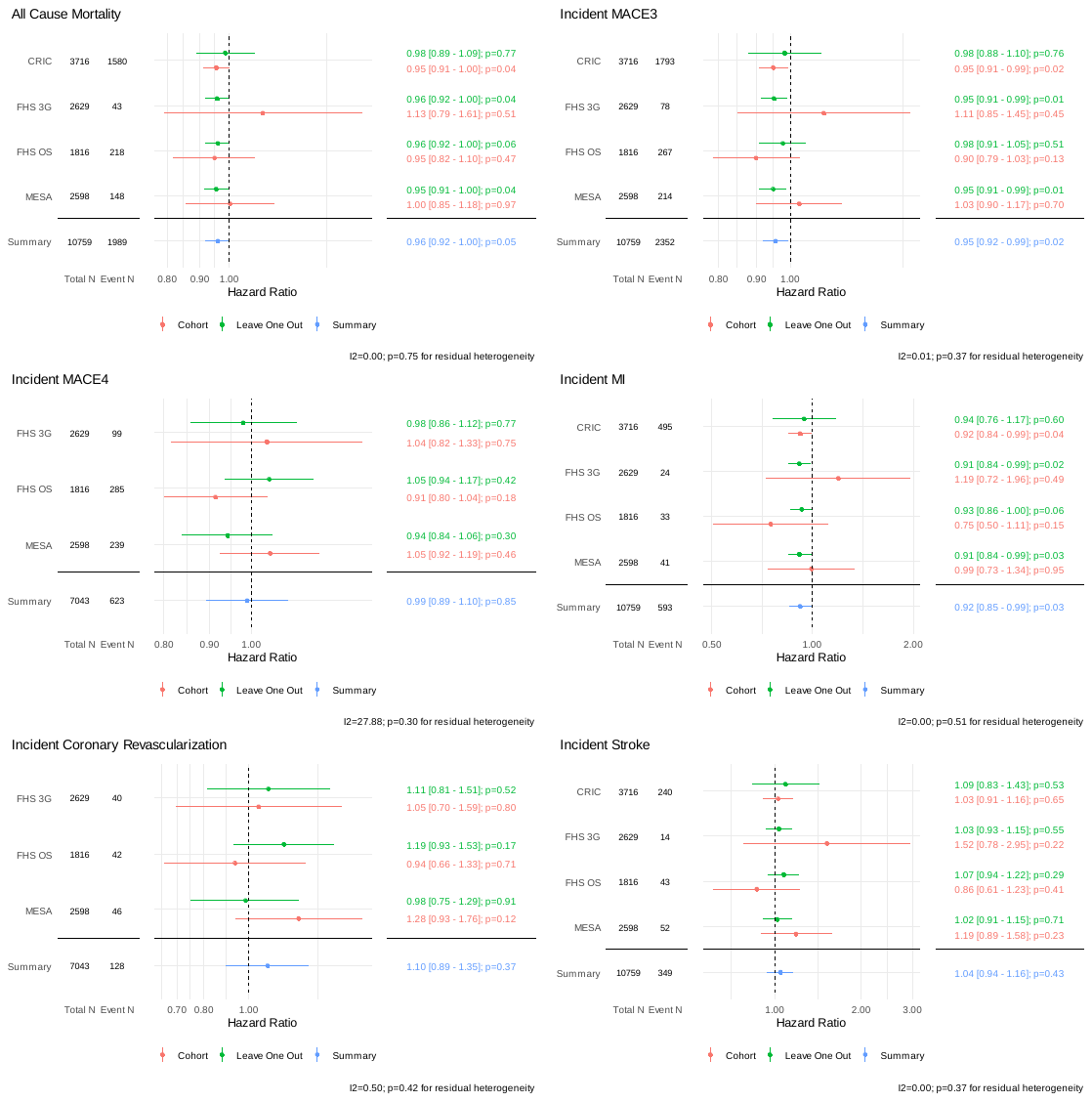

### Platelet Volume (Unadjusted)

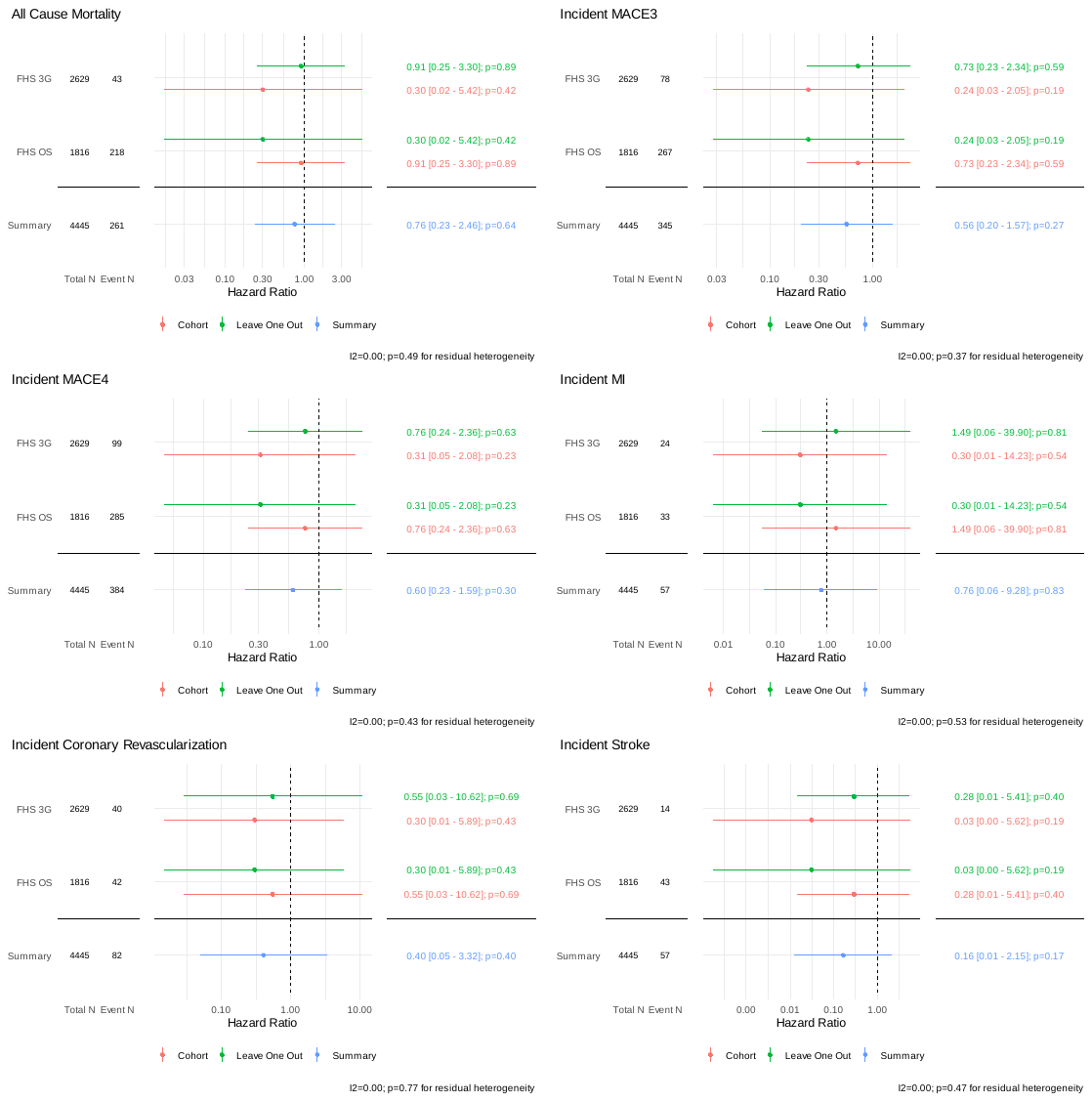

### Platelet Volume (Age/Sex/Race Adjusted)

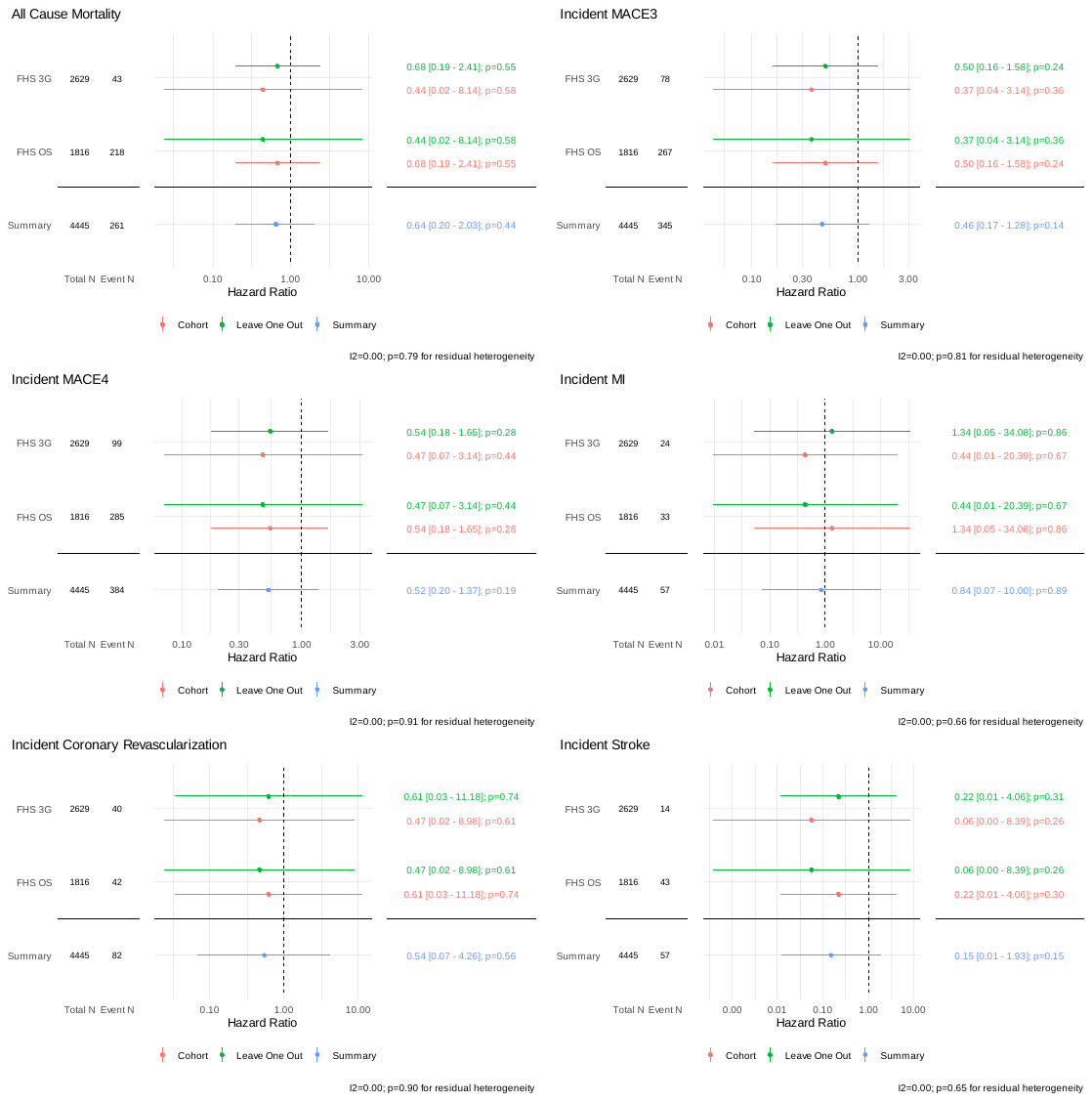

### Platelet Volume (Fully Adjusted)

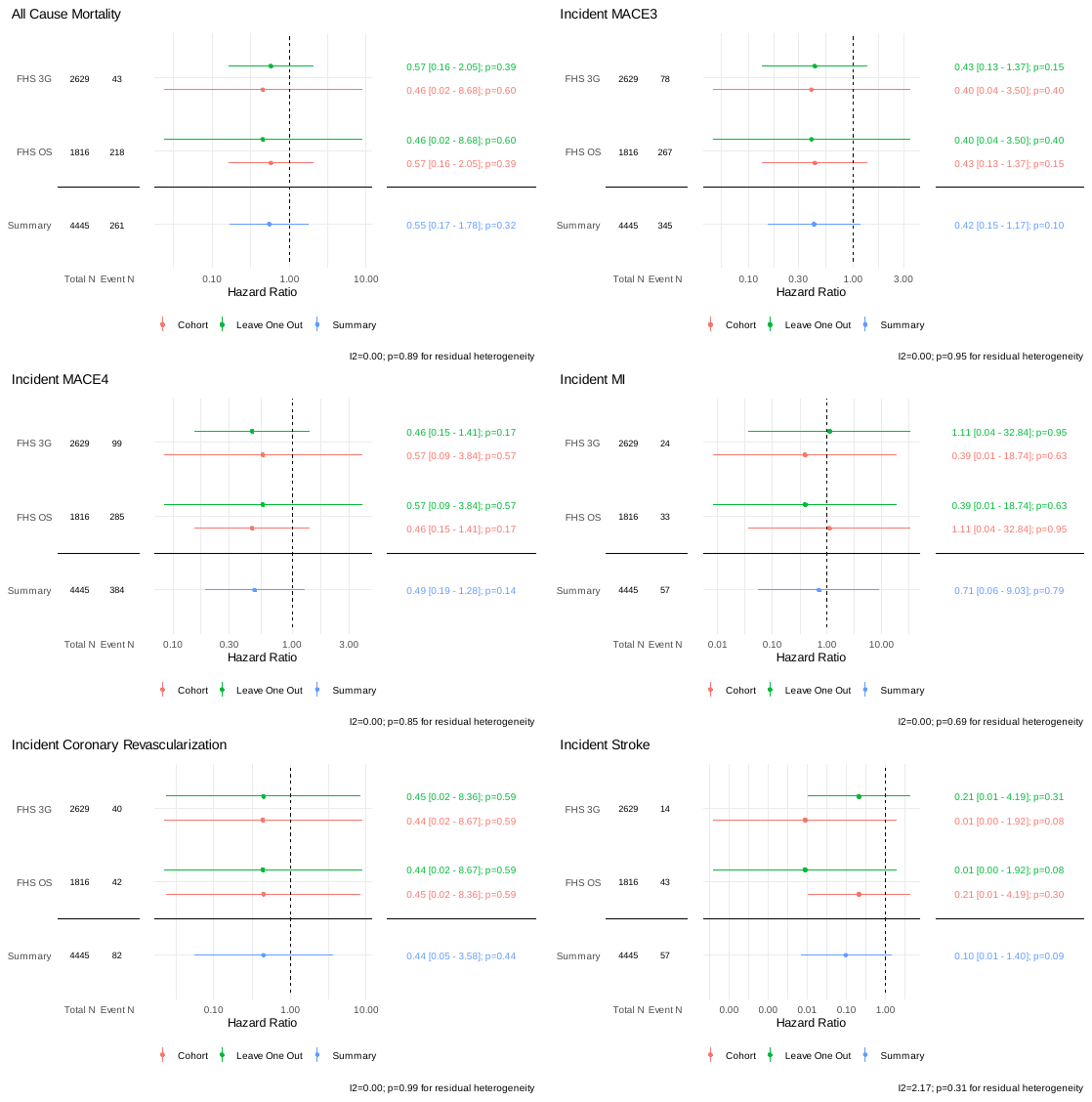

### Platelet Volume (Fully Adjusted w/ hsCRP)

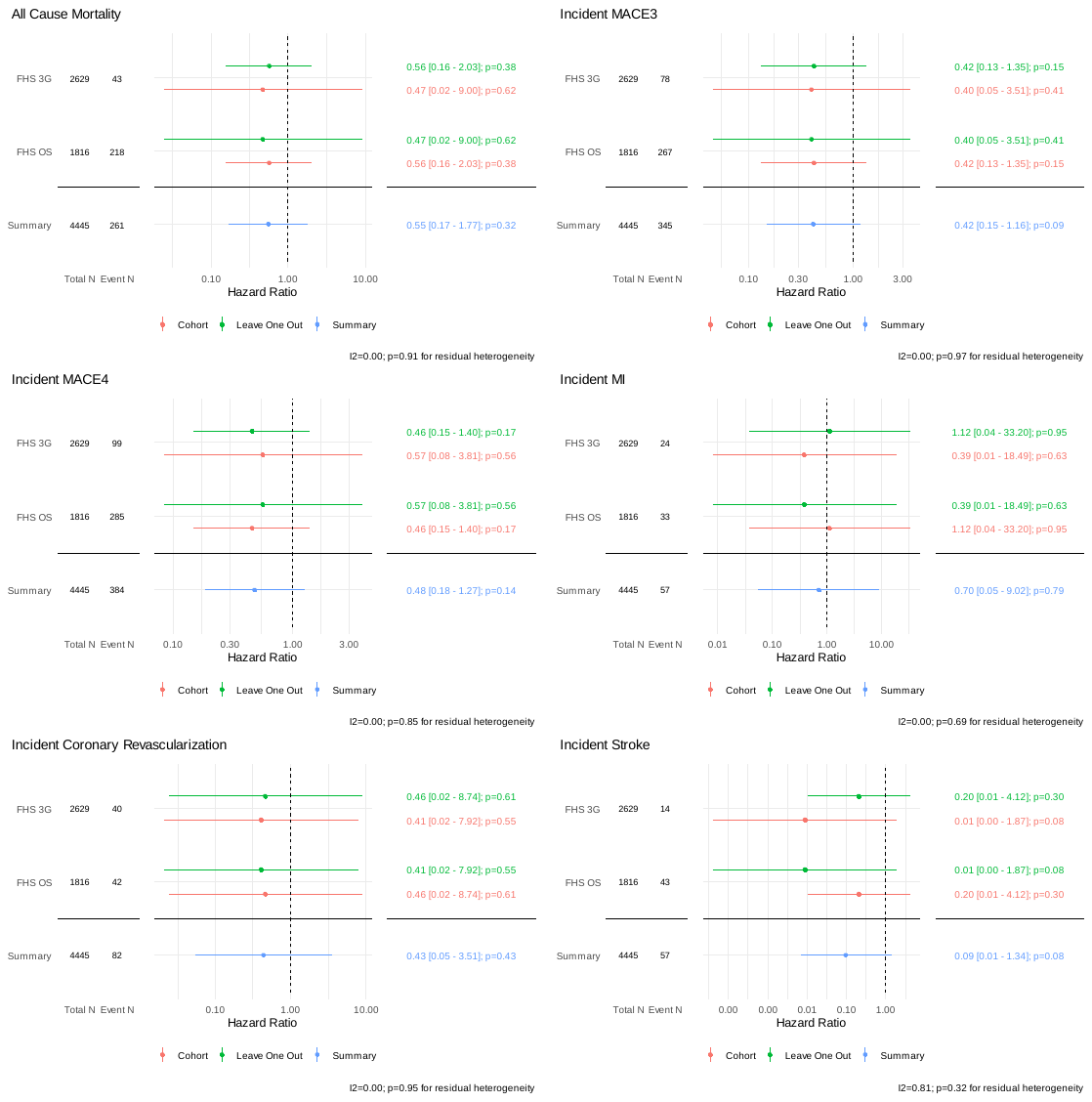

### RBC Count (Unadjusted)

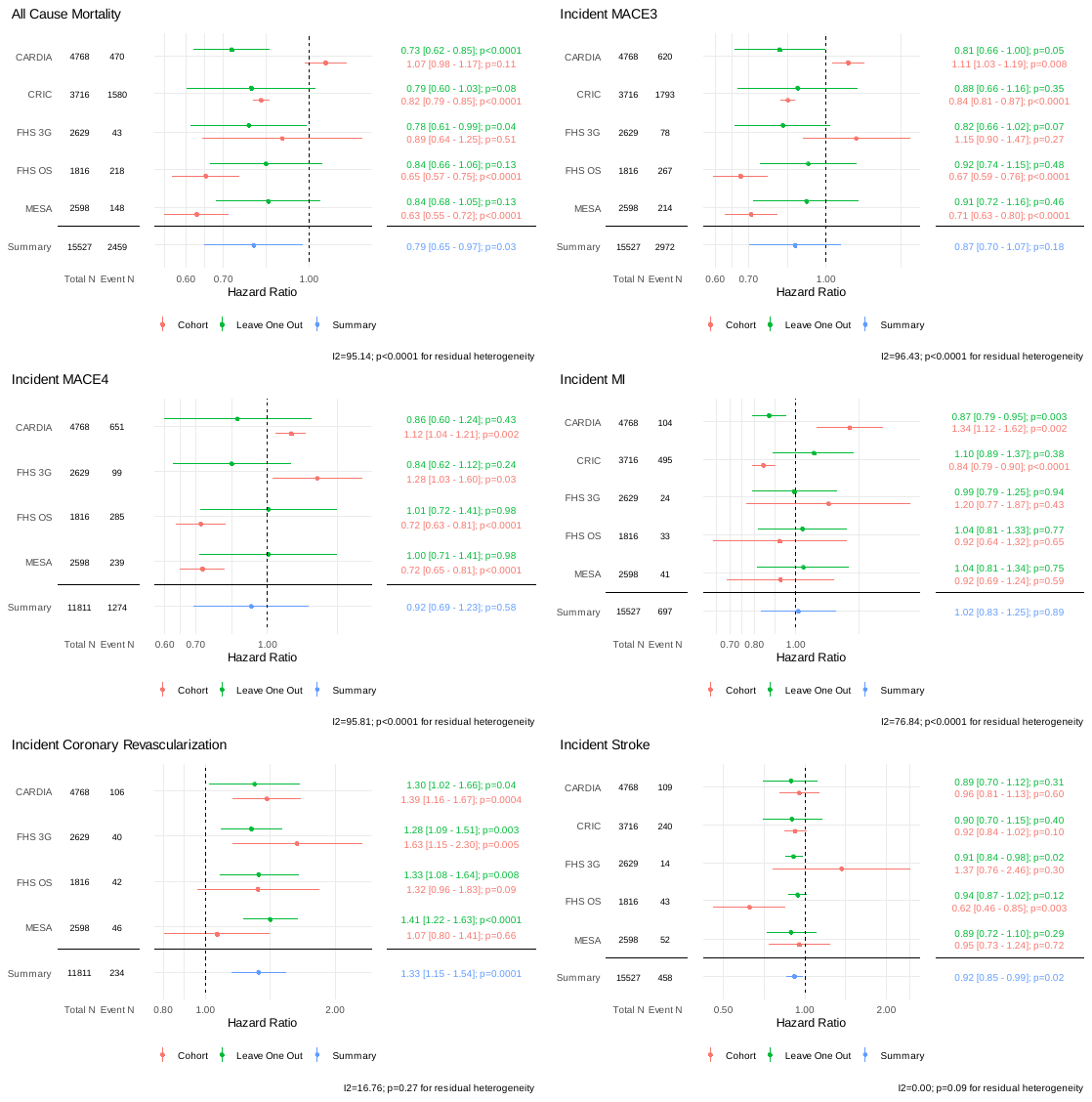

### RBC Count (Age/Sex/Race Adjusted)

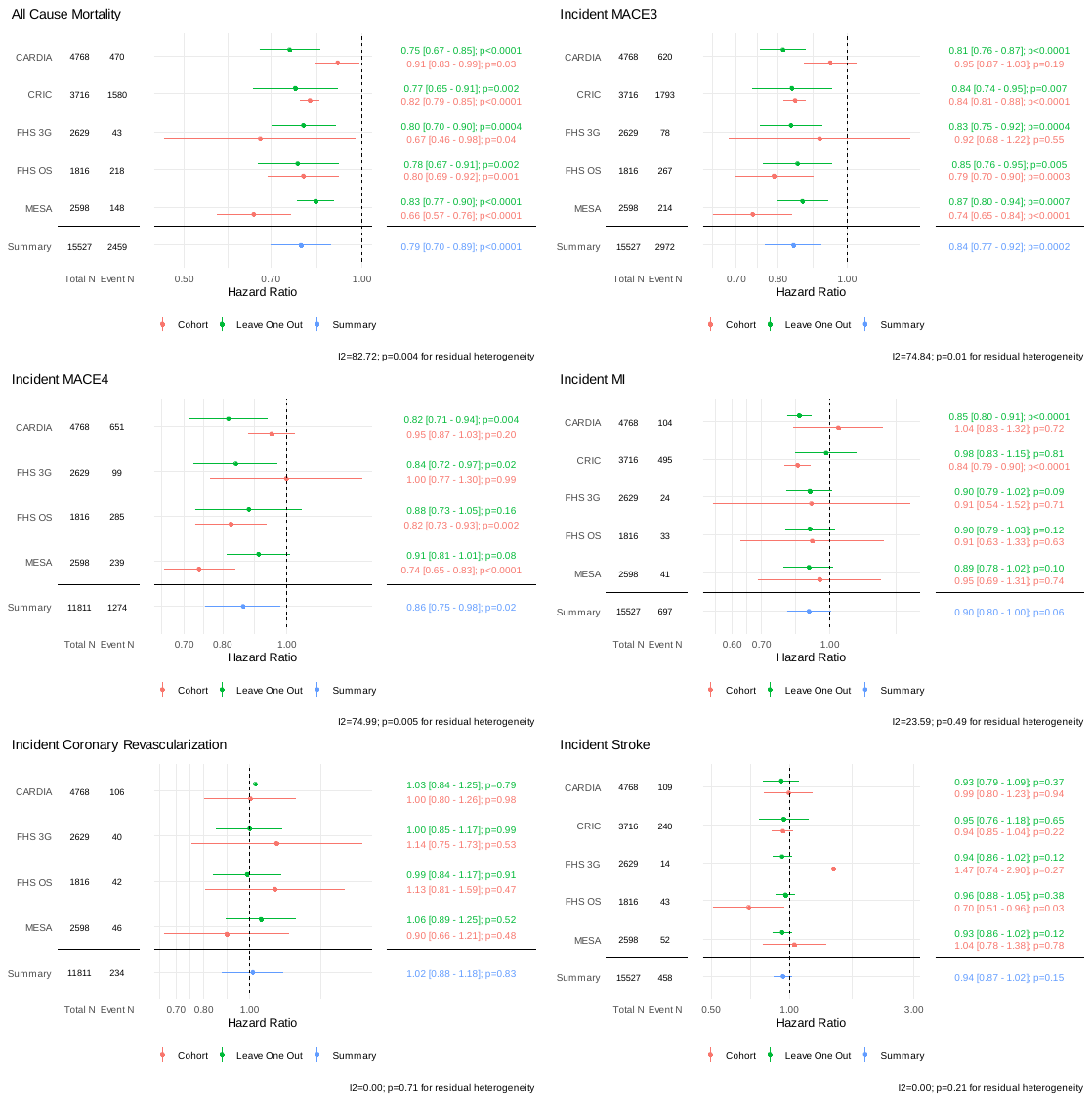

### RBC Count (Fully Adjusted)

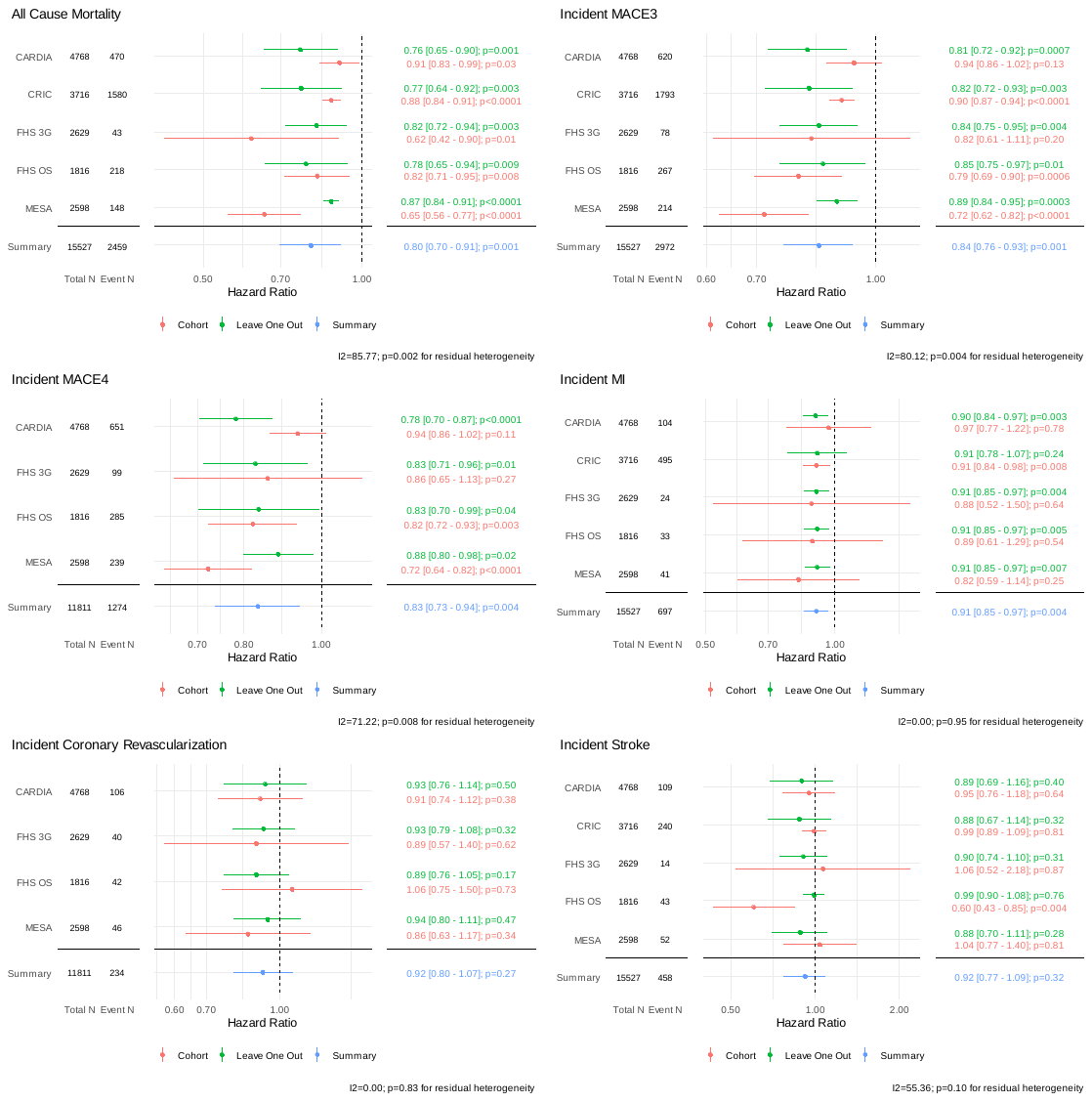

### RBC Count (Fully Adjusted w/ hsCRP)

### Red Cell Distribution Width (Unadjusted)

### Red Cell Distribution Width (Age/Sex/Race Adjusted)

### Red Cell Distribution Width (Fully Adjusted)

### Red Cell Distribution Width (Fully Adjusted w/ hsCRP)

### WBC Count (Unadjusted)

### WBC Count (Age/Sex/Race Adjusted)

### WBC Count (Fully Adjusted)

### WBC Count (Fully Adjusted w/ hsCRP)

**Supplemental Figure 4.  Forest plots for CBC-derived parameters for CV endpoints and all-cause mortality.** Forest plots of standardized β-coefficients of all CBC components and CBC-derived scores for CV endpoints and all-cause mortality. The tables on the left indicate the total subjects and events in each cohort. Hazard ratios (circles) and their 95% confidence intervals (bars) are displayed in different colors for individual cohort estimates (coral), summary meta-analysis (blue), and leave one cohort analysis sensitivity analyses (green). Cox models in each cohort were adjusted for age, sex, race, smoking, body mass index, systolic and diastolic blood pressure, use of antihypertensive medications, diabetes status, total cholesterol, HDL, LDL, and triglycerides.

**Supplemental Figure 5.  Impact of subclinical alterations of CBC-derived scores on all-cause mortality and MACE3 within the normal range.** Predicted hazard ratio (blue line) and 95% confidence interval (grey region) across percentiles of the Neutrophils & Lymphocytes score and RBC Quantity score. Hazard ratios were computed using meta-analyzed, fully adjusted ꞵ-Coefficients. Percentiles were calculated using survey-weighted mean and standard deviations from NHANES. Selected percentiles, all within two standard deviations of the mean, are highlighted.
